# Epidemiology of menstrual-related absenteeism in 44 low and middle-income countries

**DOI:** 10.1101/2024.05.07.24307016

**Authors:** Miranda Starr, Rebecca Harding, Ricardo Ataide, Naomi VonDinklage, Sheela S Sinharoy, Yasmin Jayasinghe, Lucinda Manda-Taylor, Jane Fisher, Sabine Braat, Sant-Rayn Pasricha

**Affiliations:** Population Health Immunity Division, Walter and Eliza Hall Institute of Medical Research, Parkville, VIC, Australia; Department of Medicine at the Peter Doherty Institute, University of Melbourne, Parkville, VIC, Australia; Hubert Department of Global Health, Rollins School of Public Health, Emory University, Atlanta, GA, 30322, USA; Department of Obstetrics, Gynaecology and Newborn Health, University of Melbourne, Royal Women’s Hospital, Parkville, VIC, Australia; Department of Gynaecology, Royal Children’s Hospital, Melbourne, VIC, Australia; Murdoch Children’s Research Institute, Melbourne, VIC, Australia; School of Global and Public Health, Kamuzu University of Health Sciencies, Private Bag 360, Blantyre, 3, Malawi; Global and Women’s Health, School of Public Health and Preventive Medicine, Monash University, 553 St Kilda Rd, Melbourne, Victoria, 3004, Australia; Diagnostic Haematology, The Royal Melbourne Hospital, Parkville, VIC, Australia; Clinical Haematology, The Peter MacCallum Cancer Centre and The Royal Melbourne Hospital, Parkville, VIC, Australia; Department of Medical Biology, University of Melbourne, Parkville, VIC, Australia

**Author notes:** **Corresponding author:** Professor Sant-Rayn Pasricha Population Health and Immunity Division, Walter and Eliza Hall Institute of Medical Research, Parkville, VIC 3052, Australia. Equal contribution (Miranda Starr, Rebecca Harding). Equal contribution (Sabine Braat, Sant-Rayn Pasricha).

## Abstract

**Background:** Menstrual-related absenteeism from work, school, or social activities is an important functional indicator of poor menstrual health that disrupts women and girls’ daily lives and exacerbates gender inequality. We sought to estimate the prevalence and factors contributing to menstrual-related absenteeism across low- and middle-income countries (LMICs).

**Methods:** We analysed cross-sectional data from 47 nationally representative Multiple Indicator Cluster Surveys from 2017-2023 comprising 3,193,042 individuals from 555,869 households across 44 countries. The outcome of interest was menstrual-related absenteeism from work, school, or social activities during the respondent’s last menstrual period. Independent factors included women’s age, household wealth index, use of menstrual materials (e.g. pads, tampons, or cloth), availability of a private place to wash at home during menstruation, and contraceptive use (hormonal and other). Univariable and multivariable associations between each factor and menstrual-related absenteeism were obtained using log binomial models. Prevalences and associations were pooled by geographical region and across all surveys using a random effects meta-analysis.

**Findings:** We included 673,380 women and girls aged 15-49 years in this analysis. The pooled global prevalence of menstrual-related absenteeism was 15·0% [95% CI: 12·7-17·3], with the highest prevalence in South Asia (19·7% [11·6-27·8]) and West and Central Africa (18·5% [13·5-23·5]). After pooling data across surveys, girls aged 15-19 years were found to endure a higher prevalence of menstrual-related absenteeism compared to older age-groups. There was no association between menstrual-related absenteeism and household wealth or the use of menstrual materials. In contrast, having a private place to wash at home was associated with an increased prevalence of menstrual-related absenteeism (global adjusted Prevalence Ratio [PR]: 1·25 [1·05-1·48]). Menstrual-related absenteeism was less prevalent in women and girls using contraception (global adjusted PR any contraception vs no contraception: 0·86 [0·82-0·90]), including those using hormonal contraception (global adjusted PR hormonal contraception vs non-hormonal or no contraception: 0·85 [0·78-0·93]).

**Interpretation:** Menstrual-related absenteeism is prevalent, especially in Asia and Africa, and particularly in adolescent girls. The age-independent protective effect of hormonal contraception suggests symptoms such as heavy menstrual bleeding or pain drive absenteeism. Improving access to private wash facilities outside the home and medical solutions for menstrual symptoms may reduce menstrual-related absenteeism, but further prospective research is urgently needed.

**Funding:** National Health and Medical Research Council, Australia.

**Research in Context:** *Evidence before this study:* Menstrual health is a historically understudied topic, with limited knowledge on the epidemiology of menstrual health issues and contributing factors. One consequence of poor menstrual health is menstrual-related absenteeism from school, work, or social activities, which can interfere with women’s quality of life and contribute to gender inequality. We searched PubMed, PsychINFO, Embase, and Medline for articles investigating menstrual-related absenteeism across low- and middle-income countries (LMICs) from data inception to April 2023, without language restrictions. Numerous qualitative studies have investigated women’s experience of menstrual health in LMICs, but there was a lack of epidemiological studies which are needed to understand the breadth of this issue. Studies were similarly focussed on single countries, and often on particular sub-populations such as adolescents and girls attending school. A global perspective on menstrual health is therefore lacking, requiring studies representative of the entire population of menstruating women. Additionally, many of the quantitative studies available were descriptive only, with no investigation of the associations between menstrual-related absenteeism and contributing factors. Where investigative studies were available, menstrual hygiene management was often the focus of analyses, with limited investigation of other factors which may contribute to menstrual health.

*Added value of this study:* In this study, we extracted data from 47 nationally representative Multiple Indicator Cluster Surveys from 44 different LMICs. Data on menstrual-related absenteeism was available for more than 673 000 women and girls aged 15-49 years of age, who were included in our study. This study broadens epidemiological knowledge surrounding menstrual health by estimating the prevalence of and investigating the factors relating to menstrual-related absenteeism including age, wealth, use of sanitary products, availability of private wash facilities at home, and contraception use, across a diverse array of countries. Our study was therefore able to compare the importance of these factors and identify priorities for future menstrual health research and interventions. Additionally, the standardised nature of MICS surveys allowed for comparisons between survey populations, facilitating a more global understanding of the menstrual health problem.

*Implications of all the available evidence:* Our work identified menstrual-related absenteeism as a common health concern for women and girls in LMICs, particularly in South Asia and parts of sub-Saharan Africa. Adolescents and young women were at the highest risk of menstrual-related absenteeism, hereby identifying this population as a key priority for future menstrual health programs and research. Our results suggest that private out-of-home sanitation facilities are lacking in LMICs, which has been cited in previous literature. However, more research is needed to determine what improvements are required to better facilitate menstrual health and hygiene. Additionally, our results identified hormonal contraception to be protective against menstrual-related absenteeism, which we hypothesize is due to improved menstrual symptoms. Future work is needed to better understand the importance of menstrual symptoms for women and girl’s quality of life, and the efficacy of different treatments for these symptoms.

## Introduction

Menstrual health encompasses the physical, mental, and social impacts of menstruation on a woman’s life.^1^ This involves the ability to manage menstrual hygiene through access to appropriate menstrual materials and water, sanitation, and hygiene (WASH) facilities. Additionally, menstrual health also encompasses access to appropriate information and education on menstruation, diagnosis and treatment of menstrual disorders, a respectful environment free from stigma, and the freedom to choose what activities to participate in while menstruating.^1^

Women and girls from low and middle-income countries (LMICs) may experience poor menstrual health, causing a range of physical, mental, and social consequences.^2^ These factors may culminate in menstrual-related absences from work, school, or social activities, disrupting women’s and girl’s lives. Menstrual-related absenteeism in the workplace is likely common, with Performance Monitoring and Accountability 2020 (PMA2020) surveys estimating 19%, 11%, and 17% of women and girls in Burkina Faso, Niger, and Nigeria, respectively, miss work due to menstruation.^3^ The same study observed similar rates of menstrual-related absenteeism from school. Other studies have identified much higher rates of absenteeism, with a national survey in Bangladesh reporting 41% of menstruating girls aged 11-17 years missed school due to menstruation.^4^ These disruptions have the potential to interfere with women and girl’s educational and career developments and pose a serious barrier to achieving Sustainable Development Goal 5: Achieve gender equality and empower all women and girls.^5^

There remains a critical gap in our understanding of the impact of suboptimal menstrual health. While previous reports have descriptively analysed the global distribution of menstrual health indicators from national health surveys, investigation of factors contributing to menstrual health outcomes, along with exploration of region-specific trends, would help inform future menstrual health research and interventions.^6^ We reasoned that menstrual-related absences could serve as an indicator of poor menstrual health. Here, we leveraged data from Multiple Indicator Cluster Surveys (MICS) to estimate the prevalence of menstrual-related absenteeism from work, school, or social activities across LMICs, and to explore potential risk factors for this outcome.

## Methods

### Study design

We undertook a multi-national analysis of cross-sectional MICS data.^7^ MICS surveys measure indicators of women’s and children’s health, and have been conducted in 120 countries since the mid-1990s. MICS surveys follow a standardised study design, incorporating multi-stage sampling to select a nationally representative sample of households. Members of selected households are then interviewed using questionnaires directed at the household overall, and questionnaires addressing specific household members (men 15-49 years, women 15-49 years, children <5 years, children 5-17 years). Our study’s outcome data was derived from the women’s questionnaire delivered to consenting women and girls aged 15-49 years in the sampled households. The household dataset was also utilised to capture broader demographic factors.

STROBE guidelines were used in preparing this report and the GATHER checklist is presented in the appendix (p 2).^8^ All MICS surveys used in this study had previously undergone ethical review before the commencement of fieldwork. As this study was a secondary analysis of de-identified data, no additional ethics approval was required. Approval from UNICEF for data access was received before analysis.

### Procedures

The outcome of interest for this study was MICS indicator WS.13, in which women and girls who menstruated in the last 12 months were asked, ‘Due to your last menstruation, were there any social activities, school or workdays that you did not attend?’ Possible responses included ‘yes,’ ‘no,’ and ‘don’t know/no such activity.’ This indicator was added to the sixth round of MICS in 2017, and as of September 2023, data was available for 48 surveys across 44 countries.^7^

We first developed hypotheses for associations with menstrual-related absenteeism based on current literature. We expected menstrual-related absenteeism to be more common in young women and girls who may have had less menstrual education, may have lesser empowerment, and may experience more severe menstrual symptoms.^9–11^ Women and girls in the lower wealth quintiles were hypothesised to have reduced access to menstrual-related resources and, therefore, may suffer from more menstrual-related absenteeism, along with women and girls who did not use any menstrual materials.^12^ Women and girls with private facilities to wash and change menstrual materials at home were hypothesised to experience higher menstrual-related absenteeism as they may be better able to meet their menstrual health needs at home compared to at work or school. Lastly, women and girls who used hormonal contraception were expected to experience less severe menstrual symptoms and, therefore, less menstrual-related absenteeism.^13,14^

Based on our pre-specified hypotheses we selected independent variables to reflect factors potentially influencing menstrual health: women’s age (5-year age groups from 15-49 years), household wealth (quintile 1 (poorest) to quintile 5 (richest)), use of menstrual products (pads, tampons, or cloth), availability of a private place to wash and change menstrual materials at home during menstruation, and contraceptive use (appendix p 3-4).

Household wealth quintiles were developed by MICS for each survey using a wealth index based on principal component analyses of household characteristics such as dwelling characteristics, water and sanitation, and ownership of consumer goods. As such, the quintiles represent the relative wealth *within* the survey population. Contraception use was included due to the efficacy of hormonal contraceptives for improving menstrual pain and heavy menstrual bleeding.^13,14^ It was categorised in two different ways: current use of any contraception, and current use of hormonal contraception defined as the use of at least one of the following methods: injectables, implants, or the pill. Women and girls using an intra-uterine device (IUD) were excluded from hormonal contraception analyses as questionnaire items did not differentiate between the use of copper or progesterone-releasing IUDs, which could have divergent effects on menstrual symptoms. Area type (urban/rural) and reusable sanitary materials were also investigated as independent variables (appendix p 28-29, 46-47, 50-51).

### Statistical analysis

The distribution of demographic factors and independent variables was described for each survey cohort (appendix p 6-25, 27-31). The prevalence of menstrual-related absenteeism in each survey population was calculated for the entire survey sample and stratified by the independent variables of interest (appendix p 26, 32-34, 46-57). Using log-binomial regression, the univariable association between each independent variable and menstrual-related absenteeism was estimated as prevalence ratios (PR) with associated 95% confidence intervals (CI). If any convergence issues arose, a logistic regression model was fitted followed by the marginal standardisation technique to estimate the PR and the delta method for the standard error and CI.^15^

Multivariable associations with menstrual-related absenteeism were also obtained for each independent variable of interest using the same methods, with adjustment for relevant confounders informed by directed acyclic graphs (DAGs) created using Daggity (appendix p 35).^16^ The adjustment sets consisted of women’s age and marital status (currently, formerly, or never married/in union) for household wealth; household wealth, women and girl’s highest level of education (pre-school or none, primary, secondary, higher or vocational), and area type for use of sanitary menstrual products; household wealth and area type for a private place to wash and change at home; household wealth, area, women’s age, and educational attainment for use of contraception (any method or hormonal methods). Our DAGs identified no confounders for the association between age and menstrual-related absenteeism. However, area type was included as a confounder for age due to its potential to determine life expectancy and contribute to menstrual-related absenteeism.^17,18^

A two-step random effects meta-analysis with the Sidik-Jonkman method was used to generate pooled prevalence estimates and prevalence ratios, and associated 95% confidence intervals, with results pooled across MICS-defined geographical regions and across all surveys.^19–21^ Our study included four surveys conducted in separate Pakistan provinces, which were treated as independent in all meta-analyses.

The population used for demographic analyses and prevalence estimates included all women and girls without missing data for the outcome variable (i.e., who responded ‘yes’, ‘no’, ‘don’t know/no such activity’ or ‘no response’) and relevant independent variables. When undertaking univariable and multivariable analyses, the analysis set was reduced to only women and girls who answered ‘yes’ or ‘no’ to the outcome variable and relevant independent variables.

All analyses were completed in Stata 17 (StataCorp. 2021. College Station, TX: StataCorp LLC) using the SVY commands to account for complex survey design using women’s survey weights, with stratification by area type and district, region, or province depending on the survey. META commands were also used for pooling results. Strata with single sampling units were centred at the grand mean to allow the calculation of standard errors.^22^ No finite population correction was applied due to the unknown population size of women 15-49 years in each survey population. The sampling variability might therefore be overestimated in some surveys, in particular in countries with small population size. Some figures were additionally produced using R Statistical Software (v4.3.1; R core team 2023) or QGIS 2.10 Pisa software.

## Results

An overview of all surveys and their sample demographics is shown in the appendix (p 5-25). One eligible survey (Democratic People’s Republic of Korea, 2017) was excluded due to restricted data, leading to a sample of 47 MICS surveys conducted across 44 countries from May 2017 to February 2023. Surveys were conducted across a range of geographical regions, with ten surveys in West and Central Africa, 4 in Eastern and Southern Africa, 4 in the Middle East and North Africa, 7 in South Asia, 8 in East Asia and the Pacific, 7 in Europe and Central Asia, and 7 in Latin America and the Caribbean (Figure 1). In total, these surveys comprised 3,193,042 individuals from 555,869 households, of which 673,380 women and girls from 479,424 households had information for the outcome variable and were included in this study. The total survey-weighted sample size was 677,035 women and girls, with survey-weighted populations ranging from 728 in Tuvalu to 58,198 in Bangladesh (appendix p 5).

**Figure 1:**
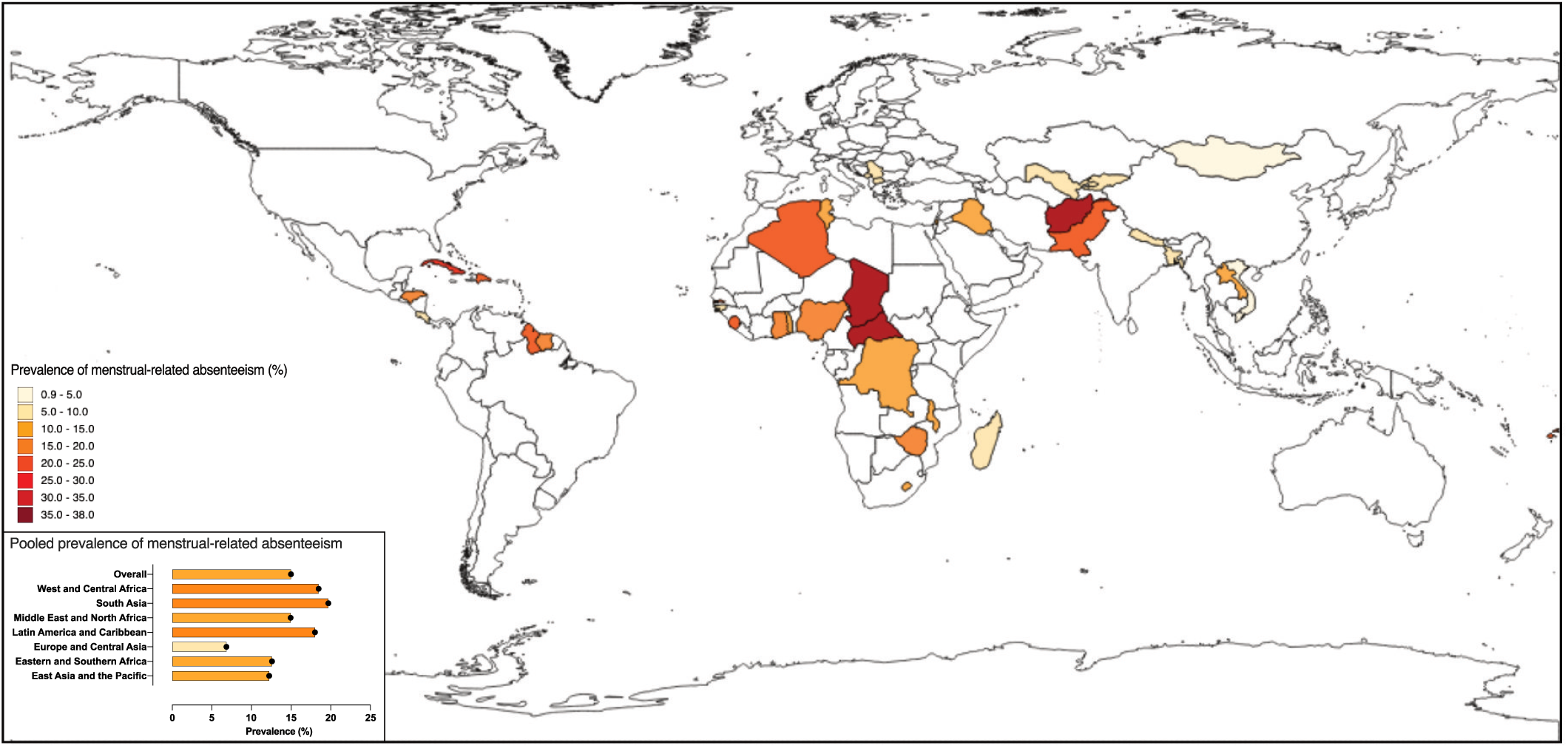
Prevalence of menstrual-related absenteeism by survey population. Each MICS survey country is coloured to represent the prevalence of menstrual-related absenteeism in surveyed women and girls aged 15-49 years. Countries shown in white have no available MICS6 survey. The bar graph represents the prevalence of menstrual-related absenteeism pooled by region, and an overall pooled estimate across all surveys. Certain pacific countries are not shown but include; Kiribati (prevalence of menstrual-related absenteeism = 16.1%), Samoa (9.0%), and Tonga (15.6%). Pakistan had four region-specific surveys from which results were pooled to generate a country-specific estimate (22.6%).

Across all surveys, the pooled prevalence of menstrual-related absenteeism was 15·0% [95% CI: 12·7-17·3] (appendix p 26). The region-pooled prevalence of menstrual-related absenteeism was highest in South Asia (19·7% [11·6-27·8]), followed by West and Central Africa (18·5% [13·5-23·5]), Latin America and Caribbean (18·0% [13·1-22·9]), Middle East and North Africa (14·9% [8·7-21·1]), Eastern and Southern Africa (12·6% [9·4-15·8]), East Asia and the Pacific (12·2% [7·6-16·9]), and Europe and Central Asia (6·8% [4·6-9·1]) (figure 1, appendix p 26). The prevalence of menstrual-related absenteeism by country ranged from 0·9% in Turkmenistan to 38·0% in the Sindh province of Pakistan. The prevalence of menstrual-related absenteeism across each independent variable is described in detail in the appendix (p 32-34, 46-57).

We first investigated the association between women’s age and menstrual-related absenteeism. The average age of women and girls across all surveys was 30·0 years [95% CI: 29·4-30·6] (appendix p 27). In 35 of 47 surveys, menstrual-related absenteeism was highest in women and girls aged 15-19 or 20-24 years (appendix p 32-33). In these age-groups the overall pooled prevalence of menstrual-related absenteeism was 17·7% [15·1-20·3] in 15-19 year olds and 16·2% [13·8-18·6] in 20-24 year olds. After adjusting for area type, the overall pooled prevalence ratios indicated that 15-19 year-olds (reference group) had the highest prevalence of menstrual-related absenteeism when compared to all other age groups, with pooled prevalence ratios ranging from 0·75 [0·68- 0·82] in 35-39 year-olds to 0·92 [0·87-0·97] in 20-24 year-olds (figure 2, appendix p 39-41).

**Figure 2:**
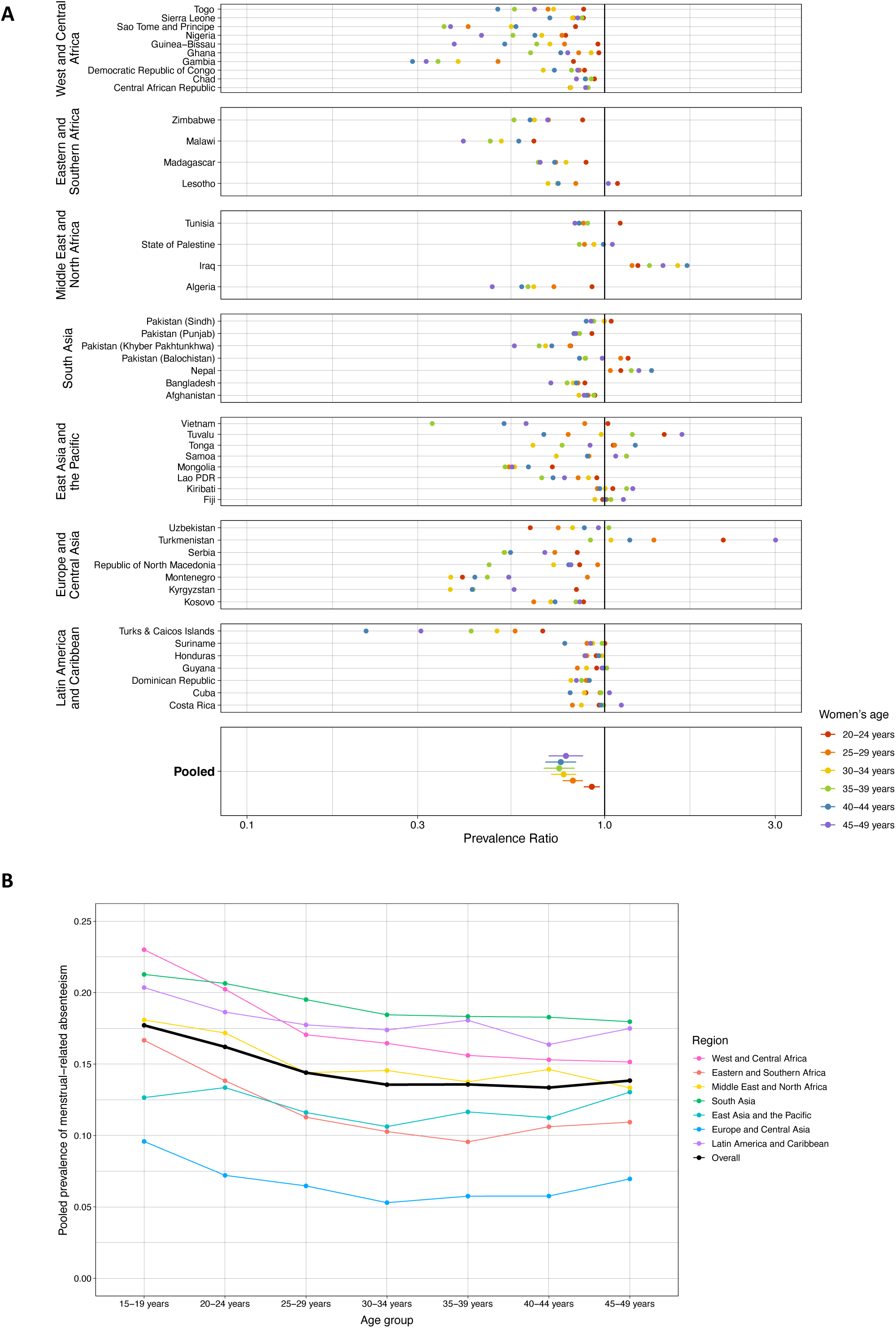
Association between age and menstrual-related absenteeism. (A) Dot plot with survey-specific prevalence ratios comparing the prevalence of menstrual-related absenteeism across different age groups to 15-19 year-olds as the reference group. A vertical line highlights the null prevalence ratio of 1. Prevalence ratios below this value indicate a higher prevalence of menstrual-related absenteeism in the reference group (15-19 year-olds), while prevalence ratios above this value indicate a higher prevalence in the non-reference age group. All prevalence ratios are adjusted for area type (urban/rural). (B) Line graph showing the prevalence of menstrual-related absenteeism pooled by region and age group. The pooled prevalence across all surveys is shown by the black line.

Next, we compared the prevalence of menstrual-related absenteeism across household wealth quintiles, with the reference being the third (median) quintile (figure 3). After adjusting for age and marital status, there was no definite evidence of an association between wealth index and menstrual-related absenteeism (Quintile 1 [poorest]: adjusted PR −1·09 [95% CI: 1·00-1·19]; Quintile 2: 1·01 [0·96-1·06]; Quintile 4: 0·94 [0·89-1·00]; Quintile 5 [richest]: 0·91 [0·83-0·99]) (appendix pp 44-45).

**Figure 3:**
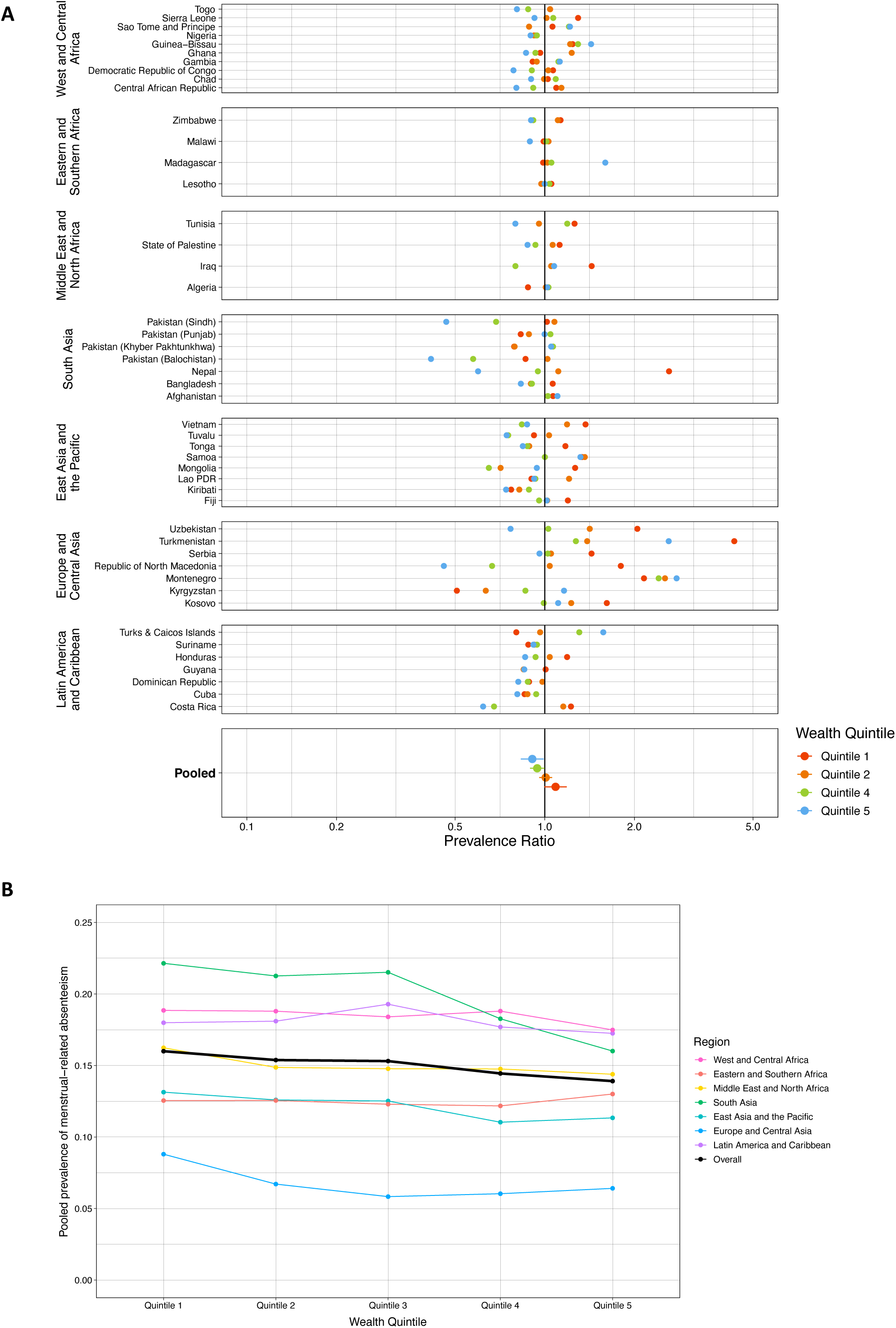
Association between household wealth quintile and menstrual-related absenteeism. (A) Dot plot with survey-specific prevalence ratios comparing the prevalence of menstrual-related absenteeism across different household wealth quintiles, with the third (median) quintile as the reference. A vertical line highlights the null prevalence ratio of 1. Prevalence ratios below this value indicate a higher prevalence of menstrual-related absenteeism in the reference group (quintile 3), while prevalence ratios above this value indicate a higher prevalence of menstrual-related absenteeism in the non-reference wealth quintile. (B) Line graph showing the prevalence of menstrual-related absenteeism pooled by region and wealth quintile. The pooled prevalence across all surveys is shown by the black line. Wealth quintiles represent the relative wealth within a country and are derived from household wealth scores calculated based on housing characteristics and household and personal assets. Quintile 1 represents the poorest households, while quintile 5 represents the richest.

Likewise, we observed no consistent association between the use of menstrual materials and menstrual-related absenteeism within or across regions, with an overall pooled prevalence ratio of 0·97 [95% CI: 0·85-1·11] after adjusting for wealth, education, and area type (figure 4A, appendix p 48-49). However, country-specific associations were observed, with the use of menstrual materials associated with lower absenteeism in countries such as Nepal (adjusted PR: 0·46 [0·37-0·57]) but higher absenteeism in countries such as Chad (2·01 [1·64-2·46]).

**Figure 4:**
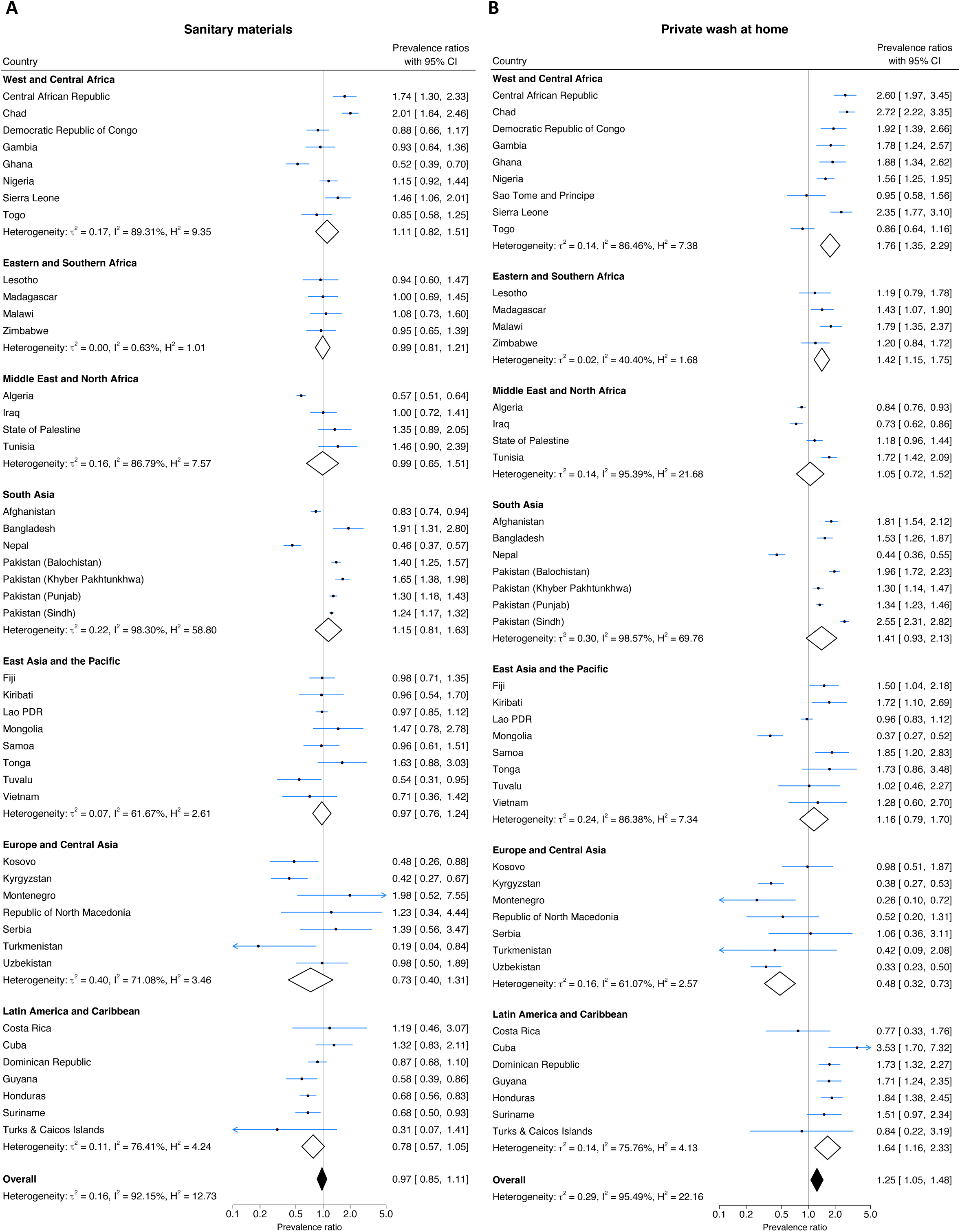
Association between menstrual-related absenteeism and use of menstrual materials (A) or availability of a private place to wash at home during menstruation (B). (A) Forest plot showing prevalence ratios comparing menstrual-related absenteeism in women and girls using menstrual materials compared to those using no menstrual materials (reference). Prevalence ratios below the null value of 1 favour using menstrual materials, while prevalence ratios above the null value favour using no menstrual materials. Analyses are adjusted for wealth, education, and area type. (B) Forest plot showing prevalence ratios comparing the prevalence of menstrual-related absenteeism in women and girls who have a private place to wash at home during menstruation to those who do not (reference). Prevalence ratios below the null value of 1 favour having a private place to wash at home, while prevalence ratios above the null value favour not having a private place to wash at home. Analyses are adjusted for wealth and area. Region-pooled prevalence ratios are shown as diamonds with a white fill. The overall pooled prevalence ratio is shown as the black-filled diamond. Measures of heterogeneity within regions and across all surveys are captured by 𝑟^2^, I^2^, and H^2^ alongside pooled estimates.

We next investigated the association between menstrual-related absenteeism and having a private place to wash and change at home during menstruation. The pooled prevalence of women and girls with a private place to wash and change at home during menstruation ranged from 78·9% [95% CI: 63·5-94·3] in the Middle East and North Africa to 97·3% [95·9-98·8] in Europe and Central Asia, with an overall pooled prevalence of 91·6% [89·2, 93·9] (appendix p 30). Having a private place to wash and change at home was associated with a higher prevalence of menstrual-related absenteeism, with an overall pooled prevalence ratio of 1·25 [1·05, 1·48] after adjusting for wealth and area type (figure 4B, appendix p 52-53). This trend was observed in region-pooled estimates in West and Central Africa, Eastern and Southern Africa, and Latin America and the Caribbean, though an association in the opposite direction was observed in Europe and Central Asia (adjusted PR: 0·48 [0·32-0·73]).

Finally, we explored the association between contraception use and menstrual-related absenteeism. The region-pooled proportion of women and girls using any method of contraception varied from 24·1% [95% CI: 18·3-29·8] in West and Central Africa to 60·2% [54·6-65·7] in the Middle East and North Africa, with an overall pooled prevalence of 38·6% [33·8-43·5] (appendix p 31). Contraception use was associated with lower menstrual-related absenteeism, with an overall pooled prevalence ratio of 0·86 [0·82-0·90] after adjusting for age, wealth, education level, and area type (Figure 5A, appendix p 54-55). Contraception use was associated with a reduced prevalence of menstrual-related absenteeism in 18 of 47 surveys, with no surveys demonstrating increased menstrual-related absenteeism in contraception users.

**Figure 5:**
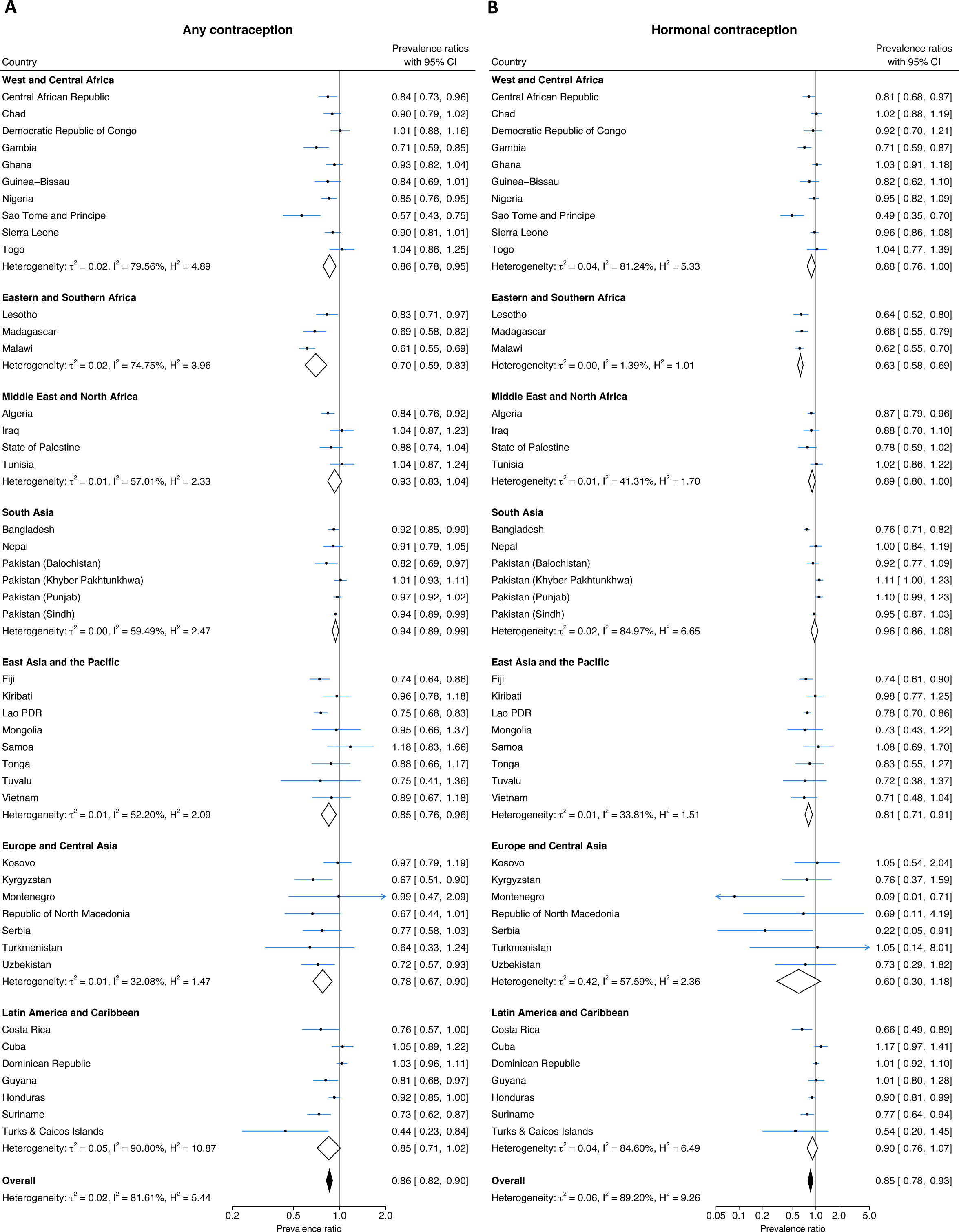
Association between contraception use and menstrual-related absenteeism. (A) Forest plot showing prevalence ratios comparing menstrual-related absenteeism in women and girls using contraception compared to those not using contraception. Prevalence ratios below the null value of 1 favour contraception use, while prevalence ratios above the null value favour the use of no contraception. (B) Forest plot showing prevalence ratios comparing menstrual-related absenteeism in women and girls using hormonal contraception compared to those using only non-hormonal or no contraception. Prevalence ratios below the null value of 1 favour the use of hormonal contraception, while prevalence ratios above the null value favour the use of only non-hormonal or no contraception. All analyses are adjusted for women’s age, wealth, education, and area. Region-pooled prevalence ratios are shown as diamonds with a white fill. The overall pooled prevalence ratio is shown as the black-filled diamond. Measures of heterogeneity within regions and across all surveys are captured by 𝑟^2^, I^2^, and H^2^ alongside pooled estimates.

The region-pooled proportion of women and girls on contraception who were using a hormonal method ranged from 11·4% [95% CI: 5·3-17·5] in Europe and Central Asia to 78·0% [66·4-89·7] in Eastern and Southern Africa, with an overall pooled prevalence of 52·2% [44·9-59·5] (appendix p 31). After adjusting for age, wealth, education, and area type, the use of hormonal contraception was associated with lower menstrual-related absenteeism when compared to the use of only non-hormonal or no contraception (overall adjusted PR: 0·85 [0·78-0·93]) (Figure 5B, appendix p 56-57). Hormonal contraception was associated with a reduced prevalence of menstrual-related absenteeism in 15 of 47 surveys, with no surveys observing an increase in menstrual-related absenteeism in hormonal contraception users.

## Discussion

Our analysis of multinational data from 44 low- and middle-income countries summarises the prevalence, highlights the burden, and uncovers factors associated with menstrual-related absenteeism from work, school, or social activities. Menstrual-related absenteeism occurs in 15% of 15-49 year-old women and girls globally, and is most common in South Asia and parts of sub-Saharan Africa. Menstrual-related absenteeism prevalence is highest among younger women, emphasising the vulnerability of this group and the urgent need for adolescent girls to be prioritised in future research and programs. Women and girls who use hormonal contraception appear to report less menstrual-related absenteeism independent of age, suggesting that menstrual symptoms drive this outcome. Our analyses provide new evidence to inform future menstrual health research and public health interventions.

We interpret menstrual-related absenteeism as an indicator of suboptimal menstrual health impacting a woman’s health or social functioning. Both any contraception and hormonal contraception specifically were associated with reduced menstrual-related absenteeism in this study, however the high level of similarity between these results suggests that hormonal methods may be driving this effect. Hormonal contraception improves many menstrual symptoms, including heavy menstrual bleeding and menstrual pain.^13,14,23,24^ Both symptoms have been shown to contribute to menstrual-related absenteeism in adolescents in countries such as Malawi (47·9% absent due to heavy bleeding, 19·5% due to cramps/headache/pain), India (31·8% absent due to excessive bleeding, 76·3% absent due to pain/discomfort), and Ghana (40·0% absent due to heavy bleeding, 57·3% absent due to pain).^25–27^ Additionally, the 2018 Mongolia MICS survey measured reasons for menstrual-related absenteeism and found 76·4% of women and girls were absent due to menstrual pain, with 18·6% absent due to heavy bleeding. The association between hormonal contraceptive use and reduced menstrual-related absenteeism may therefore be mediated by menstrual symptom control, indicating the potential value of such treatments as a possible menstrual health intervention independent from the benefits to family-planning. Where hormonal contraception as a menstrual health intervention is culturally inappropriate in unmarried adolescents or women, programs may be needed to increase awareness among healthcare providers and caregivers of the benefits of hormonal contraception for treatment of menstrual disorders. In parallel, improved access to other medical treatments such as non-steroidal anti-inflammatory drugs or tranexamic acid could improve menstrual symptoms and alleviate absenteeism.

Socioeconomic disadvantage is considered an important contributor to poor menstrual health. However, our study did not find clear associations between relative household wealth and menstrual-related absenteeism. Similar results have been observed in local cross-sectional and mixed methods studies assessing menstrual-related absenteeism across asset-based wealth groups.^28,29^ One possible explanation may be that wealth quintiles measure a household’s relative wealth within a population; if the population are largely poor or wealth is similar across a large proportion of the population, then the composition of quintiles across the population may be similar, limiting the utility of comparison between quintiles. Another explanation may be that wealth does not guarantee improved access to menstrual materials and WASH facilities, for example in rural areas with limited resources. Additionally, women and girls may not have access to their household’s wealth or the agency to spend such money on menstrual materials. Further work is needed to explore these possibilities.

We found no clear association between the use of menstrual materials and menstrual-related absenteeism. Similar results were observed in a Ugandan study, where the daily use of disposable pads was not significantly associated with menstrual-related absenteeism from school.^30^ However, others have identified a lack of access to pads as a risk factor for absenteeism.^12^ Other unexplored factors related to the use of menstrual materials include product quality and sufficiency, availability of disposal facilities, and private areas for changing and washing outside the home, which are important for a woman’s menstrual hygiene security.^31,32^ These factors are often neglected in programs and are not currently captured in large-scale health surveys such as MICS or DHS. Further studies are needed better to understand their contribution to poor menstrual hygiene and health.

Having a private place to wash and change at home during menstruation was associated with an increased prevalence of menstrual-related absenteeism in many regions. This implies limited access to out-of-home private wash facilities, causing women and girls to remain home when menstruating to better manage their hygiene. A lack of supportive infrastructure for menstrual health and hygiene is common in schools and workplaces in LMICs.^2^ The implementation of female-friendly toilets could help overcome this issue and may be especially beneficial to women and girls experiencing heavy menstrual bleeding who need to wash and change materials frequently.^33,34^ Future studies are needed to investigate the impacts of improved, female-friendly WASH facilities in schools and workplaces on women’s health, education, employment, and quality of life.

Our study has several limitations. By design, this study leveraged data from the MICS surveys, so we could not obtain further information surrounding women’s menstrual health, including the direct self-reported causes of absenteeism and the prevalence and impact of menstrual symptoms. Likewise, unmeasured confounders such as women’s menstrual education, stigma, and other social or cultural determinants, may have influenced our findings. The cross-sectional design of MICS additionally makes causality challenging to confirm. Although MICS surveys apply a consistent methodology, differences in survey implementation across countries may have also contributed to the observed heterogeneity, making it difficult to discern significant trends or outliers. Face-to-face interviews to collect data on sensitive topics such as menstrual health and contraceptive use may have variably influenced answers and, hence, our analyses between different contexts. Ultimately, the lasting impacts of poor menstrual health may be better evaluated through prospective targeted studies. Despite these limitations, using MICS surveys also offered strengths, including providing nationally representative data from standardised methodologies to allow between-country comparisons of results. Our findings call attention to the burden of this understudied problem, are hypothesis-generating, and inform important avenues for future prospective research. Menstrual-related absenteeism is a common health and social issue for women in LMICs. Future work should prioritise identifying efficacious, culturally safe interventions for improving menstrual health in low-income countries, particularly in adolescent women.

## Data sharing

MICS data are the property of the UNICEF MICS program, and the data use agreement precludes redistribution. However, MICS study datasets are available upon request to the MICS programme for legitimate research purposes (available at: https://mics.unicef.org/surveys). Statistical code used for this analysis can be made available upon request to the corresponding author.

## Data Availability

All data produced in the present work are contained in the manuscript.

## Acknowledgements

This work was made possible through the Victorian State Government Operational Support Program and the Australian Government NHMRC IRIISS. S-RP is funded by the Australian National Health and Medical Research Council (NHMRC) Fellowship GNT2009047.

## References

1. Hennegan J, Winkler IT, Bobel C, et al. Menstrual health: a definition for policy, practice, and research. Sexual and Reproductive Health Matters 2021; 29(1): 31–8.

2. Hennegan J, Shannon AK, Rubli J, Schwab KJ, Melendez-Torres GJ. Women’s and girls’ experiences of menstruation in low- and middle-income countries: A systematic review and qualitative metasynthesis. PLoS Med 2019; 16(5): e1002803.

3. Hennegan J, OlaOlorun FM, Oumarou S, et al. School and work absenteeism due to menstruation in three West African countries: findings from PMA2020 surveys. Sexual and Reproductive Health Matters 2021; 29(1): 409–24.

4. Mahbub-Ul A, Stephen PL, Amal KH, et al. Menstrual hygiene management among Bangladeshi adolescent schoolgirls and risk factors affecting school absence: results from a cross-sectional survey. BMJ Open 2017; 7(7): e015508.

5. United Nations. Transforming Our World: The 2030 Agenda for Sustainable Development, 2015.

6. Progress on household drinking water, sanitation and hygiene 2000-2022: special focus on gender. New York: United Nations Children’s Fund (UNICEF) and World Health Organization (WHO), 2023.

7. Multiple Indicator Cluster Surveys. Surveys. https://mics.unicef.org/surveys (accessed 21 June 2023).

8. von Elm E, Altman DG, Egger M, Pocock SJ, Gøtzsche PC, Vandenbroucke JP. The Strengthening the Reporting of Observational Studies in Epidemiology (STROBE) statement: guidelines for reporting observational studies. The Lancet 2007; 370(9596): 1453–7.

9. Getahun MB, Nigatu AG. Knowledge of the Ovulatory Period and Associated Factors Among Reproductive Women in Ethiopia: A Population-Based Study Using the 2016 Ethiopian Demographic Health Survey. International Journal of Women’s Health 2020; 12(null): 701–7.

10. Prateek B, Saurabh S. A Cross Sectional Study of Knowledge and Practices about Reproductive Health among Female Adolescents in an Urban Slum of Mumbai. Journal of Family and Reproductive Health 2011; 5(4).

11. Ju H, Jones M, Mishra G. The Prevalence and Risk Factors of Dysmenorrhea. Epidemiologic Reviews 2013; 36(1): 104–13.

12. Tegegne TK, Sisay MM. Menstrual hygiene management and school absenteeism among female adolescent students in Northeast Ethiopia. BMC Public Health 2014; 14(1): 1118.

13. Zahradnik H-P, Hanjalic-Beck A, Groth K. Nonsteroidal anti-inflammatory drugs and hormonal contraceptives for pain relief from dysmenorrhea: a review. Contraception 2010; 81(3): 185–96.

14. Uhm S, Perriera L. Hormonal Contraception as Treatment for Heavy Menstrual Bleeding: A Systematic Review. Clinical Obstetrics and Gynecology 2014; 57(4).

15. Localio AR, Margolis DJ, Berlin JA. Relative risks and confidence intervals were easily computed indirectly from multivariable logistic regression. J Clin Epidemiol 2007; 60(9): 874–82.

16. Textor J, van der Zander B, Gilthorpe MS, Liśkiewicz M, Ellison GT. Robust causal inference using directed acyclic graphs: the R package ‘dagitty’. International Journal of Epidemiology 2017; 45(6): 1887–94.

17. Smith KB, Humphreys JS, Wilson MGA. Addressing the health disadvantage of rural populations: How does epidemiological evidence inform rural health policies and research? Australian Journal of Rural Health 2008; 16(2): 56–66.

18. Jahan F, Shuchi NS, Shoab AK, et al. Changes in the menstrual hygiene management facilities and usage among Bangladeshi school girls and its effect on school absenteeism from 2014 to 2018. Global Health Action 2024; 17(1): 2297512.

19. Sidik K, Jonkman JN. A simple confidence interval for meta-analysis. Stat Med 2002; 21(21): 3153–9.

20. Zou G. A modified poisson regression approach to prospective studies with binary data. Am J Epidemiol 2004; 159(7): 702–6.

21. Mansournia MA, Nazemipour M, Naimi AI, Collins GS, Campbell MJ. Reflection on modern methods: demystifying robust standard errors for epidemiologists. Int J Epidemiol 2021; 50(1): 346–51.

22. StataCorp. Stata 18 Survey Data Reference Manual. College Station, TX: Stata Press, 2023.

23. Lethaby A, Wise MR, Weterings MA, Bofill Rodriguez M, Brown J. Combined hormonal contraceptives for heavy menstrual bleeding. Cochrane Database Syst Rev 2019; 2(2): Cd000154.

24. Wong CL, Farquhar C, Roberts H, Proctor M. Oral contraceptive pill for primary dysmenorrhoea. Cochrane Database Syst Rev 2009; 2009(4): Cd002120.

25. Grant M, Lloyd C, Mensch B. Menstruation and School Absenteeism: Evidence from Rural Malawi. Comparative Education Review 2013; 57(2): 260–84.

26. Vashisht A, Pathak R, Agarwalla R, Patavegar BN, Panda M. School absenteeism during menstruation amongst adolescent girls in Delhi, India. J Family Community Med 2018; 25(3): 163–8.

27. Asumah MN, Adnani QES, Dzantor EK, et al. Menstruation-Related School Absenteeism: An Urban Centre Study in the Northern Region of Ghana. Women 2023; 3(4): 497–507.

28. Hennegan J, OlaOlorun FM, Oumarou S, et al. School and work absenteeism due to menstruation in three West African countries: findings from PMA2020 surveys. Sex Reprod Health Matters 2021; 29(1): 1915940.

29. Shah V, Nabwera H, Sonko B, et al. Effects of Menstrual Health and Hygiene on School Absenteeism and Drop-Out among Adolescent Girls in Rural Gambia. International Journal of Environmental Research and Public Health 2022; 19(6): 3337.

30. Miiro G, Rutakumwa R, Nakiyingi-Miiro J, et al. Menstrual health and school absenteeism among adolescent girls in Uganda (MENISCUS): a feasibility study. BMC Women’s Health 2018; 18(1): 4.

31. Crofts T, Fisher J. Menstrual hygiene in Ugandan schools: an investigation of low-cost sanitary pads. *Journal of Water*, Sanitation and Hygiene for Development 2012; 2(1): 50–8.

32. Kambala C, Chinangwa A, Chipeta E, Torondel B, Morse T. Acceptability of menstrual products interventions for menstrual hygiene management among women and girls in Malawi. Reproductive Health 2020; 17(1): 185.

33. Schmitt ML, Clatworthy D, Ogello T, Sommer M. Making the Case for a Female-Friendly Toilet. Water 2018; 10(9): 1193.

34. Sinharoy SS, Chery L, Patrick M, et al. Prevalence of heavy menstrual bleeding and associations with physical health and wellbeing in low-income and middle-income countries: a multinational cross-sectional study. The Lancet Global Health 2023; 11(11): e1775–e84.

