## Appendix for "Epidemiology of menstrual-related absenteeism in 44 low and middle-income countries"

### Supplementary Materials

**Table S1: GATHER checklist**

**Checklist of information that should be included in new reports of global health estimates**

| Item # | Checklist item | Reported on page # |
|---|---|---|
| **Objectives and funding** | | |
| 1 | Define the indicator(s), populations (including age, sex, and geographic entities), and time period(s) for which estimates were made. | 6-8<br>Table S2, S3 |
| 2 | List the funding sources for the work. | 3 |
| **Data Inputs** | | |
| *For all data inputs from multiple sources that are synthesized as part of the study:* | | |
| 3 | Describe how the data were identified and how the data were accessed. | 6 |
| 4 | Specify the inclusion and exclusion criteria. Identify all ad-hoc exclusions. | 6-8 |
| 5 | Provide information on all included data sources and their main characteristics. For each data source used, report reference information or contact name/institution, population represented, data collection method, year(s) of data collection, sex and age range, diagnostic criteria or measurement method, and sample size, as relevant. | 6<br>Table S3-S10 |
| 6 | Identify and describe any categories of input data that have potentially important biases (e.g., based on characteristics listed in item 5). | 14<br>Table S2 |
| *For data inputs that contribute to the analysis but were not synthesized as part of the study:* | | |
| 7 | Describe and give sources for any other data inputs. | N/A |
| *For all data inputs:* | | |
| 8 | Provide all data inputs in a file format from which data can be efficiently extracted (e.g., a spreadsheet rather than a PDF), including all relevant meta-data listed in item 5. For any data inputs that cannot be shared because of ethical or legal reasons, such as third-party ownership, provide a contact name or the name of the institution that retains the right to the data. | 14 |
| **Data analysis** | | |
| 9 | Provide a conceptual overview of the data analysis method. A diagram may be helpful. | Figure S1 |
| 10 | Provide a detailed description of all steps of the analysis, including mathematical formulae. This description should cover, as relevant, data cleaning, data pre-processing, data adjustments and weighting of data sources, and mathematical or statistical model(s). | 6-8<br>Table S2 |
| 11 | Describe how candidate models were evaluated and how the final model(s) were selected. | Figure S1 |
| 12 | Provide the results of an evaluation of model performance, if done, as well as the results of any relevant sensitivity analysis. | N/A |
| 13 | Describe methods for calculating uncertainty of the estimates. State which sources of uncertainty were, and were not, accounted for in the uncertainty analysis. | 6-8 |
| 14 | State how analytic or statistical source code used to generate estimates can be accessed. | 14 |
| **Results and Discussion** | | |
| 15 | Provide published estimates in a file format from which data can be efficiently extracted. | Table S4-S28 |
| 16 | Report a quantitative measure of the uncertainty of the estimates (e.g. uncertainty intervals). | Figures 4-5<br>Table S11-S28 |
| 17 | Interpret results in light of existing evidence. If updating a previous set of estimates, describe the reasons for changes in estimates. | 12-13 |
| 18 | Discuss limitations of the estimates. Include a discussion of any modelling assumptions or data limitations that affect interpretation of the estimates. | 14 |

*This checklist should be used in conjunction with the GATHER statement and Explanation and Elaboration document, found on gather-statement.org*

**Table S2: Overview of study variables and associated MICS6 questionnaire items.**

| Variable type | MICS Questionnaire | MICS variable | Description | Questionnaire Item |
|---|---|---|---|---|
| **Outcome** | Women | UN16* | Menstrual-related absenteeism | *Q*: Due to your last menstruation, were there any social activities, school or work days that you did not attend?<br>*A*: Yes/No/DK or Not sure or No such activity |
| **Independent** | Women | WAGE (categorical)<br>WB4 (continuous) | Woman's age | *Q*: How old are you?<br>*A*: Age (in completed years) |
| | | windex5 | Wealth quintiles | N/A<br>(Wealth quintiles represent the relative wealth within a country, and are derived from wealth scores which are calculated based on housing characteristics and both household and personal assets.)<br><br>*Note*: Available in both women's and household datasets. |
| | | UN17† | Availability of a private place to wash at home | *Q*: During your last menstrual period were you able to wash and change in privacy while at home?<br>*A*: Yes/No/DK |
| | | UN18† | Use of sanitary products (menstrual materials) | *Q*: Did you use any materials such as sanitary pads, tampons or cloth?<br>*A*: Yes/No/DK |
| | | CP2 | Current contraceptive use | *Q*: Couples use various ways or methods to delay or avoid getting pregnant. Are you currently doing something or using any method to delay or avoid getting pregnant?<br>*A*: Yes/No |
| | | Derived from CP4 | Hormonal contraception | N/A<br>Defines injectables, implants and pill as hormonal contraceptive methods.<br>Defines all other contraceptive methods as non-hormonal, except for IUD which is excluded from this analysis.<br>Comparator group includes all women and girls not using any contraception, and women and girls only using non-hormonal methods. |
| **Independent (excluded from main text‡)** | Household | HH6§ | Area | *Q:* Area:<br>*A:* Urban/Rural |
| | Women | UN19¶ | Use of reusable sanitary products | *Q:* Were the materials reusable?<br>*A:* Yes/No/DK |
| **Confounders** | Women | MSTATUS‖ | Woman's marital status | *Q:* Marital/Union status of woman<br>*A:* Currently married or in union/Formerly married or in union/Never married or in union |
| | | WB6** | Woman's education | *Q:* What is the highest level and grade or year of school you have attended?<br>*A:* Early childhood education/Primary/Lower secondary/Upper secondary/Higher |
| **Demographics** | Women | CP4†† | Contraceptive methods | *Q*: What are you doing to delay or avoid a pregnancy?<br>*A*: Female sterilisation/Male sterilisation/IUD/Injectables/Implants/Pill/Male condom/Female condom/Diaphragm/Foam or Jelly/Lactational amenorrhea method (LAM)/Periodic abstinence or rhythm/Withdrawal/Other (specify)<br><br>Note: Some countries measured additional methods. |
| | | WB14 | Women's literacy | *Q:* Now I would like you to read this sentence to me. *Respondent is shown a sentence card.*<br>*A:* Cannot read at all/Able to read only parts of sentence/Able to read whole sentence/No sentence in required language or braille<br><br>Probe: Can you read part of the sentence to me? |
| | | CM1 | Ever given birth | *Q:* Have you ever given birth?<br>*A*: Yes/No |
| | | CP1 | Current pregnancy status | *Q:* Are you pregnant now?<br>*A:* Yes, currently pregnant/No/DK or not sure |
| | | CP3 | Ever used contraception | *Q:* Have you ever done anything or used any method to delay or avoid getting pregnant?<br>*A:* Yes/No |
| | | AF11 | Woman's ability for self-care | *Q:* Do you have difficulty with self-care, such as washing all over or dressing?<br>*A:* No difficulty/Some difficulty/A lot of difficulty/Cannot care for self at all |
| | Household | HC1A‡‡ | Religion of household head | *Q:* What is the religion of (name of household head)?<br>*A:* Specific to each survey. |

* Central African Republic (2018-2019), Democratic Republic of the Congo (2017-2018), Togo (2017) and Tunisia (2018) surveys only measured absenteeism from work or social activities (not school). Zimbabwe (2019) survey measured menstrual-related absenteeism from social, cultural or religious, school or work activities. Nepal (2019) survey individually measured menstrual-related absenteeism from a range of activities - absenteeism from school or work, and absenteeism from social gatherings/meetings were combined to generate the outcome variable. The Nepal (2019) survey also did not offer an option for 'DK or Not sure or No such activity' when measuring menstrual-related absenteeism.

† Mongolia (2018) survey responses for the availability of a private place to wash during menstruation were recoded to match official reports. It was assumed that this variable was incorrectly coded, and therefore the 'yes' and 'no' responses were swapped to achieve the same results obtained in the following two publications:

Progress on household drinking water, sanitation and hygiene 2000-2022: special focus on gender. New York: United Nations Children's Fund (UNICEF) and World Health Organization (WHO); 2023.
National Statistical Office of Mongolia. Social Indicator Sample Survey-2018, Survey Findings Report. Ulaanbaatar, Mongolia; 2019.
† Costa Rica (2018) asked about use of sanitary pads, tampons, menstrual cup and cloths. Malawi (2019-2020) and Zimbabwe (2019) surveys asked about use of sanitary pads, cotton wool, menstrual cup, tampons or cloth. São Tomé and Príncipe (2019) asked about the use of pads, tampons, and pieces of towels in three different questions.
‡ Excluded from main text means results are only presented in supplementary materials.
§ Area response options in Lao PDR included urban, rural with road, and rural without road. These responses were recoded to urban, and rural (includes both rural with road and rural without road) for all analyses.
¶ UN19 (use of reusable sanitary materials) was only asked to women and girls who answered 'yes' to UN18 (use of sanitary materials).
|| MSTATUS is derived by MICS from other variables.
** Highest level of education answer options differed between surveys and were recoded for consistency into: Pre-school or none, primary, secondary, and higher education or vocational training.
†† CP4 (contraceptive methods) only asked to women and girls who answered 'yes' to CP2 (current use of contraception).
‡‡ Religion of household head was recoded for consistency into Christian/Islam or Muslim/ Animist/Buddhist/Hindu/Other/None/Don't know or no response.

**Table S3: Summary of included surveys.**

| Country | Survey year | Country income classification* | GDP per capita (2015 US$)† | Human Development Index‡ | Number of women and girls reporting menstruating in the previous 12 months (weighted) |
|---|---|---|---|---|---|
| **West and Central Africa** | | | | | |
| Central African Republic | 2018-2019 | Low | 379.9 | 0.404 | 7093 |
| Chad | 2019 | Low | 652.7 | 0.394 | 18211 |
| Democratic Republic of Congo | 2017-2018 | Low | 483.6 | 0.479 | 16987 |
| Gambia | 2018 | Low | 645.9 | 0.500 | 12177 |
| Ghana | 2017-2018 | Lower-Middle | 1863.6 | 0.632 | 12855 |
| Guinea-Bissau | 2018-2019 | Low | 627.3 | 0.483 | 10913 |
| Nigeria | 2021 | Lower-Middle | 2429.6 | 0.535 | 33195 |
| Sao Tome & Principe | 2019 | Lower-Middle | 1676.4 | 0.618 | 2858 |
| Sierra Leone | 2017 | Low | 611.1 | 0.477 | 13700 |
| Togo | 2017 | Low | 796.9 | 0.539 | 6080 |
| **Eastern and Southern Africa** | | | | | |
| Lesotho | 2018 | Lower-Middle | 1061.4 | 0.514 | 5648 |
| Madagascar | 2018 | Low | 470.4 | 0.501 | 14060 |
| Malawi | 2019-2020 | Low | 568.1 | 0.512 | 20498 |
| Zimbabwe | 2019 | Lower-Middle | 1343.0 | 0.593 | 8543 |
| **Middle East and North Africa** | | | | | |
| Algeria | 2018-2019 | Lower-Middle | 4170.6 | 0.745 | 33078 |
| Iraq | 2018 | Upper-Middle | 4711.0 | 0.686 | 19733 |
| State of Palestine (West Bank and Gaza) | 2019-2020 | Upper-Middle | 3150.5 | 0.715 | 6425 |
| Tunisia | 2018 | Lower-Middle | 4070.1 | 0.731 | 5668 |
| **South Asia** | | | | | |
| Afghanistan | 2022-2023 | Low | 363.7 | 0.478 | 40257 |
| Bangladesh | 2019 | Lower-Middle | 1558.0 | 0.661 | 58198 |
| Nepal | 2019 | Lower-Middle | 1061.5 | 0.602 | 13446 |
| Pakistan (Balochistan) | 2019-2020 | Lower-Middle | 1431.3 | 0.544 | 32395 |
| Pakistan (Khyber Pakhtunkhwa) | 2019 | Lower-Middle | 1452.9 | 0.544 | 37504 |
| Pakistan (Sindh) | 2018-2019 | Lower-Middle | 1446.7 | 0.544 | 27398 |
| Pakistan (Punjab) | 2017-2018 | Lower-Middle | 1409.0 | 0.544 | 68491 |
| **East Asia and the Pacific** | | | | | |
| Fiji | 2021 | Upper-Middle | 4442.9 | 0.730 | 4726 |
| Kiribati | 2018-2019 | Lower-Middle | 1432.7 | 0.624 | 3519 |
| Lao PDR | 2017 | Lower-Middle | 2358.4 | 0.607 | 22346 |
| Mongolia | 2018 | Lower-Middle | 4242.3 | 0.739 | 9489 |
| Samoa | 2019-2020 | Lower-Middle | 4322.45 | 0.707 | 3858 |
| Tonga | 2019 | Upper-Middle | 4628.4 | 0.745 | 2678 |
| Tuvalu | 2019-2020 | Upper-Middle | 4087.0 | 0.641 | 728 |
| Vietnam | 2020-2021 | Lower-Middle | 3380.6 | 0.703 | 10147 |
| **Europe and Central Asia** | | | | | |
| Kosovo | 2019-2020 | Upper-Middle | 4105.1 | — | 5020 |
| Kyrgyzstan | 2018 | Lower-Middle | 1197.6 | 0.692 | 5175 |
| Montenegro | 2018 | Upper-Middle | 7381.8 | 0.832 | 2156 |
| Republic of North Macedonia | 2018-2019 | Upper-Middle | 5285.5 | 0.770 | 3023 |
| Serbia | 2019 | Upper-Middle | 6567.9 | 0.802 | 3527 |
| Turkmenistan | 2019 | Upper-Middle | 7422.4 | 0.745 | 4946 |
| Uzbekistan | 2021-2022 | Lower-Middle | 3414.7 | 0.727 | 4360 |
| **Latin America and Caribbean** | | | | | |
| Costa Rica | 2018 | Upper-Middle | 12471.0 | 0.809 | 6795 |
| Cuba | 2019 | Upper-Middle | 8042.9 | 0.764 | 8243 |
| Dominican Republic | 2019 | Upper-Middle | 8205.1 | 0.767 | 20677 |
| Guyana | 2019-2020 | High | 7737.8 | 0.714 | 5411 |
| Honduras | 2019 | Lower-Middle | 2446.1 | 0.621 | 17572 |
| Suriname | 2018 | Upper-Middle | 8751.0 | 0.730 | 6441 |
| Turks & Caicos Islands | 2019-2020 | High | 21780.6 | 0.618 | 789 |

*World Bank country income classifications based on 2022 gross national income per capita (GNI)
†Average GDP per capita for survey year(s) except for Afghanistan, where 2021 data was used
‡Human development index (HDI) 2021. Low human development = HDI<0.550, Medium human development = 0.550≤HDI<0.700, High human development = HDI≥0.700.

**Table S4: Demographic characteristics of women and girls from West and Central Africa.**

| Demographics | CAR* | Chad | DRC† | Gambia | Ghana | Guinea-Bissau‡ | Nigeria§ | Sao Tome & Principe | Sierra Leone | Togo |
|---|---|---|---|---|---|---|---|---|---|---|
| **Age (years)** | | | | | | | | | | |
| 15-19 | 1539/7093 (21.7) | 4577/18211 (25.1) | 4486/16987 (26.4) | 2819/12177 (23.1) | 2761/12855 (21.5) | 2328/10913 (21.3) | 7700/33195 (23.2) | 676/2858 (23.6) | 2818/13700 (20.6) | 1320/6080 (21.7) |
| 20-24 | 1423/7093 (20.1) | 3332/18211 (18.3) | 3123/16987 (18.4) | 2494/12177 (20.5) | 2052/12855 (16.0) | 2238/10913 (20.5) | 5478/33195 (16.5) | 470/2858 (16.4) | 2661/13700 (19.4) | 994/6080 (16.3) |
| 25-29 | 1253/7093 (17.7) | 3202/18211 (17.6) | 2647/16987 (15.6) | 2011/12177 (16.5) | 1918/12855 (14.9) | 1911/10913 (17.5) | 4891/33195 (14.7) | 388/2858 (13.6) | 2395/13700 (17.5) | 1006/6080 (16.6) |
| 30-34 | 1067/7093 (15.0) | 2611/18211 (14.3) | 2437/16987 (14.3) | 1731/12177 (14.2) | 1884/12855 (14.7) | 1511/10913 (13.8) | 4467/33195 (13.5) | 413/2858 (14.5) | 1899/13700 (13.9) | 866/6080 (14.3) |
| 35-39 | 869/7093 (12.3) | 2201/18211 (12.1) | 2008/16987 (11.8) | 1505/12177 (12.4) | 1716/12855 (13.3) | 1373/10913 (12.6) | 4492/33195 (13.5) | 401/2858 (14.0) | 1790/13700 (13.1) | 813/6080 (13.4) |
| 40-44 | 565/7093 (8.0) | 1438/18211 (7.9) | 1498/16987 (8.8) | 982/12177 (8.1) | 1499/12855 (11.7) | 910/10913 (8.3) | 3660/33195 (11.0) | 316/2858 (11.0) | 1198/13700 (8.7) | 661/6080 (10.9) |
| 45-49 | 376/7093 (5.3) | 849/18211 (4.7) | 787/16987 (4.6) | 636/12177 (5.2) | 1025/12855 (8.0) | 643/10913 (5.9) | 2507/33195 (7.6) | 195/2858 (6.8) | 939/13700 (6.9) | 419/6080 (6.9) |
| **Wealth Quintile** | | | | | | | | | | |
| Q1 (Poorest) | 1327/7093 (18.7) | 3316/18211 (18.2) | 2741/16987 (16.1) | 2004/12177 (16.5) | 2045/12855 (15.9) | 1915/10913 (17.5) | 5357/33195 (16.1) | 513/2858 (17.9) | 2406/13700 (17.6) | 856/6080 (14.1) |
| Q2 | 1334/7093 (18.8) | 3410/18211 (18.7) | 2901/16987 (17.1) | 2081/12177 (17.1) | 2323/12855 (18.1) | 1989/10913 (18.2) | 5799/33195 (17.5) | 535/2858 (18.7) | 2418/13700 (17.6) | 1000/6080 (16.5) |
| Q3 | 1339/7093 (18.9) | 3573/18211 (19.6) | 2984/16987 (17.6) | 2364/12177 (19.4) | 2608/12855 (20.3) | 2076/10913 (19.0) | 6481/33195 (19.5) | 546/2858 (19.1) | 2508/13700 (18.3) | 1142/6080 (18.8) |
| Q4 | 1419/7093 (20.0) | 3688/18211 (20.3) | 3667/16987 (21.6) | 2651/12177 (21.8) | 2778/12855 (21.6) | 2299/10913 (21.1) | 7420/33195 (22.4) | 619/2858 (21.7) | 2812/13700 (20.5) | 1450/6080 (23.9) |
| Q5 (Richest) | 1674/7093 (23.6) | 4224/18211 (23.2) | 4695/16987 (27.6) | 3076/12177 (25.3) | 3101/12855 (24.1) | 2635/10913 (24.1) | 8139/33195 (24.5) | 645/2858 (22.6) | 3557/13700 (26.0) | 1630/6080 (26.8) |
| **Area** | | | | | | | | | | |
| Urban | 2802/7093 (39.5) | 3893/18211 (21.4) | 8959/16987 (52.7) | 8843/12177 (72.6) | 6636/12855 (51.6) | 4464/10913 (40.9) | 15993/33195 (48.2) | 1928/2858 (67.4) | 6922/13700 (50.5) | 3062/6080 (50.4) |
| Rural | 4290/7093 (60.5) | 14318/18211 (78.6) | 8028/16987 (47.3) | 3334/12177 (27.4) | 6219/12855 (48.4) | 6449/10913 (59.1) | 17202/33195 (51.8) | 930/2858 (32.6) | 6778/13700 (49.5) | 3018/6080 (49.6) |
| **Religion of household head¶** | | | | | | | | | | |
| Christian | 6422/7093 (90.5) | 7301/18211 (40.1) | 15144/16984 (89.2) | 455/12177 (3.7) | 9351/12855 (72.7) | 2600/10913 (23.8) | 17455/33195 (52.6) | 2292/2780 (82.5) | 3157/13700 (23.0) | 3344/6080 (55.0) |
| Islam/Muslim | 451/7093 (6.4) | 10088/18211 (55.4) | 285/16984 (1.7) | 11722/12177 (96.3) | 2435/12855 (18.9) | 6115/10913 (56.0) | 15530/33195 (46.8) | – | 10513/13700 (76.7) | 1108/6080 (18.2) |
| Animist | 96/7093 (1.3) | 188/18211 (1.0) | 437/16984 (2.6) | – | – | 944/10913 (8.7) | – | – | – | 1187/6080 (19.5) |
| Buddhist | – | – | – | – | – | – | – | – | – | – |
| Hindu | – | – | – | – | – | – | – | – | – | – |
| Other | 97/7093 (1.4) | 83/18211 (0.5) | 814/16984 (4.8) | – | 552/12855 (4.3) | 72/10913 (0.7) | 189/33195 (0.6) | 185/2780 (6.6) | 21/13700 (0.2) | 135/6080 (2.2) |
| None | 27/7093 (0.4) | 553/18211 (3.0) | 287/16984 (1.7) | – | 512/12855 (4.0) | 1182/10913 (10.8) | 19/33195 (0.1) | 303/2780 (10.9) | 1/13700 (0.0) | 305/6080 (5.0) |
| Don't know/No response | 0/7093 (0.0) | 0/18211 (0.0) | 18/16984 (0.1) | 1/12177 (0.0) | 5/12855 (0.0) | 0/10913 (0.0) | 2/33195 (0.0) | 0/2780 (0.0) | 9/13700 (0.1) | 0/6080 (0.0) |
| **Marital/union status** | | | | | | | | | | |
| Currently married/in union | 4750/7093 (67.0) | 12128/18211 (66.6) | 9086/16987 (53.5) | 7404/12177 (60.8) | 7053/12855 (54.9) | 6372/10913 (58.4) | 19248/33195 (58.0) | 1485/2858 (52.0) | 8254/13700 (60.2) | 3762/6080 (61.9) |

| | | | | | | | | | | |
|---|---|---|---|---|---|---|---|---|---|---|
| Formerly married/in union | 836/7093 (11.8) | 1501/18211 (8.2) | 1415/16987 (8.3) | 677/12177 (5.6) | 1216/12855 (9.5) | 631/10913 (5.8) | 1819/33195 (5.5) | 300/2858 (10.5) | 975/13700 (7.1) | 423/6080 (7.0) |
| Never married/in union | 1504/7093 (21.2) | 4566/18211 (25.1) | 6483/16987 (38.2) | 4095/12177 (33.6) | 4586/12855 (35.7) | 3907/10913 (35.8) | 12114/33195 (36.5) | 1066/2858 (37.3) | 4470/13700 (32.6) | 1893/6080 (31.1) |
| **Highest level of school attended‖** | | | | | | | | | | |
| Pre-school or none | 2408/7091 (34.0) | 11073/18210 (60.8) | 2102/16980 (12.4) | 4311/12176 (35.4) | 2225/12855 (17.3) | 4395/10913 (40.3) | 7835/33184 (23.6) | 66/2855 (2.3) | 6408/13700 (46.8) | 1500/6080 (24.7) |
| Primary | 2878/7091 (40.6) | 3911/18210 (21.5) | 4334/16980 (25.5) | 1853/12176 (15.2) | 2135/12855 (16.6) | 4797/10913 (44.0) | 4289/33184 (12.9) | 1071/2855 (37.5) | 1766/13700 (12.9) | 1951/6080 (32.1) |
| Secondary | 1642/7091 (23.2) | 2968/18210 (16.3) | 9279/16980 (54.6) | 4800/12176 (39.4) | 7717/12855 (60.0) | 1281/10913 (11.7) | 15863/33184 (47.8) | 1526/2855 (53.4) | 4867/13700 (35.5) | 2383/6080 (39.2) |
| Higher education or vocational training | 163/7091 (2.3) | 258/18210 (1.4) | 1265/16980 (7.5) | 1212/12176 (10.0) | 778/12855 (6.0) | 439/10913 (4.0) | 5194/33184 (15.7) | 191/2855 (6.7) | 659/13700 (4.8) | 246/6080 (4.0) |
| Don't know/no response | 1/7091 (0.0) | 0/18210 (0.0) | 0/16980 (0.0) | 0/12176 (0.0) | 0/12855 (0.0) | 1/10913 (0.0) | 4/33184 (0.0) | 2/2855 (0.1) | 0/13700 (0.0) | 0/6080 (0.0) |
| **Literacy** | | | | | | | | | | |
| Cannot read at all | 4772/5287 (90.3) | 13920/15038 (92.6) | 4894/6430 (76.1) | 5647/6165 (91.6) | 3936/4360 (90.3) | 4433/9203 (48.2) | 12104/28010 (43.2) | 304/1141 (26.6) | 7662/8174 (93.7) | 2535/3440 (73.7) |
| Able to read only parts of sentence | 304/5287 (5.7) | 994/15038 (6.6) | 1119/6430 (17.4) | 337/6165 (5.5) | 263/4360 (6.0) | 1343/9203 (14.6) | 5815/28010 (20.8) | 358/1141 (31.4) | 397/8174 (4.9) | 518/3440 (15.1) |
| Able to read whole sentence | 124/5287 (2.3) | 110/15038 (0.7) | 381/6430 (5.9) | 129/6165 (2.1) | 157/4360 (3.6) | 1839/9203 (20.0) | 9964/28010 (35.6) | 463/1141 (40.6) | 111/8174 (1.4) | 383/3440 (11.1) |
| No sentence in required language/ braille | 84/5287 (1.6) | 6/15038 (0.0) | 0/6430 (0.0) | 53/6165 (0.9) | 4/4360 (0.1) | 1587/9203 (17.2) | 75/28010 (0.3) | 11/1141 (0.9) | 0/8174 (0.0) | 0/3440 (0.0) |
| No response | 3/5287 (0.1) | 9/15038 (0.1) | 37/6430 (0.6) | 0/6165 (0.0) | 0/4360 (0.0) | 0/9203 (0.0) | 52/28010 (0.2) | 5/1141 (0.4) | 4/8174 (0.0) | 3/3440 (0.1) |
| **Difficulty with self-care** | | | | | | | | | | |
| No difficulty | 6126/6290 (97.4) | 14996/15478 (96.9) | 14249/14395 (99.0) | 10433/10491 (99.4) | 10944/11096 (98.6) | 9534/9630 (99.0) | – | 2394/2436 (98.3) | 11977/12169 (98.4) | 5162/5242 (98.5) |
| Some difficulty | 145/6290 (2.3) | 440/15478 (2.8) | 126/14395 (0.9) | 54/10491 (0.5) | 142/11096 (1.3) | 90/9630 (0.9) | – | 39/2436 (1.6) | 175/12169 (1.4) | 50/5242 (1.0) |
| A lot of difficulty | 17/6290 (0.3) | 30/15478 (0.2) | 20/14395 (0.1) | 0/10491 (0.0) | 7/11096 (0.1) | 6/9630 (0.1) | – | 0/2436 (0.0) | 14/12169 (0.1) | 21/5242 (0.4) |
| Cannot care for self at all | 0/6290 (0.0) | 7/15478 (0.0) | 0/14395 (0.0) | 1/10491 (0.0) | 2/11096 (0.0) | 0/9630 (0.0) | – | 0/2436 (0.0) | 1/12169 (0.0) | 7/5242 (0.1) |
| No response | 3/6290 (0.0) | 5/15478 (0.0) | 0/14395 (0.0) | 4/10491 (0.0) | 0/11096 (0.0) | 0/9630 (0.0) | – | 3/2436 (0.1) | 2/12169 (0.0) | 1/5242 (0.0) |
| **Ever given birth** | | | | | | | | | | |
| Yes | 5381/7093 (75.9) | 12936/18211 (71.0) | 10704/16987 (63.0) | 7506/12177 (61.6) | 8619/12855 (67.0) | 7635/10913 (70.0) | 20104/33195 (60.6) | 2006/2858 (70.2) | 9877/13700 (72.1) | 4103/6080 (67.5) |
| No | 1711/7093 (24.1) | 5275/18211 (29.0) | 6283/16987 (37.0) | 4671/12177 (38.4) | 4236/12855 (33.0) | 3278/10913 (30.0) | 13092/33195 (39.4) | 852/2858 (29.8) | 3824/13700 (27.9) | 1976/6080 (32.5) |
| **Currently pregnant** | | | | | | | | | | |
| Yes | 933/7093 (13.2) | 2792/18208 (15.3) | 2250/16987 (13.2) | 1075/12177 (8.8) | 889/12855 (6.9) | 924/10913 (8.5) | 3151/33195 (9.5) | 177/2858 (6.2) | 1215/13700 (8.9) | 476/6080 (7.8) |
| No | 6138/7093 (86.5) | 15321/18208 (84.1) | 14655/16987 (86.3) | 10971/12177 (90.1) | 11927/12855 (92.8) | 9890/10913 (90.6) | 29754/33195 (89.6) | 2649/2858 (92.7) | 12363/13700 (90.2) | 5579/6080 (91.8) |
| Don't know/No response | 21/7093 (0.3) | 95/18208 (0.5) | 83/16987 (0.5) | 132/12177 (1.1) | 40/12855 (0.3) | 99/10913 (0.9) | 291/33195 (0.9) | 31/2858 (1.1) | 122/13700 (0.9) | 25/6080 (0.4) |
| **Currently using contraception** | | | | | | | | | | |
| Yes | 1201/6160 (19.5) | 1347/15525 (8.7) | 4540/14738 (30.8) | 1416/11103 (12.8) | 2772/11966 (23.2) | 3378/10034 (33.7) | 6168/30044 (20.5) | 1024/2681 (38.2) | 3714/12485 (29.7) | 1346/5604 (24.0) |

| | | | | | | | | | | |
|---|---|---|---|---|---|---|---|---|---|---|
| No | 4952/6160 (80.4) | 14149/15525 (91.1) | 10156/14738 (68.9) | 9675/11103 (87.1) | 9194/11966 (76.8) | 6656/10034 (66.3) | 23785/30044 (79.2) | 1647/2681 (61.5) | 8750/12485 (70.1) | 4247/5604 (75.8) |
| No response | 7/6160 (0.1) | 28/15525 (0.2) | 42/14738 (0.3) | 12/11103 (0.1) | 0/11966 (0.0) | 0/10034 (0.0) | 92/30044 (0.3) | 9/2681 (0.3) | 21/12485 (0.2) | 11/5604 (0.2) |
| **Ever used contraception**** | | | | | | | | | | |
| Yes | 469/5891 (8.0) | 633/17013 (3.7) | 1408/12447 (11.3) | 1479/10761 (13.7) | 2065/10083 (20.5) | 703/7684 (9.1) | 2612/27028 (9.7) | 630/1834 (34.4) | 1818/9986 (18.2) | 838/4734 (17.7) |
| No | 5407/5891 (91.8) | 16376/17013 (96.3) | 11016/12447 (88.5) | 9262/10761 (86.1) | 8016/10083 (79.5) | 6981/7684 (90.9) | 24292/27028 (89.9) | 1201/1834 (65.5) | 8140/9986 (81.5) | 3894/4734 (82.2) |
| No response | 16/5891 (0.3) | 4/17013 (0.0) | 24/12447 (0.2) | 20/10761 (0.2) | 2/10083 (0.0) | 0/7684 (0.0) | 123/27028 (0.5) | 2/1834 (0.1) | 28/9986 (0.3) | 2/4734 (0.1) |
| **Current use of hormonal contraception††** | | | | | | | | | | |
| Yes | 686/6152 (11.1) | 819/15483 (5.3) | 1322/14634 (9.0) | 1298/11044 (11.8) | 2035/11866 (17.1) | 1830/9481 (19.3) | 3467/29656 (11.7) | 745/2628 (28.4) | 3512/12440 (28.2) | 767/5554 (13.8) |
| No | 5466/6152 (88.9) | 14660/15483 (94.7) | 13311/14634 (91.0) | 9747/11044 (88.2) | 9831/11866 (82.9) | 7644/9481 (80.6) | 26183/29656 (88.3) | 1879/2628 (71.5) | 8928/12440 (71.8) | 4786/5554 (86.2) |
| No response | 0/6152 (0.0) | 4/15483 (0.0) | 2/14634 (0.0) | 0/11044 (0.0) | 0/11866 (0.0) | 7/9481 (0.1) | 6/29656 (0.0) | 3/2628 (0.1) | 0/12440 (0.0) | 0/5554 (0.0) |
| **Current contraceptive method use** | | | | | | | | | | |
| Female sterilisation | 15/6153 (0.2) | 24/15497 (0.2) | 59/14695 (0.4) | 24/11091 (0.2) | 164/11966 (1.4) | 2/10034 (0.0) | 43/29952 (0.1) | 13/2671 (0.5) | 11/12464 (0.1) | 40/5593 (0.7) |
| Male sterilisation | 6/6153 (0.1) | 4/15497 (0.0) | 9/14695 (0.1) | 1/11091 (0.0) | 2/11966 (0.0) | 4/10034 (0.0) | 6/29952 (0.0) | 4/2671 (0.1) | 0/12464 (0.0) | 6/5593 (0.1) |
| IUD | 1/6153 (0.0) | 13/15497 (0.1) | 61/14695 (0.4) | 47/11091 (0.4) | 101/11966 (0.8) | 553/10034 (5.5) | 296/29952 (1.0) | 43/2671 (1.6) | 25/12464 (0.2) | 39/5593 (0.7) |
| Injectables | 182/6153 (3.0) | 543/15497 (3.5) | 346/14695 (2.4) | 658/11091 (5.9) | 892/11966 (7.5) | 104/10034 (1.0) | 1462/29952 (4.9) | 339/2671 (12.7) | 1824/12464 (14.6) | 375/5593 (6.7) |
| Implants | 144/6153 (2.3) | 223/15497 (1.4) | 381/14695 (2.6) | 444/11091 (4.0) | 516/11966 (4.3) | 1626/10034 (16.2) | 1183/29952 (4.0) | 53/2671 (2.0) | 705/12464 (5.7) | 253/5593 (4.5) |
| Pill | 388/6153 (6.3) | 102/15497 (0.7) | 646/14695 (4.4) | 196/11091 (1.8) | 644/11966 (5.4) | 100/10034 (1.0) | 979/29952 (3.3) | 361/2671 (13.5) | 1041/12464 (8.4) | 153/5593 (2.7) |
| Male condom | 178/6153 (2.9) | 56/15497 (0.4) | 981/14695 (6.7) | 14/11091 (0.1) | 109/11966 (0.9) | 383/10034 (3.8) | 1452/29952 (4.8) | 119/2671 (4.5) | 9/12464 (0.1) | 372/5593 (6.6) |
| Female condom | 13/6153 (0.2) | 15/15497 (0.1) | 100/14695 (0.7) | 0/11091 (0.0) | 8/11966 (0.1) | 41/10034 (0.4) | 180/29952 (0.6) | 13/2671 (0.5) | 5/12464 (0.0) | 18/5593 (0.3) |
| Diaphragm | 0/6153 (0.0) | 1/15497 (0.0) | 18/14695 (0.1) | 1/11091 (0.0) | 11/11966 (0.1) | 16/10034 (0.2) | 26/29952 (0.1) | 4/2671 (0.1) | 13/12464 (0.1) | 4/5593 (0.1) |
| Foam/Jelly | 0/6153 (0.0) | 4/15497 (0.0) | 0/14695 (0.0) | 0/11091 (0.0) | 0/11966 (0.0) | 1/10034 (0.0) | 5/29952 (0.0) | 3/2671 (0.1) | 4/12464 (0.0) | 1/5593 (0.0) |
| Lactational Amenorrhea Method (LAM) | 97/6153 (1.6) | 173/15497 (1.1) | 419/14695 (2.9) | 0/11091 (0.0) | 12/11966 (0.1) | 406/10034 (4.0) | 101/29952 (0.3) | 3/2671 (0.1) | 69/12464 (0.6) | 16/5593 (0.3) |
| Periodic abstinence/Rhythm | 235/6153 (3.8) | 321/15497 (2.1) | 1930/14695 (13.1) | 1/11091 (0.0) | 281/11966 (2.4) | 125/10034 (1.2) | 899/29952 (3.0) | 47/2671 (1.8) | 20/12464 (0.2) | 160/5593 (2.9) |
| Withdrawal | 43/6153 (0.7) | 21/15497 (0.1) | 483/14695 (3.3) | 4/11091 (0.0) | 59/11966 (0.5) | 2/10034 (0.0) | 894/29952 (3.0) | 5/2671 (0.2) | 2/12464 (0.0) | 25/5593 (0.5) |
| Other | 12/6153 (0.2) | 13/15497 (0.1) | 231/14695 (1.6) | 27/11091 (0.2) | 71/11966 (0.6) | 17/10034 (0.2) | 188/29952 (0.6) | 33/2671 (1.2) | 71/12464 (0.6) | 9/5593 (0.2) |
| No response | 0/6153 (0.0) | 4/15497 (0.0) | 2/14695 (0.0) | 0/11091 (0.0) | 0/11966 (0.0) | 6/10034 (0.1) | 6/29952 (0.0) | 3/2671 (0.1) | 0/12464 (0.0) | 0/5593 (0.0) |
| **Availability of a private place to wash during last menstrual period** | | | | | | | | | | |
| Yes | 6528/7093 (92.0) | 17007/18205 (93.4) | 15352/16987 (90.4) | 11701/12177 (96.1) | 12068/12855 (93.9) | – | 30815/33195 (92.8) | 2695/2858 (94.3) | 12728/13700 (92.9) | 5565/6080 (91.5) |
| No | 563/7093 (7.9) | 1188/18205 (6.5) | 1607/16987 (9.5) | 470/12177 (3.9) | 787/12855 (6.1) | – | 2371/33195 (7.1) | 162/2858 (5.7) | 958/13700 (7.0) | 514/6080 (8.5) |

| | | | | | | | | | | |
|---|---|---|---|---|---|---|---|---|---|---|
| Don't know/No response | 1/7093 (0.0) | 11/18205 (0.1) | 28/16987 (0.2) | 6/12177 (0.0) | 1/12855 (0.0) | – | 10/33195 (0.0) | 1/2858 (0.0) | 14/13700 (0.1) | 1/6080 (0.0) |
| **Use of menstrual materials** | | | | | | | | | | |
| Yes | 6745/7093 (95.1) | 17293/18204 (95.0) | 16066/16987 (94.6) | 11958/12177 (98.2) | 12583/12855 (97.9) | – | 32202/33195 (97.0) | 2845/2858 (99.6) | 13305/13700 (97.1) | 5859/6080 (96.4) |
| No | 346/7093 (4.9) | 901/18204 (4.9) | 890/16987 (5.2) | 216/12177 (1.8) | 270/12855 (2.1) | – | 981/33195 (3.0) | 13/2858 (0.4) | 389/13700 (2.8) | 214/6080 (3.5) |
| Don't know/No response | 2/7093 (0.0) | 11/18204 (0.1) | 32/16987 (0.2) | 3/12177 (0.0) | 3/12855 (0.0) | – | 13/33195 (0.0) | 0/2858 (0.0) | 7/13700 (0.0) | 7/6080 (0.1) |
| **Use of reusable menstrual materials‡‡** | | | | | | | | | | |
| Yes | 4388/7093 (61.9) | 14681/18201 (80.7) | 9444/16983 (55.6) | 7077/12174 (58.1) | 1614/12854 (12.6) | – | 13702/33187 (41.3) | 2371/2858 (83.0) | 9257/13698 (67.6) | 3479/6078 (57.2) |
| No | 2681/7093 (37.8) | 3516/18201 (19.3) | 7530/16983 (44.3) | 5081/12174 (41.7) | 11239/12854 (87.4) | – | 19436/33187 (58.6) | 485/2858 (17.0) | 4434/13698 (32.4) | 2596/6078 (42.7) |
| Don't know/No response | 23/7093 (0.3) | 3/18201 (0.0) | 9/16983 (0.1) | 16/12174 (0.1) | 1/12854 (0.0) | – | 49/33187 (0.1) | 2/2858 (0.1) | 8/13698 (0.1) | 2/6078 (0.0) |

Demographic factors are only described for sample women and girls without missing data for the outcome variable. Some variables (e.g. literacy) may have much lower sample sizes due to incomplete data collection. All values are weighted to reflect the survey sampling design. Data are n/N (%).

*CAR = Central African Republic.

†DRC = Democratic Republic of the Congo.

‡Guinea-Bissau results for use of menstrual materials, use of reusable menstrual materials, and availability of a private place to wash were excluded as the MICS report did not consider these estimates to reflect the reality of Guinea-Bissau due to an error that was detected when adapting the Computer-Assisted Personal Interviewing (CAPI) to tablets.

§Nigeria did not measure women's difficulty with self-care (sub-group of functional disability).

¶Religion of household head categories were unique to each survey and recoded into the representative groups; Christian, Islam/Muslim, Animist, Buddhist, Hindu, other, none, and don't know/no response. Not all surveys measured these religions. The symbol '–' in religion of household head denotes a lack of measurement for this religion.

||Highest level of education answer options differed between surveys and were recoded for consistency into: Pre-school or none, primary, secondary, and higher education or vocational training.

**Ever use of contraception was only collected for women and girls who reported no current use of contraception.

††Hormonal contraception derived from use of contraceptive methods. Users of hormonal contraception include women and girls using injectables, implants or the pill. Non-users of hormonal contraception include women and girls using only non-hormonal contraceptive methods, or no contraception. Women and girls using IUD were excluded from these groups.

‡‡Non-users of reusable menstrual materials include both women and girls who use no menstrual materials, and those who use only disposable menstrual materials.

**Table S5: Demographic characteristics of women and girls from Eastern and Southern Africa.**

| Demographics | Lesotho* | Madagascar | Malawi | Zimbabwe† |
|---|---|---|---|---|
| **Age (years)** | | | | |
| 15-19 | 1231/5648 (21.8) | 3616/14060 (25.7) | 5026/20498 (24.5) | 1818/8543 (21.3) |
| 20-24 | 1030/5648 (18.2) | 2625/14060 (18.7) | 3980/20498 (19.4) | 1396/8543 (16.3) |
| 25-29 | 883/5648 (15.6) | 2145/14060 (15.3) | 3119/20498 (15.2) | 1215/8543 (14.2) |
| 30-34 | 876/5648 (15.5) | 1688/14060 (12.0) | 2756/20498 (13.4) | 1281/8543 (15.0) |
| 35-39 | 686/5648 (12.1) | 1631/14060 (11.6) | 2576/20498 (12.6) | 1264/8543 (14.8) |
| 40-44 | 582/5648 (10.3) | 1456/14060 (10.4) | 1835/20498 (9.0) | 962/8543 (11.3) |
| 45-49 | 361/5648 (6.4) | 899/14060 (6.4) | 1208/20498 (5.9) | 608/8543 (7.1) |
| **Wealth quintile** | | | | |
| Q1 (Poorest) | 782/5648 (13.8) | 2267/14060 (16.1) | 3864/20498 (18.9) | 1338/8543 (15.7) |
| Q2 | 886/5648 (15.7) | 2409/14060 (17.1) | 3750/20498 (18.3) | 1416/8543 (16.6) |
| Q3 | 1109/5648 (19.6) | 2595/14060 (18.5) | 3821/20498 (18.6) | 1483/8543 (17.4) |
| Q4 | 1250/5648 (22.1) | 3069/14060 (21.8) | 4090/20498 (20.0) | 1972/8543 (23.1) |
| Q5 (Richest) | 1621/5648 (28.7) | 3721/14060 (26.5) | 4973/20498 (24.3) | 2333/8543 (27.3) |
| **Area** | | | | |
| Urban | 2738/5648 (48.5) | 3951/14060 (28.1) | 3989/20498 (19.5) | 3470/8543 (40.6) |
| Rural | 2909/5648 (51.5) | 10109/14060 (71.9) | 16509/20498 (80.5) | 5072/8543 (59.4) |
| **Religion of household head‡** | | | | |
| Christian | – | 9314/14060 (66.2) | 16698/20498 (81.5) | 7021/8543 (82.2) |
| Islam/Muslim | – | 204/14060 (1.4) | 2814/20498 (13.7) | 55/8543 (0.6) |
| Animist | – | 487/14060 (3.5) | – | – |
| Buddhist | – | – | 1/20498 (0.0) | – |
| Hindu | – | – | 12/20498 (0.1) | – |
| Other | – | 1242/14060 (8.8) | 439/20498 (2.1) | 308/8543 (3.6) |
| None | – | 2812/14060 (20.0) | 527/20498 (2.6) | 1158/8543 (13.6) |
| Don't know/No response | – | 2/14060 (0.0) | 8/20498 (0.0) | 0/8543 (0.0) |
| **Marital/union status** | | | | |
| Currently married/in union | 2917/5648 (51.6) | 8541/14060 (60.7) | 12319/20498 (60.1) | 5106/8543 (59.8) |
| Formerly married/in union | 671/5648 (11.9) | 1726/14060 (12.3) | 3069/20498 (15.0) | 1220/8543 (14.3) |
| Never married/in union | 2060/5648 (36.5) | 3792/14060 (27.0) | 5108/20498 (24.9) | 2216/8543 (25.9) |
| **Highest level of school attended§** | | | | |
| Pre-school or none | 33/5647 (0.6) | 2162/14055 (15.4) | 1638/20497 (8.0) | 66/8543 (0.8) |
| Primary | 1600/5647 (28.3) | 6108/14055 (43.5) | 12927/20497 (63.1) | 1943/8543 (22.7) |
| Secondary | 3204/5647 (56.7) | 5254/14055 (37.4) | 5364/20497 (26.2) | 5576/8543 (65.3) |
| Higher education or vocational training | 811/5647 (14.4) | 531/14055 (3.8) | 568/20497 (2.8) | 957/8543 (11.2) |
| Don't know/no response | 0/5647 (0.0) | 0/14055 (0.0) | 0/20497 (0.0) | 0/8543 (0.0) |
| **Literacy** | | | | |
| Cannot read at all | 180/1633 (11.0) | 3328/8274 (40.2) | 5546/14632 (37.9) | 467/2002 (23.3) |
| Able to read only parts of sentence | 233/1633 (14.2) | 1890/8274 (22.8) | 1633/14632 (11.2) | 156/2002 (7.8) |
| Able to read whole sentence | 1215/1633 (74.4) | 2980/8274 (36.0) | 7386/14632 (50.5) | 1368/2002 (68.4) |
| No sentence in required language/braille | 3/1633 (0.2) | 74/8274 (0.9) | 45/14632 (0.3) | 0/2002 (0.0) |
| No response | 2/1633 (0.1) | 2/8274 (0.0) | 22/14632 (0.2) | 10/2002 (0.5) |
| **Difficulty with self-care** | | | | |
| No difficulty | 4902/4941 (99.2) | 11725/12001 (97.7) | 17229/17521 (98.3) | 7355/7427 (99.0) |
| Some difficulty | 29/4941 (0.6) | 253/12001 (2.1) | 274/17521 (1.6) | 66/7427 (0.9) |
| A lot of difficulty | 8/4941 (0.2) | 20/12001 (0.2) | 17/17521 (0.1) | 4/7427 (0.1) |
| Cannot care for self at all | 2/4941 (0.0) | 0/12001 (0.0) | 1/17521 (0.0) | 0/7427 (0.0) |
| No response | 0/4941 (0.0) | 2/12001 (0.0) | 0/17521 (0.0) | 2/7427 (0.0) |
| **Ever given birth** | | | | |
| Yes | 3701/5648 (65.5) | 10080/14060 (71.7) | 15104/20498 (73.7) | 6139/8543 (71.9) |
| No | 1947/5648 (34.5) | 3980/14060 (28.3) | 5394/20498 (26.3) | 2404/8543 (28.1) |
| **Currently pregnant** | | | | |
| Yes | 203/5648 (3.6) | 1163/14060 (8.3) | 1539/20498 (7.5) | – |
| No | 5402/5648 (95.7) | 12818/14060 (91.2) | 18867/20498 (92.0) | – |
| Don't know/No response | 42/5648 (0.7) | 79/14060 (0.6) | 92/20498 (0.5) | – |
| **Currently using contraception** | | | | |
| Yes | 2879/5444 (52.9) | 5208/12897 (40.4) | 9766/18959 (51.5) | – |
| No | 2560/5444 (47.0) | 7685/12897 (59.6) | 9185/18959 (48.4) | – |
| No response | 5/5444 (0.1) | 4/12897 (0.0) | 8/18959 (0.0) | – |
| **Ever used contraception¶** | | | | |
| Yes | 989/2768 (35.7) | 2444/8852 (27.6) | 3922/10732 (36.5) | – |
| No | 1775/2768 (64.1) | 6406/8852 (72.4) | 6770/10732 (63.1) | – |
| No response | 5/2768 (0.2) | 1/8852 (0.0) | 40/10732 (0.4) | – |
| **Current use of hormonal contraception‖** | | | | |
| Yes | 1852/5351 (34.6) | 4385/12770 (34.3) | 7819/18783 (41.6) | – |
| No | 3498/5351 (65.4) | 8386/12770 (65.7) | 10964/18783 (58.4) | – |
| No response | 0/5351 (0.0) | 0/12770 (0.0) | 0/18783 (0.0) | – |
| **Current contraceptive method use** | | | | |

| | | | | |
|---|---|---|---|---|
| Female sterilisation | 74/5439 (1.4) | 55/12893 (0.4) | 1330/18952 (7.0) | – |
| Male sterilisation | 18/5439 (0.3) | 3/12893 (0.0) | 10/18952 (0.1) | – |
| IUD | 88/5439 (1.6) | 123/12893 (1.0) | 168/18952 (0.9) | – |
| Injectables | 937/5439 (17.2) | 3102/12893 (24.1) | 4884/18952 (25.8) | – |
| Implants | 240/5439 (4.4) | 701/12893 (5.4) | 2667/18952 (14.1) | – |
| Pill | 694/5439 (12.8) | 613/12893 (4.8) | 459/18952 (2.4) | – |
| Male condom | 1016/5439 (18.7) | 57/12893 (0.4) | 274/18952 (1.4) | – |
| Female condom | 82/5439 (1.5) | 3/12893 (0.0) | 58/18952 (0.3) | – |
| Diaphragm | 15/5439 (0.3) | 7/12893 (0.1) | 24/18952 (0.1) | – |
| Foam/Jelly | 3/5439 (0.1) | 2/12893 (0.0) | 1/18952 (0.0) | – |
| Lactational Amenorrhea Method (LAM) | – | 33/12893 (0.3) | 30/18952 (0.2) | – |
| Periodic abstinence/Rhythm | 21/5439 (0.4) | 536/12893 (4.2) | 41/18952 (0.2) | – |
| Withdrawal | 18/5439 (0.3) | 67/12893 (0.5) | 38/18952 (0.2) | – |
| Other | 29/5439 (0.5) | 26/12893 (0.2) | 63/18952 (0.3) | – |
| No response | 0/5439 (0.0) | 0/12893 (0.0) | 0/18952 (0.0) | – |
| **Availability of a private place to wash during last menstrual period** | | | | |
| Yes | 5349/5648 (94.7) | 12765/14060 (90.8) | 18968/20498 (92.5) | 8253/8543 (96.6) |
| No | 296/5648 (5.2) | 1291/14060 (9.2) | 1528/20498 (7.5) | 290/8543 (3.4) |
| Don't know/No response | 3/5648 (0.1) | 4/14060 (0.0) | 2/20498 (0.0) | 0/8543 (0.0) |
| **Use of menstrual materials** | | | | |
| Yes | 5539/5648 (98.1) | 13186/14060 (93.8) | 19954/20498 (97.3) | 8355/8543 (97.8) |
| No | 108/5648 (1.9) | 873/14060 (6.2) | 543/20498 (2.6) | 185/8543 (2.2) |
| Don't know/No response | 1/5648 (0.0) | 1/14060 (0.0) | 2/20498 (0.0) | 2/8543 (0.0) |
| **Use of reusable menstrual materials**** | | | | |
| Yes | 427/5647 (7.6) | 10262/14059 (73.0) | 14039/20497 (68.5) | 1845/8541 (21.6) |
| No | 5214/5647 (92.3) | 3792/14059 (27.0) | 6457/20497 (31.5) | 6696/8541 (78.4) |
| Don't know/No response | 6/5647 (0.1) | 5/14059 (0.0) | 1/20497 (0.0) | 0/8541 (0.0) |

Demographic factors are only described for sample women and girls without missing data for the outcome variable. Some variables (e.g. literacy) may have much lower sample sizes due to incomplete data collection. All values are weighted to reflect the survey sampling design. Data are n/N (%).

*Lesotho: Variables or categories not measured include: religion of household head and use of the contraceptive method Lactational Amenorrhea Method (LAM).

†Zimbabwe: Variables or categories not measured include: pregnancy status, ever and current use of contraception and use of contraceptive methods.

‡Religion of household head categories were unique to each survey and recoded into the representative groups; Christian, Islam/Muslim, Animist, Buddhist, Hindu, other, none, and don't know/no response. Not all surveys measured these religions. The symbol '–' in religion of household head denotes a lack of measurement for this religion. Note that Lesotho did not measure any religion of the household head.

§Highest level of education answer options differed between surveys and were recoded for consistency into: Pre-school or none, primary, secondary, and higher education or vocational training.

¶Ever use of contraception was only collected for women and girls who reported no current use of contraception.

||Hormonal contraception derived from use of contraceptive methods. Users of hormonal contraception include women and girls using injectables, implants or the pill. Non-users of hormonal contraception include women and girls using only non-hormonal contraceptive methods, or no contraception. Women and girls using IUD were excluded from these groups.

**Non-users of reusable menstrual materials include both women and girls who use no menstrual materials, and those who use only disposable menstrual materials.

**Table S6: Demographic characteristics of women and girls from Middle East and North Africa.**

| Demographics | Algeria | Iraq | State of Palestine | Tunisia |
|---|---|---|---|---|
| **Age (years)** | | | | |
| 15-19 | 4722/33078 (14.3) | 1205/19733 (6.1) | 168/6425 (2.6) | 9/5668 (0.2) |
| 20-24 | 5092/33078 (15.4) | 2881/19733 (14.6) | 989/6425 (15.4) | 202/5668 (3.6) |
| 25-29 | 5323/33078 (16.1) | 3580/19733 (18.1) | 1350/6425 (21.0) | 623/5668 (11.0) |
| 30-34 | 5203/33078 (15.7) | 3597/19733 (18.2) | 1200/6425 (18.7) | 1240/5668 (21.9) |
| 35-39 | 5018/33078 (15.2) | 3486/19733 (17.7) | 1086/6425 (16.9) | 1319/5668 (23.3) |
| 40-44 | 4495/33078 (13.6) | 2864/19733 (14.5) | 928/6425 (14.4) | 1308/5668 (23.1) |
| 45-49 | 3224/33078 (9.7) | 2120/19733 (10.7) | 704/6425 (11.0) | 968/5668 (17.1) |
| **Wealth quintile** | | | | |
| Q1 (Poorest) | 6412/33078 (19.4) | 3711/19733 (18.8) | 1259/6425 (19.6) | 943/5668 (16.6) |
| Q2 | 6439/33078 (19.5) | 4056/19733 (20.6) | 1208/6425 (18.8) | 1151/5668 (20.3) |
| Q3 | 6604/33078 (20.0) | 3969/19733 (20.1) | 1228/6425 (19.1) | 1139/5668 (20.1) |
| Q4 | 6679/33078 (20.2) | 4043/19733 (20.5) | 1392/6425 (21.7) | 1225/5668 (21.6) |
| Q5 (Richest) | 6945/33078 (21.0) | 3955/19733 (20.0) | 1337/6425 (20.8) | 1210/5668 (21.3) |
| **Area** | | | | |
| Urban | 21058/33078 (63.7) | 13961/19733 (70.7) | 4974/6425 (77.4) | 3937/5668 (69.5) |
| Rural | 12020/33078 (36.3) | 5773/19733 (29.3) | 948/6425 (14.7) | 1731/5668 (30.5) |
| **Religion of household head*** | | | | |
| Christian | – | 67/19733 (0.3) | – | – |
| Islam/Muslim | – | 19645/19733 (99.6) | – | – |
| Animist | – | – | – | – |
| Buddhist | – | – | – | – |
| Hindu | – | – | – | – |
| Other | – | 22/19733 (0.1) | – | – |
| None | – | – | – | – |
| Don’t know/No response | – | 0/19733 (0.0) | – | – |
| **Marital/union status** | | | | |
| Currently married/in union | 17637/33078 (53.3) | 13961/19733 (70.7) | 6425/6425 (100.0) | 5390/5668 (95.1) |
| Formerly married/in union | 1091/33078 (3.3) | 5773/19733 (29.3) | 0/6425 (0.0) | 278/5668 (4.9) |
| Never married/in union | 14347/33078 (43.4) | 0/19733 (0.0) | 0/6425 (0.0) | 0/5668 (0.0) |
| **Highest level of school attended†** | | | | |
| Pre-school or none | 3270/33075 (9.9) | 3179/19733 (16.1) | 22/6424 (0.3) | 565/5660 (10.0) |
| Primary | 3851/33075 (11.6) | 8615/19733 (43.7) | 1622/6424 (25.2) | 1614/5660 (28.5) |
| Secondary | 17869/33075 (54.0) | 5338/19733 (27.1) | 2174/6424 (33.8) | 2195/5660 (38.8) |
| Higher education or vocational training | 8080/33075 (24.4) | 2601/19733 (13.2) | 2606/6424 (40.6) | 1285/5660 (22.7) |
| Don’t know/no response | 5/33075 (0.0) | 0/19733 (0.0) | 0/6424 (0.0) | 0/5660 (0.0) |
| **Literacy** | | | | |
| Cannot read at all | 3454/7232 (47.8) | 4538/11794 (38.5) | 41/75 (55.6) | 813/2188 (37.2) |
| Able to read only parts of sentence | 1522/7232 (21.0) | 2594/11794 (22.0) | 22/75 (28.9) | 351/2188 (16.0) |
| Able to read whole sentence | 2238/7232 (30.9) | 4632/11794 (39.3) | 11/75 (14.4) | 1018/2188 (46.5) |
| No sentence in required language/braille | 4/7232 (0.1) | 28/11794 (0.2) | 0/75 (0.0) | 3/2188 (0.1) |
| No response | 15/7232 (0.2) | 3/11794 (0.0) | 1/75 (1.1) | 3/2188 (0.1) |
| **Difficulty with self-care** | | | | |
| No difficulty | 29620/30307 (97.7) | 18896/19285 (98.0) | 6354/6394 (99.4) | 5407/5668 (95.4) |
| Some difficulty | 641/30307 (2.1) | 317/19285 (1.6) | 34/6394 (0.5) | 232/5668 (4.1) |
| A lot of difficulty | 38/30307 (0.1) | 69/19285 (0.4) | 4/6394 (0.1) | 26/5668 (0.5) |
| Cannot care for self at all | 8/30307 (0.0) | 3/19285 (0.0) | 0/6394 (0.0) | 2/5668 (0.0) |
| No response | 0/30307 (0.0) | 0/19285 (0.0) | 2/6394 (0.0) | 1/5668 (0.0) |
| **Ever given birth** | | | | |
| Yes | 16438/18739 (87.7) | 17701/19733 (89.7) | 5880/6425 (91.5) | 5066/5668 (89.4) |
| No | 2301/18739 (12.3) | 2033/19733 (10.3) | 545/6425 (8.5) | 602/5668 (10.6) |
| **Currently pregnant** | | | | |
| Yes | 2080/17652 (11.8) | 2008/19733 (10.2) | 718/6425 (11.2) | 348/5668 (6.1) |
| No | 15492/17652 (87.8) | 17627/19733 (89.3) | 5669/6425 (88.2) | 5300/5668 (93.5) |
| Don’t know/No response | 81/17652 (0.5) | 97/19733 (0.5) | 38/6425 (0.6) | 20/5668 (0.3) |
| **Currently using contraception** | | | | |
| Yes | 9907/15679 (63.2) | 10143/17725 (57.2) | 3788/5707 (66.4) | 2865/5320 (53.9) |
| No | 5758/15679 (36.7) | 7550/17725 (42.6) | 1914/5707 (33.5) | 2449/5320 (46.0) |
| No response | 14/15679 (0.1) | 32/17725 (0.2) | 5/5707 (0.1) | 6/5320 (0.1) |
| **Ever used contraception‡** | | | | |
| Yes | 3442/8120 (42.4) | 2657/9591 (27.7) | 822/2637 (31.2) | 1060/2803 (37.8) |
| No | 4662/8120 (57.4) | 6906/9591 (72.0) | 1810/2637 (68.6) | 1739/2803 (62.0) |
| No response | 17/8120 (0.2) | 28/9591 (0.3) | 4/2637 (0.2) | 4/2803 (0.1) |
| **Current use of hormonal contraception§** | | | | |
| Yes | 7335/15205 (48.2) | 3873/15983 (24.2) | 510/3916 (13.0) | 1618/3715 (43.6) |
| No | 7857/15205 (51.7) | 12109/15983 (75.8) | 3404/3916 (86.9) | 2096/3715 (56.4) |
| No response | 13/15205 (0.1) | 2/15983 (0.0) | 1/3916 (0.0) | 0/3715 (0.0) |
| **Current contraceptive method use** | | | | |
| Female sterilisation | 55/15665 (0.4) | 552/17693 (3.1) | 117/5702 (2.0) | 76/5314 (1.4) |

| | | | | |
|---|---|---|---|---|
| Male sterilisation | 14/15665 (0.1) | 18/17693 (0.1) | 22/5702 (0.4) | 7/5314 (0.1) |
| IUD | 460/15665 (2.9) | 1710/17693 (9.7) | 1786/5702 (31.3) | 1599/5314 (30.1) |
| Injectables | 8/15665 (0.0) | 714/17693 (4.0) | 54/5702 (0.9) | 82/5314 (1.5) |
| Implants | 54/15665 (0.3) | 39/17693 (0.2) | 11/5702 (0.2) | 59/5314 (1.1) |
| Pill | 7305/15665 (46.6) | 3170/17693 (17.9) | 479/5702 (8.4) | 1637/5314 (30.8) |
| Male condom | 398/15665 (2.5) | 660/17693 (3.7) | 371/5702 (6.5) | 71/5314 (1.3) |
| Female condom | 10/15665 (0.1) | 18/17693 (0.1) | 11/5702 (0.2) | 10/5314 (0.2) |
| Diaphragm | 2/15665 (0.0) | 19/17693 (0.1) | 5/5702 (0.1) | 0/5314 (0.0) |
| Foam/Jelly | 2/15665 (0.0) | 11/17693 (0.1) | 1/5702 (0.0) | 5/5314 (0.1) |
| Lactational Amenorrhea Method (LAM) | 122/15665 (0.8) | 102/17693 (0.6) | 52/5702 (0.9) | 15/5314 (0.3) |
| Periodic abstinence/Rhythm | 1058/15665 (6.8) | 322/17693 (1.8) | 303/5702 (5.3) | 399/5314 (7.5) |
| Withdrawal | 787/15665 (5.0) | 3282/17693 (18.5) | 726/5702 (12.7) | 12/5314 (0.2) |
| Other | 35/15665 (0.2) | 14/17693 (0.1) | 18/5702 (0.3) | 30/5314 (0.6) |
| No response | 13/15665 (0.1) | 2/17693 (0.0) | 1/5702 (0.0) | 0/5314 (0.0) |
| **Availability of a private place to wash during last menstrual period** | | | | |
| Yes | 29824/33074 (90.2) | 14889/16799 (88.6) | 5170/6425 (80.5) | 3185/5668 (56.2) |
| No | 3209/33074 (9.7) | 1907/16799 (11.4) | 1247/6425 (19.4) | 2475/5668 (43.7) |
| Don't know/No response | 41/33074 (0.1) | 3/16799 (0.0) | 8/6425 (0.1) | 8/5668 (0.1) |
| **Use of menstrual materials** | | | | |
| Yes | 31262/33074 (94.5) | 16110/16799 (95.9) | 6219/6425 (96.8) | 5440/5668 (96.0) |
| No | 1764/33074 (5.3) | 686/16799 (4.1) | 205/6425 (3.2) | 226/5668 (4.0) |
| Don't know/No response | 48/33074 (0.1) | 3/16799 (0.0) | 1/6425 (0.0) | 3/5668 (0.1) |
| **Use of reusable menstrual materials¶** | | | | |
| Yes | 1665/33055 (5.0) | 1871/16796 (11.1) | 137/6424 (2.1) | 207/5667 (3.7) |
| No | 31320/33055 (94.8) | 14923/16796 (88.9) | 6253/6424 (97.3) | 5457/5667 (96.3) |
| Don't know/No response | 71/33055 (0.2) | 2/16796 (0.0) | 34/6424 (0.5) | 3/5667 (0.1) |

Demographic factors are only described for sample women and girls without missing data for the outcome variable. Some variables (e.g. literacy) may have much lower sample sizes due to incomplete data collection. All values are weighted to reflect the survey sampling design. Data are n/N (%).

*Religion of household head categories were unique to each survey and recoded into the representative groups; Christian, Islam/Muslim, Animist, Buddhist, Hindu, other, none, and don't know/no response. Not all surveys measured these religions. The symbol '–' in religion of household head denotes a lack of measurement for this religion. Note that Algeria, State of Palestine, and Tunisia did not measure any religion of the household head.

†Highest level of education answer options differed between surveys and were recoded for consistency into: Pre-school or none, primary, secondary, and higher education or vocational training.

‡Ever use of contraception was only collected for women and girls who reported no current use of contraception.

§Hormonal contraception derived from use of contraceptive methods. Users of hormonal contraception include women and girls using injectables, implants or the pill. Non-users of hormonal contraception include women and girls using only non-hormonal contraceptive methods, or no contraception. Women and girls using IUD were excluded from these groups.

¶Non-users of reusable menstrual materials include both women and girls who use no menstrual materials, and those who use only disposable menstrual materials.

**Table S7: Demographic characteristics of women and girls from South Asia.**

| Demographics | Afghanistan* | Bangladesh | Nepal | Pakistan (Balochistan) | Pakistan (Khyber Pakhtunkhwa) | Pakistan (Sindh) | Pakistan (Punjab) |
|---|---|---|---|---|---|---|---|
| **Age (years)** | | | | | | | |
| 15-19 | 11568/40257 (28.7) | 11654/58198 (20.0) | 2582/13446 (19.2) | 7094/32395 (21.9) | 8521/37504 (22.7) | 6059/27398 (22.1) | 14261/68491 (20.8) |
| 20-24 | 8713/40257 (21.6) | 9740/58198 (16.7) | 2332/13446 (17.3) | 6233/32395 (19.2) | 7009/37504 (18.7) | 5022/27398 (18.3) | 13032/68491 (19.0) |
| 25-29 | 6920/40257 (17.2) | 9371/58198 (16.1) | 2183/13446 (16.2) | 6264/32395 (19.3) | 6819/37504 (18.2) | 4709/27398 (17.2) | 11810/68491 (17.2) |
| 30-34 | 4488/40257 (11.1) | 9535/58198 (16.4) | 1974/13446 (14.7) | 4764/32395 (14.7) | 5424/37504 (14.5) | 4227/27398 (15.4) | 9777/68491 (14.3) |
| 35-39 | 3852/40257 (9.6) | 8495/58198 (14.6) | 1847/13446 (13.7) | 3647/32395 (11.3) | 4581/37504 (12.2) | 3507/27398 (12.8) | 9043/68491 (13.2) |
| 40-44 | 2802/40257 (7.0) | 5803/58198 (10.0) | 1509/13446 (11.2) | 2403/32395 (7.4) | 3098/37504 (8.3) | 2338/27398 (8.5) | 6317/68491 (9.2) |
| 45-49 | 1913/40257 (4.8) | 3601/58198 (6.2) | 1018/13446 (7.6) | 1990/32395 (6.1) | 2051/37504 (5.5) | 1536/27398 (5.6) | 4250/68491 (6.2) |
| **Wealth quintile** | | | | | | | |
| Q1 (Poorest) | 6858/40257 (17.0) | 10098/58198 (17.4) | 2294/13446 (17.1) | 5927/32395 (18.3) | 6830/37504 (18.2) | 4684/27398 (17.1) | 11403/68491 (16.6) |
| Q2 | 7427/40257 (18.4) | 10953/58198 (18.8) | 2525/13446 (18.8) | 6440/32395 (19.9) | 7160/37504 (19.1) | 5037/27398 (18.4) | 13168/68491 (19.2) |
| Q3 | 7932/40257 (19.7) | 11727/58198 (20.1) | 2629/13446 (19.6) | 6603/32395 (20.4) | 7537/37504 (20.1) | 5606/27398 (20.5) | 14005/68491 (20.4) |
| Q4 | 8690/40257 (21.6) | 12377/58198 (21.3) | 2883/13446 (21.4) | 6413/32395 (19.8) | 7832/37504 (20.9) | 5986/27398 (21.8) | 14632/68491 (21.4) |
| Q5 (Richest) | 9349/40257 (23.2) | 13044/58198 (22.4) | 3114/13446 (23.2) | 7011/32395 (21.6) | 8144/37504 (21.7) | 6086/27398 (22.2) | 15282/68491 (22.3) |
| **Area** | | | | | | | |
| Urban | 11249/40257 (27.9) | 13742/58198 (23.6) | 9393/13446 (69.9) | 8638/32395 (26.7) | 6352/37504 (16.9) | 15102/27398 (55.1) | 26508/68491 (38.7) |
| Rural | 29007/40257 (72.1) | 44456/58198 (76.4) | 4052/13446 (30.1) | 23756/32395 (73.3) | 31152/37504 (83.1) | 12296/27398 (44.9) | 41983/68491 (61.3) |
| **Religion of household head†** | | | | | | | |
| Christian | – | 295/58198 (0.5) | 289/13446 (2.1) | – | – | – | – |
| Islam/Muslim | – | 52576/58198 (90.3) | 466/13446 (3.5) | – | – | – | – |
| Animist | – | – | – | – | – | – | – |
| Buddhist | – | 482/58198 (0.8) | 912/13446 (6.8) | – | – | – | – |
| Hindu | – | 4844/58198 (8.3) | 11470/13446 (85.3) | – | – | – | – |
| Other | – | – | 307/13446 (2.3) | – | – | – | – |
| None | – | – | 2/13446 (0.0) | – | – | – | – |
| Don't know/No response | – | 0/58198 (0.0) | 0/13446 (0.0) | – | – | – | – |
| **Marital/union status** | | | | | | | |
| Currently married/in union | 24319/40257 (60.4) | 45352/58198 (77.9) | 9911/13446 (73.7) | 20119/32395 (62.1) | 24541/37504 (65.4) | 17556/27398 (64.1) | 42122/68491 (61.5) |
| Formerly married/in union | 767/40257 (1.9) | 2245/58198 (3.9) | 268/13446 (2.0) | 481/32395 (1.5) | 507/37504 (1.4) | 636/27398 (2.3) | 2063/68491 (3.0) |
| Never married/in union | 15169/40257 (37.7) | 10601/58198 (18.2) | 3266/13446 (24.3) | 11788/32395 (36.4) | 12456/37504 (33.2) | 9204/27398 (33.6) | 24305/68491 (35.5) |
| **Highest level of school attended‡** | | | | | | | |
| Pre-school or none | 26673/40135 (66.5) | 8208/58195 (14.1) | 3322/13446 (24.7) | 24291/32313 (75.2) | 21529/37469 (57.5) | 14132/27387 (51.6) | 22121/68483 (32.3) |
| Primary | 4691/40135 (11.7) | 12866/58195 (22.1) | 1797/13446 (13.4) | 2146/32313 (6.6) | 4638/37469 (12.4) | 3286/27387 (12.0) | 12599/68483 (18.4) |
| Secondary | 7065/40135 (17.6) | 32309/58195 (55.5) | 7059/13446 (52.5) | 4206/32313 (13.0) | 7031/37469 (18.8) | 5538/27387 (20.2) | 19525/68483 (28.5) |
| Higher education or vocational training | 1706/40135 (4.2) | 4813/58195 (8.3) | 1268/13446 (9.4) | 1669/32313 (5.2) | 4271/37469 (11.4) | 4430/27387 (16.2) | 14235/68483 (20.8) |
| Don't know/no response | 0/40135 (0.0) | 0/58195 (0.0) | 0/13446 (0.0) | 2/32313 (0.0) | 0/37469 (0.0) | 0/27387 (0.0) | 3/68483 (0.0) |
| **Literacy** | | | | | | | |
| Cannot read at all | 27085/31483 (86.0) | 13247/21076 (62.9) | 3119/5119 (60.9) | 23814/26520 (89.8) | 21875/26201 (83.5) | 14233/17430 (81.7) | 23573/34730 (67.9) |
| Able to read only parts of sentence | 1980/31483 (6.3) | 4126/21076 (19.6) | 1048/5119 (20.5) | 1536/26520 (5.8) | 2305/26201 (8.8) | 1646/17430 (9.4) | 4065/34730 (11.7) |
| Able to read whole sentence | 2408/31483 (7.6) | 3690/21076 (17.5) | 943/5119 (18.4) | 1136/26520 (4.3) | 1948/26201 (7.4) | 1543/17430 (8.9) | 7012/34730 (20.2) |
| No sentence in required language/braille | 6/31483 (0.0) | 3/21076 (0.0) | 2/5119 (0.0) | 16/26520 (0.1) | 17/26201 (0.1) | 3/17430 (0.0) | 3/34730 (0.0) |
| No response | 3/31483 (0.0) | 10/21076 (0.0) | 7/5119 (0.1) | 19/26520 (0.1) | 56/26201 (0.2) | 6/17430 (0.0) | 77/34730 (0.2) |
| **Difficulty with self-care§** | | | | | | | |
| No difficulty | – | 50964/51535 (98.9) | 11748/11938 (98.4) | – | – | – | – |
| Some difficulty | – | 508/51535 (1.0) | 163/11938 (1.4) | – | – | – | – |
| A lot of difficulty | – | 46/51535 (0.1) | 20/11938 (0.2) | – | – | – | – |
| Cannot care for self at all | – | 17/51535 (0.0) | 7/11938 (0.1) | – | – | – | – |
| No response | – | 0/51535 (0.0) | 0/11938 (0.0) | – | – | – | – |
| **Ever given birth** | | | | | | | |

| | | | | | | | |
|---|---|---|---|---|---|---|---|
| Yes | 22030/25087 (87.8) | 42430/47599 (89.1) | 9143/10179 (89.8) | 18230/20606 (88.5) | 21782/25049 (87.0) | 15606/18194 (85.8) | 38264/44185 (86.6) |
| No | 3057/25087 (12.2) | 5169/47599 (10.9) | 1036/10179 (10.2) | 2376/20606 (11.5) | 3267/25049 (13.0) | 2588/18194 (14.2) | 5921/44185 (13.4) |
| **Currently pregnant** | | | | | | | |
| Yes | – | 2625/45352 (5.8) | 502/10179 (4.9) | 3682/20119 (18.3) | 3225/24541 (13.1) | 2071/17556 (11.8) | 4612/42122 (10.9) |
| No | – | 42565/45352 (93.9) | 9655/10179 (94.8) | 15055/20119 (74.8) | 21145/24541 (86.2) | 15365/17556 (87.5) | 37270/42122 (88.5) |
| Don’t know/No response | – | 161/45352 (0.4) | 22/10179 (0.2) | 1382/20119 (6.9) | 170/24541 (0.7) | 121/17556 (0.7) | 241/42122 (0.6) |
| **Currently using contraception** | | | | | | | |
| Yes | – | 30239/42727 (70.8) | 4750/9677 (49.1) | 4483/16437 (27.3) | 8369/21315 (39.3) | 3856/15486 (24.9) | 14859/37511 (39.6) |
| No | – | 12476/42727 (29.2) | 4911/9677 (50.8) | 11504/16437 (70.0) | 12827/21315 (60.2) | 11598/15486 (74.9) | 22592/37511 (60.2) |
| No response | – | 12/42727 (0.0) | 16/9677 (0.2) | 450/16437 (2.7) | 119/21315 (0.6) | 32/15486 (0.2) | 60/37511 (0.2) |
| **Ever used contraception¶** | | | | | | | |
| Yes | – | 9316/15112 (61.6) | 1520/5429 (28.0) | 1900/15636 (12.1) | 1898/16172 (11.7) | 986/13700 (7.2) | 2866/27263 (10.5) |
| No | – | 5791/15112 (38.3) | 3905/5429 (71.9) | 13333/15636 (85.3) | 14143/16172 (87.5) | 12669/13700 (92.5) | 24327/27263 (89.2) |
| No response | – | 5/15112 (0.0) | 5/5429 (0.1) | 403/15636 (2.6) | 131/16172 (0.8) | 45/13700 (0.3) | 71/27263 (0.3) |
| **Current use of hormonal contraception‖** | | | | | | | |
| Yes | – | 23538/42407 (55.5) | 2323/9420 (24.7) | 2227/15823 (14.1) | 3986/20933 (19.0) | 1499/15146 (9.9) | 2053/36132 (5.7) |
| No | – | 18870/42407 (44.5) | 7097/9420 (75.3) | 13556/15823 (85.7) | 16927/20933 (80.9) | 13643/15146 (90.1) | 34072/36132 (94.3) |
| No response | – | 0/42407 (0.0) | 0/9420 (0.0) | 40/15823 (0.3) | 20/20933 (0.1) | 4/15146 (0.0) | 7/36132 (0.0) |
| **Current contraceptive method use**** | | | | | | | |
| Female sterilisation | – | 1351/42715 (3.2) | 1293/9661 (13.4) | 125/15987 (0.8) | 295/21196 (1.4) | 810/15454 (5.2) | 3405/37451 (9.1) |
| Male sterilisation | – | 247/42715 (0.6) | 367/9661 (3.8) | 61/15987 (0.4) | 21/21196 (0.1) | 23/15454 (0.1) | 52/37451 (0.1) |
| IUD | – | 308/42715 (0.7) | 241/9661 (2.5) | 163/15987 (1.0) | 263/21196 (1.2) | 307/15454 (2.0) | 1319/37451 (3.5) |
| Injectables | – | 5705/42715 (13.4) | 1346/9661 (13.9) | 1013/15987 (6.3) | 2934/21196 (13.8) | 792/15454 (5.1) | 1203/37451 (3.2) |
| Implants | – | 659/42715 (1.5) | 456/9661 (4.7) | 79/15987 (0.5) | 58/21196 (0.3) | 139/15454 (0.9) | 141/37451 (0.4) |
| Pill | – | 17483/42715 (40.9) | 564/9661 (5.8) | 1295/15987 (8.1) | 1455/21196 (6.9) | 655/15454 (4.2) | 805/37451 (2.1) |
| Male condom | – | 3247/42715 (7.6) | 290/9661 (3.0) | 1275/15987 (8.0) | 2872/21196 (13.5) | 1064/15454 (6.9) | 5485/37451 (14.6) |
| Female condom | – | 36/42715 (0.1) | 5/9661 (0.0) | 39/15987 (0.2) | 47/21196 (0.2) | 51/15454 (0.3) | 140/37451 (0.4) |
| Diaphragm | – | 4/42715 (0.0) | 0/9661 (0.0) | 12/15987 (0.1) | 23/21196 (0.1) | 30/15454 (0.2) | 154/37451 (0.4) |
| Foam/Jelly | – | 7/42715 (0.0) | 1/9661 (0.0) | – | – | – | – |
| Lactational Amenorrhea Method (LAM) | – | 44/42715 (0.1) | – | 534/15987 (3.3) | 151/21196 (0.7) | 87/15454 (0.6) | 530/37451 (1.4) |
| Periodic abstinence/Rhythm | – | 1375/42715 (3.2) | 61/9661 (0.6) | 583/15987 (3.6) | 199/21196 (0.9) | 82/15454 (0.5) | 284/37451 (0.8) |
| Withdrawal | – | 788/42715 (1.8) | 316/9661 (3.3) | 49/15987 (0.3) | 1253/21196 (5.9) | 87/15454 (0.6) | 2112/37451 (5.6) |
| Other | – | 29/42715 (0.1) | 6/9661 (0.1) | 26/15987 (0.2) | 26/21196 (0.1) | 45/15454 (0.3) | 85/37451 (0.2) |
| No response | – | 0/42715 (0.0) | 0/9661 (0.0) | 40/15987 (0.3) | 20/21196 (0.1) | 4/15454 (0.0) | 7/37451 (0.0) |
| **Availability of a private place to wash during last menstrual period** | | | | | | | |
| Yes | 36857/40257 (91.6) | 56287/58198 (96.7) | 11638/13446 (86.6) | 20954/32395 (64.7) | 33236/37504 (88.6) | 22902/27398 (83.6) | 61772/68491 (90.2) |
| No | 3357/40257 (8.3) | 1883/58198 (3.2) | 1802/13446 (13.4) | 10776/32395 (33.3) | 4018/37504 (10.7) | 4417/27398 (16.1) | 6504/68491 (9.5) |
| Don’t know/No response | 42/40257 (0.1) | 28/58198 (0.0) | 5/13446 (0.0) | 665/32395 (2.1) | 250/37504 (0.7) | 80/27398 (0.3) | 214/68491 (0.3) |
| **Use of menstrual materials** | | | | | | | |
| Yes | 36921/40257 (91.7) | 56148/58198 (96.5) | 12627/13446 (93.9) | 20844/32395 (64.3) | 35054/37504 (93.5) | 22335/27398 (81.5) | 60905/68491 (88.9) |
| No | 3300/40257 (8.2) | 2037/58198 (3.5) | 816/13446 (6.1) | 10847/32395 (33.5) | 2213/37504 (5.9) | 4986/27398 (18.2) | 7370/68491 (10.8) |
| Don’t know/No response | 36/40257 (0.1) | 13/58198 (0.0) | 2/13446 (0.0) | 704/32395 (2.2) | 236/37504 (0.6) | 78/27398 (0.3) | 216/68491 (0.3) |
| **Use of reusable menstrual materials††** | | | | | | | |
| Yes | 30226/40252 (75.1) | 38504/58191 (66.2) | 7925/13445 (58.9) | 9750/32164 (30.3) | 27935/37326 (74.8) | 11363/27375 (41.5) | 31425/68296 (46.0) |
| No | 10001/40252 (24.8) | 19616/58191 (33.7) | 5518/13445 (41.0) | 22150/32164 (68.9) | 9333/37326 (25.0) | 15870/27375 (58.0) | 36837/68296 (53.9) |
| Don’t know/No response | 24/40252 (0.1) | 71/58191 (0.1) | 2/13445 (0.0) | 264/32164 (0.8) | 58/37326 (0.2) | 141/27375 (0.5) | 33/68296 (0.0) |

Demographic factors are only described for sample women and girls without missing data for the outcome variable. Some variables (e.g. literacy) may have much lower sample sizes due to incomplete data collection. All values are weighted to reflect the survey sampling design. Data are n/N (%).

*Algeria: Variables not measured include: religion of household head, difficulty with self-care, current pregnancy status, ever used contraception, currently using contraception, and current contraceptive methods used.

†Religion of household head categories were unique to each survey and recoded into the representative groups; Christian, Islam/Muslim, Animist, Buddhist, Hindu, other, none, and don’t know/no response. Not all surveys measured these religions. The symbol ‘–’ in religion of household head denotes a lack of measurement for this religion. Note that Afghanistan and all provinces of Pakistan did not measure any religion of the household head.

‡Highest level of education answer options differed between surveys and were recoded for consistency into: Pre-school or none, primary, secondary, and higher education or vocational training.

§Difficulty with self-care was not measured in Afghanistan or in any provinces of Pakistan.

¶Ever use of contraception was only collected for women and girls who reported no current use of contraception.

||Hormonal contraception derived from use of contraceptive methods. Users of hormonal contraception include women and girls using injectables, implants or the pill. Non-users of hormonal contraception include women and girls using only non-hormonal contraceptive methods, or no contraception. Women and girls using IUD were excluded from these groups.
**Current contraceptive methods used: Not all surveys measured all the contraceptive methods listed. The symbol '–' denotes a lack of measurement for this contraceptive method.
††Non-users of reusable menstrual materials include both women and girls who use no menstrual materials, and those who use only disposable menstrual materials.

**Table S8: Demographic characteristics of women and girls from East Asia and the Pacific.**

| Demographics | Fiji | Kiribati | Lao PDR* | Mongolia | Samoa | Tonga | Tuvalu | Vietnam |
|---|---|---|---|---|---|---|---|---|
| **Age (years)** | | | | | | | | |
| 15-19 | 777/4726 (16.4) | 595/3519 (16.9) | 4372/22346 (19.6) | 1173/9489 (12.4) | 795/3858 (20.6) | 635/2678 (23.7) | 101/728 (13.9) | 1354/10147 (13.3) |
| 20-24 | 666/4726 (14.1) | 706/3519 (20.1) | 3682/22346 (16.5) | 1044/9489 (11.0) | 699/3858 (18.1) | 435/2678 (16.2) | 151/728 (20.8) | 1276/10147 (12.6) |
| 25-29 | 738/4726 (15.6) | 646/3519 (18.4) | 3575/22346 (16.0) | 1550/9489 (16.3) | 577/3858 (15.0) | 388/2678 (14.5) | 151/728 (20.8) | 1684/10147 (16.6) |
| 30-34 | 685/4726 (14.5) | 534/3519 (15.2) | 3376/22346 (15.1) | 1722/9489 (18.1) | 478/3858 (12.4) | 353/2678 (13.2) | 112/728 (15.4) | 1633/10147 (16.1) |
| 35-39 | 693/4726 (14.7) | 470/3519 (13.3) | 2977/22346 (13.3) | 1461/9489 (15.4) | 432/3858 (11.2) | 335/2678 (12.5) | 102/728 (14.1) | 1594/10147 (15.7) |
| 40-44 | 687/4726 (14.5) | 342/3519 (9.7) | 2633/22346 (11.8) | 1453/9489 (15.3) | 481/3858 (12.5) | 309/2678 (11.5) | 62/728 (8.5) | 1458/10147 (14.4) |
| 45-49 | 481/4726 (10.2) | 225/3519 (6.4) | 1730/22346 (7.7) | 1088/9489 (11.5) | 396/3858 (10.3) | 224/2678 (8.4) | 48/728 (6.5) | 1148/10147 (11.3) |
| **Wealth quintile** | | | | | | | | |
| Q1 (Poorest) | 789/4726 (16.7) | 604/3519 (17.2) | 3580/22346 (16.0) | 1749/9489 (18.4) | 743/3858 (19.3) | 503/2678 (18.8) | 132/728 (18.1) | 1795/10147 (17.7) |
| Q2 | 924/4726 (19.5) | 627/3519 (17.8) | 4026/22346 (18.0) | 1739/9489 (18.3) | 744/3858 (19.3) | 539/2678 (20.1) | 142/728 (19.5) | 2008/10147 (19.8) |
| Q3 | 957/4726 (20.2) | 690/3519 (19.6) | 4303/22346 (19.3) | 1906/9489 (20.1) | 765/3858 (19.8) | 556/2678 (20.8) | 143/728 (19.6) | 2110/10147 (20.8) |
| Q4 | 987/4726 (20.9) | 773/3519 (22.0) | 4900/22346 (21.9) | 1984/9489 (20.9) | 775/3858 (20.1) | 546/2678 (20.4) | 148/728 (20.3) | 2055/10147 (20.2) |
| Q5 (Richest) | 1069/4726 (22.6) | 824/3519 (23.4) | 5537/22346 (24.8) | 2111/9489 (22.2) | 831/3858 (21.5) | 533/2678 (19.9) | 164/728 (22.5) | 2179/10147 (21.5) |
| **Area** | | | | | | | | |
| Urban | 2942/4726 (62.3) | 2080/3519 (59.1) | 7896/22346 (35.3) | 6604/9489 (69.6) | 805/3858 (20.9) | 640/2678 (23.9) | 497/728 (68.2) | 3836/10147 (37.8) |
| Rural | 1784/4726 (37.7) | 1439/3519 (40.9) | 14450/22346 (64.7) | 2885/9489 (30.4) | 3053/3858 (79.1) | 2038/2678 (76.1) | 231/728 (31.8) | 6311/10147 (62.2) |
| **Religion of household head†** | | | | | | | | |
| Christian | – | 3354/3519 (95.3) | 333/22346 (1.5) | 118/9489 (1.2) | 3358/3858 (87.0) | 2591/2678 (96.8) | 703/728 (96.6) | – |
| Islam/Muslim | – | – | 2/22346 (0.0) | 363/9489 (3.8) | – | 5/2678 (0.2) | – | – |
| Animist | – | – | 6724/22346 (30.1) | – | – | – | – | – |
| Buddhist | – | – | 15227/22346 (68.1) | 4650/9489 (49.0) | – | – | – | – |
| Hindu | – | – | – | – | – | 0/2678 (0.0) | – | – |
| Other | – | 154/3519 (4.4) | 48/22346 (0.2) | 364/9489 (3.8) | 497/3858 (12.9) | 73/2678 (2.7) | 25/728 (3.4) | – |
| None | – | 11/3519 (0.3) | 10/22346 (0.0) | 3944/9489 (41.6) | 3/3858 (0.1) | 8/2678 (0.3) | 0/728 (0.0) | – |
| Don't know/No response | – | 0/3519 (0.0) | 2/22346 (0.0) | 50/9489 (0.5) | 0/3858 (0.0) | 0/2678 (0.0) | 0/728 (0.0) | – |
| **Marital/union status** | | | | | | | | |
| Currently married/in union | 2974/4726 (62.9) | 2348/3519 (66.7) | 15770/22346 (70.6) | 6602/9489 (69.6) | 2349/3858 (60.9) | 1408/2678 (52.6) | 488/728 (67.0) | 7028/10147 (69.3) |
| Formerly married/in union | 266/4726 (5.6) | 217/3519 (6.2) | 959/22346 (4.3) | 807/9489 (8.5) | 131/3858 (3.4) | 159/2678 (5.9) | 22/728 (3.1) | 657/10147 (6.5) |
| Never married/in union | 1482/4726 (31.4) | 950/3519 (27.0) | 5617/22346 (25.1) | 2080/9489 (21.9) | 1373/3858 (35.6) | 1093/2678 (40.8) | 218/728 (29.9) | 2458/10147 (24.2) |
| **Highest level of school attended‡** | | | | | | | | |
| Pre-school or none | 20/4723 (0.4) | 15/3518 (0.4) | 3301/22346 (14.8) | 324/9489 (3.4) | 11/3854 (0.3) | 4/2677 (0.2) | 3/727 (0.4) | 307/10147 (3.0) |
| Primary | 358/4723 (7.6) | 119/3518 (3.4) | 7607/22346 (34.0) | 412/9489 (4.3) | 96/3854 (2.5) | 28/2677 (1.1) | 53/727 (7.2) | 1037/10147 (10.2) |
| Secondary | 2609/4723 (55.2) | 3142/3518 (89.3) | 8360/22346 (37.4) | 4206/9489 (44.3) | 2660/3854 (69.0) | 1818/2677 (67.9) | 367/727 (50.5) | 6290/10147 (62.0) |
| Higher education or vocational training | 1736/4723 (36.8) | 242/3518 (6.9) | 3078/22346 (13.8) | 4547/9489 (47.9) | 1085/3854 (28.2) | 827/2677 (30.9) | 304/727 (41.8) | 2512/10147 (24.8) |
| Don't know/no response | 0/4723 (0.0) | 0/3518 (0.0) | 0/22346 (0.0) | 0/9489 (0.0) | 1/3854 (0.0) | 0/2677 (0.0) | 0/727 (0.0) | 1/10147 (0.0) |
| **Literacy** | | | | | | | | |
| Cannot read at all | 71/381 (18.6) | 44/135 (32.7) | 5289/10902 (48.5) | 165/503 (32.8) | 24/113 (21.4) | 8/35 (22.2) | 9/57 (16.7) | 415/1272 (32.6) |
| Able to read only parts of sentence | 119/381 (31.2) | 30/135 (22.4) | 2453/10902 (22.5) | 105/503 (20.8) | 36/113 (32.1) | 3/35 (8.3) | 5/57 (8.8) | 116/1272 (9.1) |
| Able to read whole sentence | 187/381 (49.1) | 58/135 (43.0) | 3158/10902 (29.0) | 227/503 (45.1) | 51/113 (45.6) | 24/35 (67.8) | 42/57 (74.5) | 736/1272 (57.8) |
| No sentence in required language/braille | 1/381 (0.2) | 1/135 (0.6) | 0/10902 (0.0) | 5/503 (0.9) | 1/113 (0.9) | 0/35 (0.0) | 0/57 (0.0) | 4/1272 (0.3) |
| No response | 3/381 (0.8) | 2/135 (1.3) | 1/10902 (0.0) | 2/503 (0.4) | 0/113 (0.0) | 1/35 (1.8) | 0/57 (0.0) | 1/1272 (0.1) |
| **Difficulty with self-care§** | | | | | | | | |
| No difficulty | 4146/4221 (98.2) | 3141/3193 (98.4) | – | 8450/8695 (97.2) | 3331/3386 (98.4) | 2266/2307 (98.2) | 671/676 (99.3) | – |
| Some difficulty | 70/4221 (1.7) | 48/3193 (1.5) | – | 205/8695 (2.4) | 54/3386 (1.6) | 33/2307 (1.5) | 5/676 (0.7) | – |
| A lot of difficulty | 4/4221 (0.1) | 3/3193 (0.1) | – | 36/8695 (0.4) | 1/3386 (0.0) | 4/2307 (0.2) | 0/676 (0.0) | – |
| Cannot care for self at all | 1/4221 (0.0) | 1/3193 (0.0) | – | 1/8695 (0.0) | 0/3386 (0.0) | 0/2307 (0.0) | 0/676 (0.0) | – |
| No response | 0/4221 (0.0) | 0/3193 (0.0) | – | 3/8695 (0.0) | 0/3386 (0.0) | 4/2307 (0.2) | 0/676 (0.0) | – |
| **Ever given birth** | | | | | | | | |
| Yes | 2983/4726 (63.1) | 2170/3519 (61.7) | 14969/22346 (67.0) | 7378/9489 (77.7) | 2443/3858 (63.3) | 1403/2678 (52.4) | 448/728 (61.5) | 7328/10147 (72.2) |

| | | | | | | | | |
|---|---|---|---|---|---|---|---|---|
| No | 1743/4726 (36.9) | 1349/3519 (38.3) | 7377/22346 (33.0) | 2112/9489 (22.3) | 1416/3858 (36.7) | 1274/2678 (47.6) | 280/728 (38.5) | 2818/10147 (27.8) |
| **Currently pregnant** | | | | | | | | |
| Yes | 198/4726 (4.2) | 233/3519 (6.6) | 1080/22346 (4.8) | 564/9489 (5.9) | 238/3858 (6.2) | 119/2678 (4.4) | 49/728 (6.7) | 279/10147 (2.7) |
| No | 4501/4726 (95.2) | 3226/3519 (91.7) | 21194/22346 (94.8) | 8887/9489 (93.7) | 3593/3858 (93.1) | 2519/2678 (94.1) | 673/728 (92.5) | 9858/10147 (97.2) |
| Don’t know/No response | 26/4726 (0.6) | 60/3519 (1.7) | 72/22346 (0.3) | 38/9489 (0.4) | 27/3858 (0.7) | 40/2678 (1.5) | 6/728 (0.8) | 10/10147 (0.1) |
| **Currently using contraception** | | | | | | | | |
| Yes | 1125/4528 (24.9) | 827/3286 (25.2) | 9286/21266 (43.7) | 3875/8925 (43.4) | 387/3620 (10.7) | 449/2559 (17.6) | 127/679 (18.8) | 5438/9868 (55.1) |
| No | 3394/4528 (75.0) | 2452/3286 (74.6) | 11966/21266 (56.3) | 5032/8925 (56.4) | 3208/3620 (88.6) | 2059/2559 (80.5) | 549/679 (80.8) | 4414/9868 (44.7) |
| No response | 8/4528 (0.2) | 6/3286 (0.2) | 14/21266 (0.1) | 18/8925 (0.2) | 25/3620 (0.7) | 50/2559 (2.0) | 3/679 (0.4) | 16/9868 (0.2) |
| **Ever used contraception¶** | | | | | | | | |
| Yes | 475/3601 (13.2) | 242/2692 (9.0) | 2277/13060 (17.4) | 2247/5615 (40.0) | 314/3471 (9.0) | 276/2228 (12.4) | 99/601 (16.5) | 1024/4708 (21.8) |
| No | 3117/3601 (86.6) | 2440/2692 (90.7) | 10767/13060 (82.4) | 3356/5615 (59.8) | 3122/3471 (89.9) | 1913/2228 (85.8) | 498/601 (82.9) | 3671/4708 (78.0) |
| No response | 9/3601 (0.3) | 9/2692 (0.3) | 17/13060 (0.1) | 11/5615 (0.2) | 35/3471 (1.0) | 40/2228 (1.8) | 3/601 (0.5) | 13/4708 (0.3) |
| **Current use of hormonal contraception‖** | | | | | | | | |
| Yes | 629/4437 (14.2) | 460/3264 (14.1) | 6891/20866 (33.0) | 919/6756 (13.6) | 265/3575 (7.4) | 210/2492 (8.4) | 109/676 (16.1) | 1358/8051 (16.9) |
| No | 3806/4437 (85.8) | 2802/3264 (85.8) | 13975/20866 (67.0) | 5836/6756 (86.4) | 3308/3575 (92.5) | 2276/2492 (91.3) | 567/676 (83.9) | 6691/8051 (83.1) |
| No response | 2/4437 (0.0) | 3/3264 (0.1) | 0/20866 (0.0) | 0/6756 (0.0) | 2/3575 (0.1) | 6/2492 (0.2) | 0/676 (0.0) | 1/8051 (0.0) |
| **Current contraceptive method use**** | | | | | | | | |
| Female sterilisation | 192/4519 (4.2) | 161/3280 (4.9) | 793/21251 (3.7) | 202/8907 (2.3) | 68/3595 (1.9) | 131/2508 (5.2) | 9/676 (1.4) | 113/9852 (1.1) |
| Male sterilisation | 1/4519 (0.0) | 6/3280 (0.2) | 5/21251 (0.0) | 8/8907 (0.1) | 0/3595 (0.0) | 2/2508 (0.1) | 0/676 (0.0) | 1/9852 (0.0) |
| IUD | 82/4519 (1.8) | 15/3280 (0.5) | 386/21251 (1.8) | 2151/8907 (24.2) | 20/3595 (0.5) | 22/2508 (0.9) | 0/676 (0.0) | 1801/9852 (18.3) |
| Injectables | 290/4519 (6.4) | 186/3280 (5.7) | 1536/21251 (7.2) | 179/8907 (2.0) | 183/3595 (5.1) | 82/2508 (3.3) | 72/676 (10.6) | 121/9852 (1.2) |
| Implants | 244/4519 (5.4) | 261/3280 (7.9) | 202/21251 (1.0) | 177/8907 (2.0) | 37/3595 (1.0) | 105/2508 (4.2) | 41/676 (6.1) | 19/9852 (0.2) |
| Pill | 133/4519 (3.0) | 25/3280 (0.8) | 5303/21251 (25.0) | 650/8907 (7.3) | 60/3595 (1.7) | 31/2508 (1.2) | 16/676 (2.4) | 1224/9852 (12.4) |
| Male condom | 85/4519 (1.9) | 18/3280 (0.5) | 319/21251 (1.5) | 391/8907 (4.4) | 5/3595 (0.1) | 16/2508 (0.6) | 1/676 (0.2) | 1215/9852 (12.3) |
| Female condom | 16/4519 (0.4) | 4/3280 (0.1) | 24/21251 (0.1) | 55/8907 (0.6) | 1/3595 (0.0) | 3/2508 (0.1) | 2/676 (0.3) | 53/9852 (0.5) |
| Diaphragm | 4/4519 (0.1) | 4/3280 (0.1) | 4/21251 (0.0) | 0/8907 (0.0) | – | 8/2508 (0.3) | 1/676 (0.1) | 5/9852 (0.1) |
| Foam/Jelly | 0/4519 (0.0) | – | 0/21251 (0.0) | 0/8907 (0.0) | – | 0/2508 (0.0) | – | 2/9852 (0.0) |
| Lactational Amenorrhea Method (LAM) | 3/4519 (0.1) | – | – | 5/8907 (0.1) | 7/3595 (0.2) | – | 1/676 (0.2) | 4/9852 (0.0) |
| Periodic abstinence/Rhythm | 33/4519 (0.7) | 14/3280 (0.4) | 771/21251 (3.6) | 240/8907 (2.7) | 9/3595 (0.2) | 20/2508 (0.8) | 1/676 (0.1) | 679/9852 (6.9) |
| Withdrawal | 109/4519 (2.4) | 56/3280 (1.7) | 427/21251 (2.0) | 9/8907 (0.1) | 9/3595 (0.2) | 37/2508 (1.5) | 1/676 (0.2) | 974/9852 (9.9) |
| Other | 20/4519 (0.4) | 32/3280 (1.0) | 55/21251 (0.3) | 13/8907 (0.1) | 7/3595 (0.2) | 20/2508 (0.8) | 8/676 (1.2) | 7/9852 (0.1) |
| No response | 2/4519 (0.0) | 3/3280 (0.1) | 0/21251 (0.0) | 0/8907 (0.0) | 2/3595 (0.1) | 0/2508 (0.0) | 0/676 (0.0) | 1/9852 (0.0) |
| **Availability of a private place to wash during last menstrual period** | | | | | | | | |
| Yes | 4525/4726 (95.8) | 3266/3519 (92.8) | 18091/22346 (81.0) | 8492/9489 (89.5) | 3266/3858 (84.7) | 2518/2678 (94.1) | 687/728 (94.4) | 9848/10147 (97.1) |
| No | 198/4726 (4.2) | 252/3519 (7.1) | 4249/22346 (19.0) | 970/9489 (10.2) | 567/3858 (14.7) | 159/2678 (5.9) | 40/728 (5.5) | 297/10147 (2.9) |
| Don’t know/No response | 3/4726 (0.1) | 2/3519 (0.0) | 6/22346 (0.0) | 27/9489 (0.3) | 25/3858 (0.6) | 0/2678 (0.0) | 1/728 (0.1) | 1/10147 (0.0) |
| **Use of menstrual materials** | | | | | | | | |
| Yes | 4594/4726 (97.2) | 3451/3519 (98.1) | 18281/22346 (81.8) | 8669/9489 (91.4) | 3541/3858 (91.8) | 2522/2678 (94.2) | 692/728 (95.0) | 9959/10147 (98.2) |
| No | 130/4726 (2.8) | 65/3519 (1.9) | 4063/22346 (18.2) | 809/9489 (8.5) | 310/3858 (8.0) | 156/2678 (5.8) | 36/728 (5.0) | 186/10147 (1.8) |
| Don’t know/No response | 2/4726 (0.0) | 2/3519 (0.1) | 2/22346 (0.0) | 12/9489 (0.1) | 8/3858 (0.2) | 0/2678 (0.0) | 0/728 (0.0) | 1/10147 (0.0) |
| **Use of reusable menstrual materials††** | | | | | | | | |
| Yes | 556/4725 (11.8) | 564/3518 (16.0) | 632/22346 (2.8) | 245/9489 (2.6) | 693/3856 (18.0) | 25/2678 (0.9) | 128/728 (17.6) | 120/10146 (1.2) |
| No | 4165/4725 (88.2) | 2953/3518 (83.9) | 21688/22346 (97.1) | 9225/9489 (97.2) | 3161/3856 (82.0) | 2644/2678 (98.7) | 599/728 (82.3) | 10026/10146 (98.8) |
| Don’t know/No response | 4/4725 (0.1) | 2/3518 (0.0) | 26/22346 (0.1) | 19/9489 (0.2) | 2/3856 (0.1) | 9/2678 (0.3) | 1/728 (0.1) | 0/10146 (0.0) |

Demographic factors are only described for sample women and girls without missing data for the outcome variable. Some variables (e.g. literacy) may have much lower sample sizes due to incomplete data collection. All values are weighted to reflect the survey sampling design. Data are n/N (%).

*Area response options in Lao PDR included urban, rural with road, and rural without road. These responses were recoded to urban, and rural (includes both rural with road and rural without road).

†Religion of household head categories were unique to each survey and recoded into the representative groups; Christian, Islam/Muslim, Animist, Buddhist, Hindu, other, none, and don’t know/no response. Not all surveys measured these religions. The symbol ‘–’ in religion of household head denotes a lack of measurement for this religion. Note that Fiji and Vietnam surveys did not measure any religion of the household head.

‡Highest level of education answer options differed between surveys and were recoded for consistency into: Pre-school or none, primary, secondary, and higher education or vocational training.

§Difficulty with self-care was not measured in Lao PDR or Vietnam surveys.

¶Ever use of contraception was only collected for women and girls who reported no current use of contraception.

||Hormonal contraception derived from use of contraceptive methods. Users of hormonal contraception include women and girls using injectables, implants or the pill. Non-users of hormonal contraception include women and girls using only non-hormonal contraceptive methods, or no contraception. Women and girls using IUD were excluded from these groups.
**Current contraceptive methods used: Not all surveys measured all the contraceptive methods listed. The symbol '–' denotes a lack of measurement for this contraceptive method.
††Non-users of reusable menstrual materials include both women and girls who use no menstrual materials, and those who use only disposable menstrual materials.

**Table S9: Demographic characteristics of women and girls from Europe and Central Asia.**

| Demographics | Kosovo | Kyrgyzstan | Montenegro | North Macedonia | Serbia* | Turkmenistan† | Uzbekistan |
|---|---|---|---|---|---|---|---|
| **Age (years)** | | | | | | | |
| 15-19 | 968/5020 (19.3) | 809/5175 (15.6) | 280/2156 (13.0) | 372/3023 (12.3) | 380/3527 (10.8) | 42/4946 (0.9) | 628/4360 (14.4) |
| 20-24 | 767/5020 (15.3) | 763/5175 (14.7) | 277/2156 (12.8) | 417/3023 (13.8) | 434/3527 (12.3) | 514/4946 (10.4) | 600/4360 (13.8) |
| 25-29 | 711/5020 (14.2) | 848/5175 (16.4) | 283/2156 (13.1) | 435/3023 (14.4) | 424/3527 (12.0) | 1093/4946 (22.1) | 722/4360 (16.6) |
| 30-34 | 618/5020 (12.3) | 813/5175 (15.7) | 315/2156 (14.6) | 438/3023 (14.5) | 537/3527 (15.2) | 1110/4946 (22.4) | 778/4360 (17.8) |
| 35-39 | 623/5020 (12.4) | 699/5175 (13.5) | 354/2156 (16.4) | 454/3023 (15.0) | 608/3527 (17.2) | 916/4946 (18.5) | 691/4360 (15.8) |
| 40-44 | 730/5020 (14.6) | 713/5175 (13.8) | 364/2156 (16.9) | 492/3023 (16.3) | 569/3527 (16.1) | 801/4946 (16.2) | 552/4360 (12.7) |
| 45-49 | 603/5020 (12.0) | 531/5175 (10.3) | 284/2156 (13.2) | 415/3023 (13.7) | 574/3527 (16.3) | 469/4946 (9.5) | 389/4360 (8.9) |
| **Wealth quintile** | | | | | | | |
| Q1 (Poorest) | 941/5020 (18.7) | 975/5175 (18.8) | 356/2156 (16.5) | 526/3023 (17.4) | 460/3527 (13.0) | 952/4946 (19.3) | 894/4360 (20.5) |
| Q2 | 998/5020 (19.9) | 970/5175 (18.7) | 355/2156 (16.5) | 587/3023 (19.4) | 647/3527 (18.4) | 956/4946 (19.3) | 858/4360 (19.7) |
| Q3 | 1041/5020 (20.7) | 1006/5175 (19.4) | 416/2156 (19.3) | 584/3023 (19.3) | 753/3527 (21.4) | 931/4946 (18.8) | 848/4360 (19.4) |
| Q4 | 1007/5020 (20.1) | 1011/5175 (19.5) | 515/2156 (23.9) | 646/3023 (21.4) | 809/3527 (22.9) | 985/4946 (19.9) | 858/4360 (19.7) |
| Q5 (Richest) | 1034/5020 (20.6) | 1214/5175 (23.5) | 514/2156 (23.8) | 679/3023 (22.5) | 857/3527 (24.3) | 1121/4946 (22.7) | 903/4360 (20.7) |
| **Area** | | | | | | | |
| Urban | 2136/5020 (42.5) | 2080/5175 (40.2) | 1478/2156 (68.5) | 1922/3023 (63.6) | 2215/3527 (62.8) | 2240/4946 (45.3) | 2066/4360 (47.4) |
| Rural | 2885/5020 (57.5) | 3096/5175 (59.8) | 678/2156 (31.5) | 1100/3023 (36.4) | 1312/3527 (37.2) | 2706/4946 (54.7) | 2295/4360 (52.6) |
| **Religion of household head†** | | | | | | | |
| Christian | – | – | – | 1947/3023 (64.4) | 3313/3527 (93.9) | – | – |
| Islam/Muslim | – | – | – | 1031/3023 (34.1) | 125/3527 (3.5) | – | – |
| Animist | – | – | – | – | – | – | – |
| Buddhist | – | – | – | – | – | – | – |
| Hindu | – | – | – | – | – | – | – |
| Other | – | – | – | 17/3023 (0.5) | 25/3527 (0.7) | – | – |
| None | – | – | – | 28/3023 (0.9) | 57/3527 (1.6) | – | – |
| Don’t know/No response | – | – | – | 0/3023 (0.0) | 7/3527 (0.2) | – | – |
| **Marital/union status** | | | | | | | |
| Currently married/in union | 3011/5020 (60.0) | 3651/5175 (70.5) | 1294/2156 (60.0) | 1987/3023 (65.7) | 2117/3527 (60.0) | 4449/4946 (90.0) | 3235/4360 (74.2) |
| Formerly married/in union | 112/5020 (2.2) | 405/5175 (7.8) | 104/2156 (4.8) | 150/3023 (4.9) | 285/3527 (8.1) | 497/4946 (10.0) | 289/4360 (6.6) |
| Never married/in union | 1894/5020 (37.7) | 1120/5175 (21.6) | 757/2156 (35.1) | 883/3023 (29.2) | 1124/3527 (31.9) | 0/4946 (0.0) | 836/4360 (19.2) |
| **Highest level of school attended‡** | | | | | | | |
| Pre-school or none | 108/5020 (2.1) | 6/5175 (0.1) | 54/2153 (2.5) | 44/3023 (1.5) | 11/3527 (0.3) | 0/4946 (0.0) | 6/4360 (0.1) |
| Primary | 131/5020 (2.6) | 9/5175 (0.2) | 27/2153 (1.3) | 53/3023 (1.8) | 258/3527 (7.3) | – | 6/4360 (0.1) |
| Secondary | 3215/5020 (64.0) | 3623/5175 (70.0) | 152/2153 (7.1) | 1692/3023 (56.0) | 1796/3527 (50.9) | 3804/4946 (76.9) | 3823/4360 (87.7) |
| Higher education or vocational training | 1566/5020 (31.2) | 1537/5175 (29.7) | 1921/2153 (89.2) | 1233/3023 (40.8) | 1462/3527 (41.5) | 1141/4946 (23.1) | 525/4360 (12.0) |
| Don’t know/no response | 0/5020 (0.0) | 0/5175 (0.0) | 0/2153 (0.0) | 0/3023 (0.0) | 0/3527 (0.0) | 0/4946 (0.0) | 0/4360 (0.0) |
| **Literacy** | | | | | | | |
| Cannot read at all | 196/1693 (11.6) | 8/16 (54.0) | 47/83 (56.5) | 44/97 (45.1) | 15/269 (5.6) | 1/2 (59.0) | 5/12 (44.0) |
| Able to read only parts of sentence | 297/1693 (17.6) | 3/16 (21.4) | 18/83 (21.2) | 8/97 (8.4) | 20/269 (7.3) | 0/2 (0.0) | 1/12 (4.9) |
| Able to read whole sentence | 1193/1693 (70.5) | 4/16 (24.7) | 19/83 (22.4) | 43/97 (44.1) | 234/269 (87.1) | 1/2 (41.0) | 6/12 (51.1) |
| No sentence in required language/braille | 1/1693 (0.1) | 0/16 (0.0) | 0/83 (0.0) | 2/97 (1.6) | 0/269 (0.0) | 0/2 (0.0) | 0/12 (0.0) |
| No response | 5/1693 (0.3) | 0/16 (0.0) | 0/83 (0.0) | 1/97 (0.7) | 0/269 (0.0) | 0/2 (0.0) | 0/12 (0.0) |
| **Difficulty with self-care** | | | | | | | |
| No difficulty | 4417/4480 (98.6) | 4568/4627 (98.7) | 1969/1984 (99.3) | 2750/2786 (98.7) | – | 4882/4942 (98.8) | 3878/3987 (97.3) |
| Some difficulty | 50/4480 (1.1) | 56/4627 (1.2) | 14/1984 (0.7) | 29/2786 (1.0) | – | 54/4942 (1.1) | 86/3987 (2.2) |
| A lot of difficulty | 9/4480 (0.2) | 3/4627 (0.1) | 0/1984 (0.0) | 3/2786 (0.1) | – | 4/4942 (0.1) | 20/3987 (0.5) |
| Cannot care for self at all | 0/4480 (0.0) | 0/4627 (0.0) | 0/1984 (0.0) | 0/2786 (0.0) | – | 0/4942 (0.0) | 2/3987 (0.0) |
| No response | 4/4480 (0.1) | 0/4627 (0.0) | 0/1984 (0.0) | 3/2786 (0.1) | – | 2/4942 (0.0) | 1/3987 (0.0) |
| **Ever given birth** | | | | | | | |
| Yes | 2857/5020 (56.9) | 3792/5175 (73.3) | 1305/2156 (60.5) | 1949/3023 (64.5) | 2130/3527 (60.4) | 4569/4946 (92.4) | 3192/4360 (73.2) |

| | | | | | | | |
|---|---|---|---|---|---|---|---|
| No | 2163/5020 (43.1) | 1384/5175 (26.7) | 851/2156 (39.5) | 1073/3023 (35.5) | 1397/3527 (39.6) | 377/4946 (7.6) | 1168/4360 (26.8) |
| **Currently pregnant** | | | | | | | |
| Yes | 157/5020 (3.1) | 380/5175 (7.3) | 80/2156 (3.7) | 113/3023 (3.7) | 81/3527 (2.3) | 484/4946 (9.8) | 307/4360 (7.1) |
| No | 4854/5020 (96.7) | 4776/5175 (92.3) | 2074/2156 (96.2) | 2897/3023 (95.9) | 3441/3527 (97.6) | 4417/4946 (89.3) | 4013/4360 (92.0) |
| Don’t know/No response | 9/5020 (0.2) | 20/5175 (0.4) | 2/2156 (0.1) | 13/3023 (0.4) | 5/3527 (0.1) | 45/4946 (0.9) | 40/4360 (0.9) |
| **Currently using contraception** | | | | | | | |
| Yes | 2144/4863 (44.1) | 1619/4795 (33.8) | 380/2076 (18.3) | 1495/2910 (51.4) | 1807/3446 (52.4) | 2481/4462 (55.6) | 2001/4053 (49.4) |
| No | 2716/4863 (55.9) | 3176/4795 (66.2) | 1676/2076 (80.7) | 1415/2910 (48.6) | 1626/3446 (47.2) | 1979/4462 (44.4) | 2049/4053 (50.5) |
| No response | 3/4863 (0.1) | 0/4795 (0.0) | 20/2076 (0.9) | 0/2910 (0.0) | 13/3446 (0.4) | 2/4462 (0.1) | 3/4053 (0.1) |
| **Ever used contraception§** | | | | | | | |
| Yes | 309/2876 (10.8) | 756/3556 (21.2) | 241/1776 (13.5) | 356/1528 (23.3) | 617/1720 (35.9) | 375/2465 (15.2) | 576/2359 (24.4) |
| No | 2560/2876 (89.0) | 2801/3556 (78.8) | 1522/1776 (85.7) | 1170/1528 (76.6) | 1095/1720 (63.7) | 2088/2465 (84.7) | 1779/2359 (75.4) |
| No response | 6/2876 (0.2) | 0/3556 (0.0) | 13/1776 (0.8) | 2/1528 (0.1) | 8/1720 (0.5) | 2/2465 (0.1) | 4/2359 (0.2) |
| **Current use of hormonal contraception¶** | | | | | | | |
| Yes | 84/4763 (1.8) | 146/3979 (3.7) | 32/2014 (1.6) | 39/2876 (1.4) | 92/3374 (2.7) | 65/2261 (2.9) | 97/2562 (3.8) |
| No | 4679/4763 (98.2) | 3834/3979 (96.3) | 1968/2014 (97.7) | 2832/2876 (98.4) | 3273/3374 (97.0) | 2196/2261 (97.1) | 2465/2562 (96.2) |
| No response | 0/4763 (0.0) | 0/3979 (0.0) | 15/2014 (0.7) | 5/2876 (0.2) | 9/3374 (0.3) | 0/2261 (0.0) | 0/2562 (0.0) |
| **Current contraceptive method use‖** | | | | | | | |
| Female sterilisation | 27/4860 (0.6) | 43/4795 (0.9) | 9/2056 (0.4) | 27/2910 (0.9) | 17/3433 (0.5) | 10/4460 (0.2) | 247/4050 (6.1) |
| Male sterilisation | 3/4860 (0.1) | 8/4795 (0.2) | 0/2056 (0.0) | 0/2910 (0.0) | 1/3433 (0.0) | 3/4460 (0.1) | 2/4050 (0.0) |
| IUD | 97/4860 (2.0) | 816/4795 (17.0) | 42/2056 (2.0) | 33/2910 (1.2) | 59/3433 (1.7) | 2199/4460 (49.3) | 1488/4050 (36.7) |
| Injectables | 3/4860 (0.1) | 15/4795 (0.3) | 0/2056 (0.0) | 1/2910 (0.0) | 0/3433 (0.0) | 14/4460 (0.3) | 63/4050 (1.5) |
| Implants | 2/4860 (0.0) | – | 1/2056 (0.1) | 0/2910 (0.0) | 0/3433 (0.0) | 0/4460 (0.0) | 3/4050 (0.1) |
| Pill | 79/4860 (1.6) | 153/4795 (3.2) | 31/2056 (1.5) | 39/2910 (1.4) | 92/3433 (2.7) | 55/4460 (1.2) | 77/4050 (1.9) |
| Male condom | 108/4860 (2.2) | 591/4795 (12.3) | 162/2056 (7.9) | 420/2910 (14.4) | 633/3433 (18.4) | 94/4460 (2.1) | 96/4050 (2.4) |
| Female condom | 0/4860 (0.0) | 5/4795 (0.1) | 0/2056 (0.0) | 1/2910 (0.0) | 2/3433 (0.1) | 0/4460 (0.0) | 2/4050 (0.0) |
| Diaphragm | 0/4860 (0.0) | 0/4795 (0.0) | 0/2056 (0.0) | 1/2910 (0.0) | 0/3433 (0.0) | 0/4460 (0.0) | 0/4050 (0.0) |
| Foam/Jelly | 0/4860 (0.0) | 2/4795 (0.0) | 0/2056 (0.0) | 0/2910 (0.0) | 1/3433 (0.0) | 1/4460 (0.0) | 1/4050 (0.0) |
| Lactational Amenorrhea Method (LAM) | – | – | – | – | – | – | 9/4050 (0.2) |
| Periodic abstinence/Rhythm | 1/4860 (0.0) | 24/4795 (0.5) | 21/2056 (1.0) | 77/2910 (2.6) | 285/3433 (8.3) | 44/4460 (1.0) | 10/4050 (0.3) |
| Withdrawal | 1857/4860 (38.2) | 57/4795 (1.2) | 102/2056 (4.9) | 1096/2910 (37.7) | 1017/3433 (29.6) | 114/4460 (2.5) | 66/4050 (1.6) |
| Other | 5/4860 (0.1) | 1/4795 (0.0) | 0/2056 (0.0) | 1/2910 (0.0) | 0/3433 (0.0) | 5/4460 (0.1) | 11/4050 (0.3) |
| No response | 0/4860 (0.0) | 0/4795 (0.0) | 15/2056 (0.7) | 5/2910 (0.2) | 9/3433 (0.3) | 0/4460 (0.0) | 0/4050 (0.0) |
| **Availability of a private place to wash during last menstrual period** | | | | | | | |
| Yes | 4943/5020 (98.5) | 4828/5175 (93.3) | 2094/2156 (97.1) | 2952/3023 (97.7) | 3489/3527 (98.9) | 4890/4946 (98.9) | 4216/4360 (96.7) |
| No | 73/5020 (1.5) | 346/5175 (6.7) | 56/2156 (2.6) | 70/3023 (2.3) | 37/3527 (1.1) | 55/4946 (1.1) | 142/4360 (3.3) |
| Don’t know/No response | 4/5020 (0.1) | 2/5175 (0.0) | 6/2156 (0.3) | 0/3023 (0.0) | 0/3527 (0.0) | 1/4946 (0.0) | 2/4360 (0.0) |
| **Use of menstrual materials** | | | | | | | |
| Yes | 4979/5020 (99.2) | 5018/5175 (97.0) | 2092/2156 (97.0) | 2980/3023 (98.6) | 3469/3527 (98.4) | 4902/4946 (99.1) | 4209/4360 (96.5) |
| No | 38/5020 (0.8) | 157/5175 (3.0) | 58/2156 (2.7) | 43/3023 (1.4) | 58/3527 (1.6) | 43/4946 (0.9) | 149/4360 (3.4) |
| Don’t know/No response | 3/5020 (0.1) | 0/5175 (0.0) | 6/2156 (0.3) | 0/3023 (0.0) | 0/3527 (0.0) | 1/4946 (0.0) | 2/4360 (0.0) |
| **Use of reusable menstrual materials**** | | | | | | | |
| Yes | 162/5018 (3.2) | 933/5175 (18.0) | 87/2150 (4.0) | 26/3023 (0.9) | 18/3527 (0.5) | 42/4945 (0.8) | 624/4359 (14.3) |
| No | 4851/5018 (96.7) | 4231/5175 (81.7) | 2060/2150 (95.8) | 2996/3023 (99.1) | 3509/3527 (99.5) | 4903/4945 (99.2) | 3728/4359 (85.5) |
| Don’t know/No response | 4/5018 (0.1) | 12/5175 (0.2) | 3/2150 (0.2) | 0/3023 (0.0) | 0/3527 (0.0) | 0/4945 (0.0) | 7/4359 (0.2) |

Demographic factors are only described for sample women and girls without missing data for the outcome variable. Some variables (e.g. literacy) may have much lower sample sizes due to incomplete data collection. All values are weighted to reflect the survey sampling design. Data are n/N (%).

*Serbia: Difficulty with selfcare not measured.

†Religion of household head categories were unique to each survey and recoded into the representative groups; Christian, Islam/Muslim, Animist, Buddhist, Hindu, other, none, and don’t know/no response. Not all surveys measured these religions. The symbol ‘–’ in religion of household head denotes a lack of measurement for this religion. Note that Kosovo, Kyrgyzstan, Montenegro, Turkmenistan and Uzbekistan surveys did not measure any religion of the household head.

‡Highest level of education answer options differed between surveys and were recoded for consistency into: Pre-school or none, primary, secondary, and higher education or vocational training. In Turkmenistan, secondary level of education represents women and girls with primary and secondary maximum educational attainment.

§Ever use of contraception was only collected for women and girls who reported no current use of contraception.
¶Hormonal contraception derived from use of contraceptive methods. Users of hormonal contraception include women and girls using injectables, implants or the pill. Non-users of hormonal contraception include women and girls using only non-hormonal contraceptive methods, or no contraception. Women and girls using IUD were excluded from these groups.
||Current contraceptive methods used: Not all surveys measured all the contraceptive methods listed. The symbol '–' denotes a lack of measurement for this contraceptive method.
**Non-users of reusable menstrual materials include both women and girls who use no menstrual materials, and those who use only disposable menstrual materials.

**Table S10: Demographic characteristics of women and girls from Latin America and Caribbean.**

| Demographics | Costa Rica | Cuba | Dominican Republic | Guyana | Honduras | Suriname | Turks & Caicos Islands* |
|---|---|---|---|---|---|---|---|
| **Age (years)** | | | | | | | |
| 15-19 | 974/6795 (14.3) | 1024/8243 (12.4) | 3680/20677 (17.8) | 973/5411 (18.0) | 3524/17572 (20.1) | 1319/6441 (20.5) | 54/789 (6.8) |
| 20-24 | 1126/6795 (16.6) | 987/8243 (12.0) | 3652/20677 (17.7) | 1021/5411 (18.9) | 3075/17572 (17.5) | 965/6441 (15.0) | 110/789 (14.0) |
| 25-29 | 1087/6795 (16.0) | 1177/8243 (14.3) | 3521/20677 (17.0) | 893/5411 (16.5) | 2781/17572 (15.8) | 916/6441 (14.2) | 92/789 (11.6) |
| 30-34 | 1085/6795 (16.0) | 1339/8243 (16.2) | 2982/20677 (14.4) | 644/5411 (11.9) | 2400/17572 (13.7) | 942/6441 (14.6) | 137/789 (17.4) |
| 35-39 | 985/6795 (14.5) | 1049/8243 (12.7) | 2785/20677 (13.5) | 572/5411 (10.6) | 2211/17572 (12.6) | 885/6441 (13.7) | 179/789 (22.7) |
| 40-44 | 838/6795 (12.3) | 1244/8243 (15.1) | 2357/20677 (11.4) | 707/5411 (13.1) | 2126/17572 (12.1) | 749/6441 (11.6) | 118/789 (14.9) |
| 45-49 | 700/6795 (10.3) | 1422/8243 (17.3) | 1699/20677 (8.2) | 602/5411 (11.1) | 1454/17572 (8.3) | 665/6441 (10.3) | 99/789 (12.6) |
| **Wealth quintile** | | | | | | | |
| Q1 (Poorest) | 1245/6795 (18.3) | 1524/8243 (18.5) | 3379/20677 (16.3) | 865/5411 (16.0) | 2749/17572 (15.6) | 1182/6441 (18.3) | 146/789 (18.5) |
| Q2 | 1351/6795 (19.9) | 1524/8243 (18.5) | 4084/20677 (19.8) | 1008/5411 (18.6) | 3286/17572 (18.7) | 1302/6441 (20.2) | 165/789 (20.9) |
| Q3 | 1447/6795 (21.3) | 1607/8243 (19.5) | 4283/20677 (20.7) | 1150/5411 (21.3) | 3607/17572 (20.5) | 1363/6441 (21.2) | 160/789 (20.2) |
| Q4 | 1389/6795 (20.4) | 1707/8243 (20.7) | 4617/20677 (22.3) | 1233/5411 (22.8) | 4030/17572 (22.9) | 1337/6441 (20.8) | 174/789 (22.1) |
| Q5 (Richest) | 1363/6795 (20.1) | 1881/8243 (22.8) | 4313/20677 (20.9) | 1154/5411 (21.3) | 3900/17572 (22.2) | 1256/6441 (19.5) | 144/789 (18.3) |
| **Area** | | | | | | | |
| Urban | 4886/6795 (71.9) | 5395/8243 (65.5) | 15878/20677 (76.8) | 1320/5411 (24.4) | 8664/17572 (49.3) | 4866/6441 (75.6) | 760/789 (96.2) |
| Rural | 1909/6795 (28.1) | 2847/8243 (34.5) | 4798/20677 (23.2) | 4090/5411 (75.6) | 8908/17572 (50.7) | 1099/6441 (17.1) | 30/789 (3.8) |
| **Religion of household head†** | | | | | | | |
| Christian | 6199/6795 (91.2) | – | 15808/20677 (76.5) | 3497/5411 (64.6) | – | 3427/6441 (53.2) | – |
| Islam/Muslim | – | – | – | 415/5411 (7.7) | – | 808/6441 (12.5) | – |
| Animist | – | – | – | – | – | – | – |
| Buddhist | – | – | – | – | – | – | – |
| Hindu | – | – | – | 1380/5411 (25.5) | – | 1516/6441 (23.5) | – |
| Other | 93/6795 (1.4) | – | 294/20677 (1.4) | 72/5411 (1.3) | – | 234/6441 (3.6) | – |
| None | 471/6795 (6.9) | – | 4575/20677 (22.1) | 47/5411 (0.9) | – | 411/6441 (6.4) | – |
| Don’t know/No response | 32/6795 (0.5) | – | 0/20677 (0.0) | 0/5411 (0.0) | – | 45/6441 (0.7) | – |
| **Marital/union status** | | | | | | | |
| Currently married/in union | 3313/6795 (48.8) | 4833/8243 (58.6) | 10883/20677 (52.6) | 3596/5411 (66.5) | 9271/17572 (52.8) | 4373/6441 (67.9) | 500/789 (63.4) |
| Formerly married/in union | 1160/6795 (17.1) | 1492/8243 (18.1) | 4492/20677 (21.7) | 556/5411 (10.3) | 2992/17572 (17.0) | 728/6441 (11.3) | 123/789 (15.6) |
| Never married/in union | 2317/6795 (34.1) | 1865/8243 (22.6) | 5298/20677 (25.6) | 1247/5411 (23.1) | 5301/17572 (30.2) | 1224/6441 (19.0) | 165/789 (20.8) |
| **Highest level of school attended‡** | | | | | | | |
| Pre-school or none | 78/6795 (1.1) | 17/8243 (0.2) | 352/20672 (1.7) | 63/5405 (1.2) | 448/17571 (2.6) | 226/6441 (3.5) | 6/789 (0.8) |
| Primary | 1423/6795 (20.9) | 174/8243 (2.1) | 3810/20672 (18.4) | 457/5405 (8.5) | 8453/17571 (48.1) | 841/6441 (13.1) | 6/789 (0.7) |
| Secondary | 3090/6795 (45.5) | 1428/8243 (17.3) | 8816/20672 (42.6) | 3857/5405 (71.4) | 6334/17571 (36.0) | 4449/6441 (69.1) | 400/789 (50.7) |
| Higher education or vocational training | 2204/6795 (32.4) | 6620/8243 (80.3) | 7693/20672 (37.2) | 1009/5405 (18.7) | 2336/17571 (13.3) | 906/6441 (14.1) | 377/789 (47.8) |
| Don’t know/no response | 0/6795 (0.0) | 3/8243 (0.0) | 1/20672 (0.0) | 18/5405 (0.3) | 0/17571 (0.0) | 18/6441 (0.3) | 0/789 (0.0) |
| **Literacy** | | | | | | | |
| Cannot read at all | 85/1500 (5.7) | 24/194 (12.2) | 944/4159 (22.7) | 182/544 (33.5) | 913/8902 (10.3) | 324/1085 (29.8) | 4/30 (12.6) |
| Able to read only parts of sentence | 67/1500 (4.4) | 16/194 (8.2) | 734/4159 (17.7) | 169/544 (31.2) | 478/8902 (5.4) | 174/1085 (16.0) | 12/30 (38.3) |
| Able to read whole sentence | 1341/1500 (89.4) | 152/194 (78.3) | 2322/4159 (55.8) | 135/544 (24.8) | 7499/8902 (84.2) | 540/1085 (49.7) | 14/30 (45.4) |
| No sentence in required language/braille | 0/1500 (0.0) | 2/194 (0.8) | 133/4159 (3.2) | 20/544 (3.7) | 5/8902 (0.1) | 36/1085 (3.3) | 1/30 (3.3) |
| No response | 7/1500 (0.5) | 1/194 (0.5) | 26/4159 (0.6) | 37/544 (6.9) | 7/8902 (0.1) | 13/1085 (1.2) | 0/30 (0.5)†† |
| **Difficulty with self-care** | | | | | | | |
| No difficulty | 6132/6213 (98.7) | 7541/7630 (98.8) | 18335/18572 (98.7) | 4710/4834 (97.4) | 15216/15554 (97.8) | 5556/5642 (98.5) | – |
| Some difficulty | 61/6213 (1.0) | 82/7630 (1.1) | 223/18572 (1.2) | 115/4834 (2.4) | 302/15554 (1.9) | 65/5642 (1.2) | – |
| A lot of difficulty | 5/6213 (0.1) | 7/7630 (0.1) | 9/18572 (0.0) | 8/4834 (0.2) | 26/15554 (0.2) | 6/5642 (0.1) | – |
| Cannot care for self at all | 15/6213 (0.2) | 0/7630 (0.0) | 3/18572 (0.0) | 0/4834 (0.0) | 2/15554 (0.0) | 0/5642 (0.0) | – |
| No response | 0/6213 (0.0) | 0/7630 (0.0) | 1/18572 (0.0) | 1/4834 (0.0) | 7/15554 (0.0) | 15/5642 (0.3) | – |
| **Ever given birth** | | | | | | | |

| | | | | | | | |
|---|---|---|---|---|---|---|---|
| Yes | 4254/6795 (62.6) | 5956/8243 (72.3) | 14156/20677 (68.5) | 3427/5411 (63.3) | 12092/17572 (68.8) | 4030/6441 (62.6) | 515/789 (65.2) |
| No | 2542/6795 (37.4) | 2287/8243 (27.7) | 6520/20677 (31.5) | 1983/5411 (36.7) | 5480/17572 (31.2) | 2410/6441 (37.4) | 275/789 (34.8) |
| **Currently pregnant** | | | | | | | |
| Yes | 225/6795 (3.3) | 230/8243 (2.8) | 886/20677 (4.3) | 216/5411 (4.0) | 805/17572 (4.6) | 229/6441 (3.6) | 18/789 (2.3) |
| No | 6537/6795 (96.2) | 7992/8243 (97.0) | 19635/20677 (95.0) | 5142/5411 (95.0) | 16668/17572 (94.9) | 6151/6441 (95.5) | 750/789 (95.1) |
| Don't know/No response | 33/6795 (0.5) | 21/8243 (0.3) | 156/20677 (0.8) | 52/5411 (1.0) | 99/17572 (0.6) | 61/6441 (0.9) | 21/789 (2.7) |
| **Currently using contraception** | | | | | | | |
| Yes | 3742/6570 (57.0) | 5738/8013 (71.6) | 9902/19791 (50.0) | 1232/5195 (23.7) | 8024/16767 (47.9) | 1922/6212 (30.9) | 264/771 (34.2) |
| No | 2821/6570 (42.9) | 2249/8013 (28.1) | 9862/19791 (49.8) | 3947/5195 (76.0) | 8721/16767 (52.0) | 4265/6212 (68.7) | 505/771 (65.5) |
| No response | 7/6570 (0.1) | 26/8013 (0.3) | 27/19791 (0.1) | 15/5195 (0.3) | 22/16767 (0.1) | 25/6212 (0.4) | 2/771 (0.3) |
| **Ever used contraception§** | | | | | | | |
| Yes | 1429/3053 (46.8) | 1009/2505 (40.3) | 4344/10775 (40.3) | 707/4179 (16.9) | 3416/9547 (35.8) | 978/4518 (21.6) | 99/526 (18.8) |
| No | 1622/3053 (53.1) | 1478/2505 (59.0) | 6404/10775 (59.4) | 3455/4179 (82.7) | 6125/9547 (64.2) | 3515/4518 (77.8) | 424/526 (80.6) |
| No response | 2/3053 (0.1) | 18/2505 (0.7) | 27/10775 (0.2) | 16/4179 (0.4) | 7/9547 (0.1) | 26/4518 (0.6) | 3/526 (0.5) |
| **Current use of hormonal contraception¶** | | | | | | | |
| Yes | 1950/6411 (30.4) | 1053/6161 (17.1) | 4734/19297 (24.5) | 558/4991 (11.2) | 3523/15999 (22.0) | 1454/6106 (23.8) | 130/761 (17.1) |
| No | 4461/6411 (69.6) | 5101/6161 (82.8) | 14563/19297 (75.5) | 4431/4991 (88.8) | 12469/15999 (77.9) | 4650/6106 (76.2) | 631/761 (82.9) |
| No response | 0/6411 (0.0) | 7/6161 (0.1) | 0/19297 (0.0) | 2/4991 (0.0) | 7/15999 (0.0) | 3/6106 (0.0) | 0/761 (0.0) |
| **Current contraceptive method use‖** | | | | | | | |
| Female sterilisation | 1032/6563 (15.7) | 1294/7987 (16.2) | 4296/19764 (21.7) | 129/5179 (2.5) | 2878/16745 (17.2) | 207/6187 (3.3) | 26/769 (3.4) |
| Male sterilisation | 189/6563 (2.9) | 1/7987 (0.0) | 4/19764 (0.0) | 4/5179 (0.1) | 27/16745 (0.2) | 0/6187 (0.0) | 3/769 (0.5) |
| IUD | 152/6563 (2.3) | 1826/7987 (22.9) | 467/19764 (2.4) | 188/5179 (3.6) | 746/16745 (4.5) | 81/6187 (1.3) | 8/769 (1.1) |
| Injectables | 462/6563 (7.0) | 79/7987 (1.0) | 1554/19764 (7.9) | 178/5179 (3.4) | 1893/16745 (11.3) | 184/6187 (3.0) | 32/769 (4.2) |
| Implants | 33/6563 (0.5) | 77/7987 (1.0) | 406/19764 (2.1) | 126/5179 (2.4) | 315/16745 (1.9) | 5/6187 (0.1) | 3/769 (0.4) |
| Pill | 1475/6563 (22.5) | 901/7987 (11.3) | 2807/19764 (14.2) | 260/5179 (5.0) | 1331/16745 (7.9) | 1268/6187 (20.5) | 95/769 (12.3) |
| Male condom | 487/6563 (7.4) | 1691/7987 (21.2) | 318/19764 (1.6) | 264/5179 (5.1) | 578/16745 (3.5) | 161/6187 (2.6) | 106/769 (13.7) |
| Female condom | 13/6563 (0.2) | 34/7987 (0.4) | 13/19764 (0.1) | 25/5179 (0.5) | 6/16745 (0.0) | 11/6187 (0.2) | 3/769 (0.3) |
| Diaphragm | 0/6563 (0.0) | 0/7987 (0.0) | 6/19764 (0.0) | 13/5179 (0.3) | 6/16745 (0.0) | 1/6187 (0.0) | 3/769 (0.4) |
| Foam/Jelly | 2/6563 (0.0) | 6/7987 (0.1) | 0/19764 (0.0) | 2/5179 (0.0) | 0/16745 (0.0) | 0/6187 (0.0) | 0/769 (0.0) |
| Lactational Amenorrhea Method (LAM) | – | 2/7987 (0.0) | – | 12/5179 (0.2) | – | – | – |
| Periodic abstinence/Rhythm | 81/6563 (1.2) | 20/7987 (0.2) | 53/19764 (0.3) | 14/5179 (0.3) | 182/16745 (1.1) | 7/6187 (0.1) | 2/769 (0.2) |
| Withdrawal | 7/6563 (0.1) | 2/7987 (0.0) | 38/19764 (0.2) | 13/5179 (0.3) | 131/16745 (0.8) | 6/6187 (0.1) | 1/769 (0.1) |
| Other | 41/6563 (0.6) | 61/7987 (0.8) | 48/19764 (0.2) | 33/5179 (0.6) | 28/16745 (0.2) | 12/6187 (0.2) | 5/769 (0.7) |
| No response | 0/6563 (0.0) | 7/7987 (0.1) | 0/19764 (0.0) | 2/5179 (0.0) | 7/16745 (0.0) | 3/6187 (0.0) | 0/769 (0.0) |
| **Availability of a private place to wash during last menstrual period** | | | | | | | |
| Yes | 6724/6795 (99.0) | 7869/8243 (95.5) | 19706/20677 (95.3) | 5051/5411 (93.3) | 16963/17572 (96.5) | 6185/6441 (96.0) | 761/789 (96.5) |
| No | 64/6795 (0.9) | 364/8243 (4.4) | 945/20677 (4.6) | 356/5411 (6.6) | 601/17572 (3.4) | 237/6441 (3.7) | 28/789 (3.5) |
| Don't know/No response | 6/6795 (0.1) | 10/8243 (0.1) | 26/20677 (0.1) | 4/5411 (0.1) | 7/17572 (0.0) | 18/6441 (0.3) | 0/789 (0.0) |
| **Use of menstrual materials** | | | | | | | |
| Yes | 6710/6795 (98.7) | 8048/8243 (97.6) | 20330/20677 (98.3) | 5219/5411 (96.5) | 17240/17572 (98.1) | 5983/6441 (92.9) | 785/789 (99.4) |
| No | 79/6795 (1.2) | 187/8243 (2.3) | 321/20677 (1.6) | 188/5411 (3.5) | 327/17572 (1.9) | 442/6441 (6.9) | 5/789 (0.6) |
| Don't know/No response | 6/6795 (0.1) | 7/8243 (0.1) | 26/20677 (0.1) | 3/5411 (0.1) | 5/17572 (0.0) | 16/6441 (0.2) | 0/789 (0.0) |
| **Use of reusable menstrual materials**** | | | | | | | |
| Yes | 131/6789 (1.9) | 216/8240 (2.6) | 455/20656 (2.2) | 106/5408 (2.0) | 538/17570 (3.1) | 227/6428 (3.5) | 9/789 (1.1) |
| No | 6627/6789 (97.6) | 8018/8240 (97.3) | 20145/20656 (97.5) | 5299/5408 (98.0) | 16976/17570 (96.6) | 6192/6428 (96.3) | 762/789 (96.6) |
| Don't know/No response | 30/6789 (0.4) | 5/8240 (0.1) | 56/20656 (0.3) | 3/5408 (0.1) | 56/17570 (0.3) | 9/6428 (0.1) | 18/789 (2.3) |

Demographic factors are only described for sample women and girls without missing data for the outcome variable. Some variables (e.g. literacy) may have much lower sample sizes due to incomplete data collection. All values are weighted to reflect the survey sampling design. Data are n/N (%).

*Turkmenistan: Difficulty with self-care and religion of household head not measured.

†Religion of household head categories were unique to each survey and recoded into the representative groups; Christian, Islam/Muslim, Animist, Buddhist, Hindu, other, none, and don't know/no response. Not all surveys measured these religions. The symbol '–' in religion of household head denotes a lack of measurement for this religion. Note that Cuba, Honduras and Turks & Caicos Islands surveys did not measure any religion of the household head.

‡Highest level of education answer options differed between surveys and were recoded for consistency into: Pre-school or none, primary, secondary, and higher education or vocational training.

§Ever use of contraception was only collected for women and girls who reported no current use of contraception.

¶Hormonal contraception derived from use of contraceptive methods. Users of hormonal contraception include women and girls using injectables, implants or the pill. Non-users of hormonal contraception include women and girls using only non-hormonal contraceptive methods, or no contraception. Women and girls using IUD were excluded from these groups.
||Current contraceptive methods used: Not all surveys measured all the contraceptive methods listed. The symbol '–' denotes a lack of measurement for this contraceptive method.
**Non-users of reusable menstrual materials include both women and girls who use no menstrual materials, and those who use only disposable menstrual materials.
††After applying survey weights, 0.15 (rounded to 0) women in Turks and Caicos Islands had no response for literacy, representing 0.5% of the sample.

**Table S11: Prevalence of menstrual-related absenteeism in study surveys.**

| Country | Prevalence of menstrual-related absenteeism | | | |
|---|---|---|---|---|
| | **Yes** | **No** | **DK or no such activity** | **No response** |
| **West and Central Africa** | | | | |
| Central African Republic (N = 7093) | 2209 (31.1) | 4850 (68.4) | 34 (0.5) | 0 (0.0) |
| Chad (N = 18211) | 5942 (32.6) | 12196 (67.0) | 64 (0.4) | 9 (0.1) |
| Democratic Republic of Congo (N = 16987) | 2412 (14.2) | 14432 (85.0) | 140 (0.8) | 3 (0.0) |
| Gambia (N = 12177) | 2464 (20.2) | 9683 (79.5) | 29 (0.2) | 2 (0.0) |
| Ghana (N = 12855) | 2426 (18.9) | 10378 (80.7) | 51 (0.4) | 0 (0.0) |
| Guinea-Bissau (N = 10913) | 892 (8.2) | 9984 (91.5) | 37 (0.3) | 0 (0.0) |
| Nigeria (N = 33195) | 5502 (16.6) | 27498 (82.8) | 185 (0.6) | 9 (0.0) |
| Sao Tome & Principe (N = 2858) | 313 (11.0) | 2508 (87.8) | 37 (1.3) | 0 (0.0) |
| Sierra Leone (N = 13700) | 2760 (20.1) | 10755 (78.5) | 183 (1.3) | 2 (0.0) |
| Togo (N = 6080) | 737 (12.1) | 5337 (87.8) | 5 (0.1) | 0 (0.0) |
| **Pooled prevalence (95% CI)** | **18.5 (13.5, 23.5)** | **80.9 (75.9, 85.9)** | **0.6 (0.3, 0.8)** | **0.0 (0.0, 0.0)** |
| **Eastern and Southern Africa** | | | | |
| Lesotho (N = 5648) | 742 (13.1) | 4900 (86.8) | 4 (0.1) | 1 (0.0) |
| Madagascar (N = 14060) | 1168 (8.3) | 12867 (91.5) | 23 (0.2) | 2 (0.0) |
| Malawi (N = 20498) | 2601 (12.7) | 16577 (80.9) | 1317 (6.4) | 4 (0.0) |
| Zimbabwe (N = 8543) | 1393 (16.3) | 7147 (83.7) | 2 (0.0) | 1 (0.0) |
| **Pooled prevalence (95% CI)** | **12.6 (9.4, 15.8)** | **85.7 (81.2, 90.2)** | **1.6 (0.0, 4.7)*** | **0.0 (0.0, 0.0)** |
| **Middle East and North Africa** | | | | |
| Algeria (N = 33078) | 8028 (24.3) | 24777 (74.9) | 252 (0.8) | 21 (0.1) |
| Iraq (N = 19733) | 2095 (10.6) | 17596 (89.2) | 41 (0.2) | 1 (0.0) |
| State of Palestine (N = 6425) | 893 (13.9) | 5501 (85.6) | 30 (0.5) | 1 (0.0) |
| Tunisia (N = 5668) | 622 (11.0) | 5018 (88.5) | 28 (0.5) | 0 (0.0) |
| **Pooled prevalence (95% CI)** | **14.9 (8.7, 21.2)** | **84.6 (78.1, 91.0)** | **0.5 (0.2, 0.7)** | **0.0 (0.0, 0.1)*** |
| **South Asia** | | | | |
| Afghanistan (N = 40257) | 12163 (30.2) | 26516 (65.9) | 1569 (3.9) | 8 (0.0) |
| Bangladesh (N = 58198) | 4574 (7.9) | 43714 (75.1) | 9895 (17.0) | 16 (0.0) |
| Nepal (N = 13446)† | 1265 (9.4) | 12179 (90.6) | – | 1 (0.0) |
| Pakistan (Balochistan) (N = 32395) | 6355 (19.6) | 22151 (68.4) | 3666 (11.3) | 222 (0.7) |
| Pakistan (Khyber Pakhtunkhwa) (N = 37504) | 6090 (16.2) | 27908 (74.4) | 3386 (9.0) | 119 (0.3) |
| Pakistan (Sindh) (N = 27398) | 10404 (38.0) | 16188 (59.1) | 783 (2.9) | 23 (0.1) |
| Pakistan (Punjab) (N = 68491) | 11389 (16.6) | 55369 (80.8) | 1543 (2.3) | 190 (0.3) |
| **Pooled prevalence (95% CI)** | **19.7 (11.6, 27.8)** | **73.5 (65.8, 81.1)** | **7.7 (3.1, 12.4)** | **0.2 (0.0, 0.4)** |
| **East Asia and the Pacific** | | | | |
| Fiji (N = 4726) | 1093 (23.1) | 3609 (76.4) | 23 (0.5) | 2 (0.0) |
| Kiribati (N = 3519) | 566 (16.1) | 2943 (83.6) | 10 (0.3) | 1 (0.0) |
| Lao PDR (N = 22346) | 2646 (11.8) | 19480 (87.2) | 221 (1.0) | 0 (0.0) |
| Mongolia (N = 9489) | 303 (3.2) | 9159 (96.5) | 28 (0.3) | 0 (0.0) |
| Samoa (N = 3858) | 346 (9.0) | 3450 (89.4) | 59 (1.5) | 3 (0.1) |
| Tonga (N = 2678) | 417 (15.6) | 2255 (84.2) | 5 (0.2) | 0 (0.0) |
| Tuvalu (N = 728) | 114 (15.6) | 613 (84.2) | 1 (0.1) | 0 (0.0) |
| Vietnam (N = 10147) | 404 (4.0) | 9667 (95.3) | 69 (0.7) | 7 (0.1) |
| **Pooled prevalence (95% CI)** | **12.2 (7.6, 16.9)** | **87.2 (82.6, 91.7)** | **0.5 (0.2, 0.8)** | **0.0 (0.0, 0.1)** |
| **Europe and Central Asia** | | | | |
| Kosovo (N = 5020) | 525 (10.5) | 4018 (80.0) | 474 (9.5) | 3 (0.1) |
| Kyrgyzstan (N = 5175) | 362 (7.0) | 4796 (92.7) | 17 (0.3) | 0 (0.0) |
| Montenegro (N = 2156) | 145 (6.7) | 2006 (93.1) | 3 (0.2) | 1 (0.0) |
| Republic of North Macedonia (N = 3023) | 200 (6.6) | 2822 (93.3) | 2 (0.1) | 0 (0.0) |
| Serbia (N = 3527) | 323 (9.2) | 3201 (90.8) | 3 (0.1) | 0 (0.0) |
| Turkmenistan (N = 4946) | 43 (0.9) | 4899 (99.1) | 3 (0.1) | 1 (0.0) |
| Uzbekistan (N = 4360) | 315 (7.2) | 4025 (92.3) | 21 (0.5) | 0 (0.0) |
| **Pooled prevalence (95% CI)** | **6.8 (4.6, 9.1)** | **91.6 (87.4, 95.9)** | **1.5 (0.0, 4.0)*** | **0.0 (0.0, 0.1)*** |
| **Latin America and Caribbean** | | | | |
| Costa Rica (N = 6795) | 461 (6.8) | 6273 (92.3) | 56 (0.8) | 5 (0.1) |
| Cuba (N = 8243) | 2271 (27.5) | 5922 (71.8) | 47 (0.6) | 3 (0.0) |
| Dominican Republic (N = 20677) | 4523 (21.9) | 16048 (77.6) | 81 (0.4) | 24 (0.1) |
| Guyana (N = 5411) | 1096 (20.3) | 4292 (79.3) | 16 (0.3) | 7 (0.1) |
| Honduras (N = 17572) | 3382 (19.2) | 14123 (80.4) | 53 (0.3) | 14 (0.1) |
| Suriname (N = 6441) | 1126 (17.5) | 5275 (81.9) | 23 (0.4) | 16 (0.2) |
| Turks & Caicos Islands (N = 789) | 101 (12.7) | 683 (86.5) | 5 (0.6) | 1 (0.1) |
| **Pooled prevalence (95% CI)** | **18.0 (13.1, 22.9)** | **81.4 (76.5, 86.2)** | **0.4 (0.3, 0.5)** | **0.1 (0.0, 0.1)** |
| **All regions** | | | | |
| **Pooled prevalence (95% CI)** | **15.0 (12.7, 17.3)** | **83.3 (80.7, 85.8)** | **1.7 (0.7, 2.7)** | **0.1 (0.0, 0.1)** |

All values are weighted to reflect the survey sampling design. Data are n (%). DK = Don't Know.
*The lower limit of the confidence interval was truncated to zero following a negative lower limit of the Wald-type confidence interval.
†Nepal (2019) survey individually measured menstrual-related absenteeism from a range of activities - absenteeism from school or work, and absenteeism from social gatherings/meetings were combined to generate the outcome variable. The Nepal (2019) survey also did not offer an option for 'DK or Not sure or No such activity' when measuring menstrual-related absenteeism.

**Table S12: Age profile of study surveys.**

| Country | Mean age (95% CI) |
|---|---|
| **West and Central Africa** | |
| Central African Republic | 28.0 (27.7, 28.2) |
| Chad | 27.3 (27.2, 27.5) |
| Democratic Republic of Congo | 27.4 (27.2, 27.7) |
| Gambia | 27.7 (27.5, 27.9) |
| Ghana | 29.4 (29.2, 29.6) |
| Guinea-Bissau | 28.2 (28.0, 28.4) |
| Nigeria | 28.7 (28.6, 28.9) |
| Sao Tome & Principe | 28.9 (28.6, 29.3) |
| Sierra Leone | 28.5 (28.3, 28.7) |
| Togo | 28.9 (28.6, 29.2) |
| **Pooled mean age (95% CI)** | **28.3 (27.9, 28.7)** |
| **Eastern and Southern Africa** | |
| Lesotho | 28.7 (28.4, 29.0) |
| Madagascar | 28.0 (27.8, 28.2) |
| Malawi | 27.9 (27.8, 28.1) |
| Zimbabwe | 29.3 (29.1, 29.5) |
| **Pooled mean age (95% CI)** | **28.5 (27.8, 29.1)** |
| **Middle East and North Africa** | |
| Algeria | 31.1 (30.9, 31.2) |
| Iraq | 32.7 (32.5, 32.9) |
| State of Palestine | 33.0 (32.8, 33.2) |
| Tunisia | 37.2 (36.9, 37.4) |
| **Pooled mean age (95% CI)** | **33.5 (30.9, 36.0)** |
| **South Asia** | |
| Afghanistan | 26.3 (26.2, 26.4) |
| Bangladesh | 29.1 (29.0, 29.2) |
| Nepal | 29.4 (29.2, 29.6) |
| Pakistan (Balochistan) | 27.9 (27.7, 28.0) |
| Pakistan (Khyber Pakhtunkhwa) | 28.0 (27.9, 28.1) |
| Pakistan (Sindh) | 28.2 (28.1, 28.3) |
| Pakistan (Punjab) | 28.5 (28.4, 28.5) |
| **Pooled mean age (95% CI)** | **28.2 (27.4, 28.9)** |
| **East Asia and the Pacific** | |
| Fiji | 31.0 (30.7, 31.3) |
| Kiribati | 29.1 (28.8, 29.3) |
| Lao PDR | 29.5 (29.4, 29.7) |
| Mongolia | 32.2 (31.9, 32.4) |
| Samoa | 29.7 (29.3, 30.0) |
| Tonga | 29.1 (28.6, 29.5) |
| Tuvalu | 29.3 (28.6, 30.0) |
| Vietnam | 31.8 (31.5, 32.0) |
| **Pooled mean age (95% CI)** | **30.2 (29.3, 31.1)** |
| **Europe and Central Asia** | |
| Kosovo | 30.7 (30.5, 31.0) |
| Kyrgyzstan | 30.8 (30.6, 31.1) |
| Montenegro | 32.6 (32.0, 33.3) |
| Republic of North Macedonia | 32.4 (31.9, 32.9) |
| Serbia | 33.5 (33.1, 33.8) |
| Turkmenistan | 33.8 (33.5, 34.0) |
| Uzbekistan | 31.0 (30.8, 31.3) |
| **Pooled mean age (95% CI)** | **32.1 (31.2, 33.1)** |
| **Latin America and Caribbean** | |
| Costa Rica | 30.9 (30.6, 31.3) |
| Cuba | 32.9 (32.6, 33.3) |
| Dominican Republic | 29.7 (29.5, 29.9) |
| Guyana | 30.1 (29.8, 30.3) |
| Honduras | 29.5 (29.3, 29.6) |
| Suriname | 30.0 (29.7, 30.4) |
| Turks & Caicos Islands | 33.6 (32.8, 34.3) |
| **Pooled mean age (95% CI)** | **30.9 (29.7, 32.1)** |
| **All regions** | |
| **Pooled mean age (95% CI)** | **30.0 (29.4, 30.6)** |

All values are weighted to reflect the survey sampling design.

**Table S13: Prevalence of urban and rural living in study participants.**

| Country | Urban | Rural |
|---|---|---|
| **West and Central Africa** | | |
| Central African Republic | 2802/7093 (39.5) | 4290/7093 (60.5) |
| Chad | 3893/18211 (21.4) | 14318/18211 (78.6) |
| Democratic Republic of Congo | 8959/16987 (52.7) | 8028/16987 (47.3) |
| Gambia | 8843/12177 (72.6) | 3334/12177 (27.4) |
| Ghana | 6636/12855 (51.6) | 6219/12855 (48.4) |
| Guinea-Bissau | 4464/10913 (40.9) | 6449/10913 (59.1) |
| Nigeria | 15993/33195 (48.2) | 17202/33195 (51.8) |
| Sao Tome & Principe | 1928/2858 (67.4) | 930/2858 (32.6) |
| Sierra Leone | 6922/13700 (50.5) | 6778/13700 (49.5) |
| Togo | 3062/6080 (50.4) | 3018/6080 (49.6) |
| **Pooled prevalence (95% CI)** | **49.5 (40.6, 58.4)** | **50.5 (41.6, 59.4)** |
| **Eastern and Southern Africa** | | |
| Lesotho | 2738/5648 (48.5) | 2909/5648 (51.5) |
| Madagascar | 3951/14060 (28.1) | 10109/14060 (71.9) |
| Malawi | 3989/20498 (19.5) | 16509/20498 (80.5) |
| Zimbabwe | 3470/8543 (40.6) | 5072/8543 (59.4) |
| **Pooled prevalence (95% CI)** | **34.1 (21.5, 46.8)** | **65.9 (53.2, 78.5)** |
| **Middle East and North Africa** | | |
| Algeria | 21058/33078 (63.7) | 12020/33078 (36.3) |
| Iraq | 13961/19733 (70.7) | 5773/19733 (29.3) |
| State of Palestine | 4974/6425 (77.4) | 948/6425 (14.7) |
| Tunisia | 3937/5668 (69.5) | 1731/5668 (30.5) |
| **Pooled prevalence (95% CI)** | **70.4 (64.7, 76.0)** | **27.7 (18.6, 36.7)** |
| **South Asia** | | |
| Afghanistan | 11249/40257 (27.9) | 29007/40257 (72.1) |
| Bangladesh | 13742/58198 (23.6) | 44456/58198 (76.4) |
| Nepal | 9393/13446 (69.9) | 4052/13446 (30.1) |
| Pakistan (Balochistan) | 8638/32395 (26.7) | 23756/32395 (73.3) |
| Pakistan (Khyber Pakhtunkhwa) | 6352/37504 (16.9) | 31152/37504 (83.1) |
| Pakistan (Sindh) | 15102/27398 (55.1) | 12296/27398 (44.9) |
| Pakistan (Punjab) | 26508/68491 (38.7) | 41983/68491 (61.3) |
| **Pooled prevalence (95% CI)** | **37.0 (22.9, 51.1)** | **63.0 (48.9, 77.1)** |
| **East Asia and the Pacific** | | |
| Fiji | 2942/4726 (62.3) | 1784/4726 (37.7) |
| Kiribati | 2080/3519 (59.1) | 1439/3519 (40.9) |
| Lao PDR* | 7896/22346 (35.3) | 14450/22346 (64.7) |
| Mongolia | 6604/9489 (69.6) | 2885/9489 (30.4) |
| Samoa | 805/3858 (20.9) | 3053/3858 (79.1) |
| Tonga | 640/2678 (23.9) | 2038/2678 (76.1) |
| Tuvalu | 497/728 (68.2) | 231/728 (31.8) |
| Vietnam | 3836/10147 (37.8) | 6311/10147 (62.2) |
| **Pooled prevalence (95% CI)** | **47.1 (33.3, 60.9)** | **52.9 (39.1, 66.7)** |
| **Europe and Central Asia** | | |
| Kosovo | 2136/5020 (42.5) | 2885/5020 (57.5) |
| Kyrgyzstan | 2080/5175 (40.2) | 3096/5175 (59.8) |
| Montenegro | 1478/2156 (68.5) | 678/2156 (31.5) |
| Republic of North Macedonia | 1922/3023 (63.6) | 1100/3023 (36.4) |
| Serbia | 2215/3527 (62.8) | 1312/3527 (37.2) |
| Turkmenistan | 2240/4946 (45.3) | 2706/4946 (54.7) |
| Uzbekistan | 2066/4360 (47.4) | 2295/4360 (52.6) |
| **Pooled prevalence (95% CI)** | **52.8 (44.2, 61.4)** | **47.2 (38.6, 55.8)** |
| **Latin America and Caribbean** | | |
| Costa Rica | 4886/6795 (71.9) | 1909/6795 (28.1) |
| Cuba | 5395/8243 (65.5) | 2847/8243 (34.5) |
| Dominican Republic | 15878/20677 (76.8) | 4798/20677 (23.2) |
| Guyana | 1320/5411 (24.4) | 4090/5411 (75.6) |
| Honduras | 8664/17572 (49.3) | 8908/17572 (50.7) |
| Suriname | 4866/6441 (75.6) | 1099/6441 (17.1) |
| Turks & Caicos Islands† | 760/789 (96.2) | 30/789 (3.8) |
| **Pooled prevalence (95% CI)** | **65.7 (48.8, 82.7)** | **33.2 (15.7, 50.7)** |
| **All regions** | | |
| **Pooled prevalence (95% CI)** | **50.6 (45.1, 56.1)** | **49.1 (43.4, 54.7)** |

All values are weighted to reflect the survey sampling design. Data are n/N (%).
*Area response options in Lao PDR included urban, rural with road, and rural without road. These responses were recoded to urban, and rural (includes both rural with road and rural without road).
†The survey weighted total sample size for Turks and Caicos Islands is 789.3 women and girls, including 759.7 (rounded to 760) women and girls in urban areas and 29.6 (rounded to 30) women and girls in rural areas.

**Table S14: Prevalence of the use of menstrual materials in study participants.**

| Country | Used menstrual materials | Used reusable menstrual materials* |
|---|---|---|
| **West and Central Africa** | | |
| Central African Republic | 6745/7093 (95.1) | 4388/6745 (65.1) |
| Chad | 17293/18204 (95.0) | 14640/17293 (84.7) |
| Democratic Republic of Congo | 16066/16987 (94.6) | 9444/16066 (58.8) |
| Gambia | 11958/12177 (98.2) | 7077/11958 (59.2) |
| Ghana | 12583/12855 (97.9) | 1614/12583 (12.8) |
| Guinea-Bissau† | – | – |
| Nigeria | 32202/33195 (97.0) | 13702/32202 (42.6) |
| Sao Tome & Principe | 2845/2858 (99.6) | 2371/2441 (97.1) |
| Sierra Leone | 13305/13700 (97.1) | 9257/13305 (69.6) |
| Togo | 5859/6080 (96.4) | 3479/5859 (59.4) |
| **Pooled prevalence (95% CI)** | **96.8 (95.7, 97.9)** | **61.0 (45.3, 76.7)** |
| **Eastern and Southern Africa** | | |
| Lesotho | 5539/5648 (98.1) | 427/5539 (7.7) |
| Madagascar | 13186/14060 (93.8) | 10262/13186 (77.8) |
| Malawi | 19954/20498 (97.3) | 14039/19954 (70.4) |
| Zimbabwe | 8355/8543 (97.8) | 1845/8355 (22.1) |
| **Pooled prevalence (95% CI)** | **96.8 (94.9, 98.7)** | **44.5 (10.4, 78.6)** |
| **Middle East and North Africa** | | |
| Algeria | 31262/33074 (94.5) | 1569/31262 (5.0) |
| Iraq | 16110/16799 (95.9) | 1871/16110 (11.6) |
| State of Palestine | 6219/6425 (96.8) | 137/6219 (2.2) |
| Tunisia | 5440/5668 (96.0) | 207/5440 (3.8) |
| **Pooled prevalence (95% CI)** | **95.8 (94.9, 96.7)** | **5.6 (1.6, 9.6)** |
| **South Asia** | | |
| Afghanistan | 36921/40257 (91.7) | 30226/36921 (81.9) |
| Bangladesh | 56148/58198 (96.5) | 38504/56148 (68.6) |
| Nepal | 12627/13446 (93.9) | 7925/12627 (62.8) |
| Pakistan (Balochistan) | 20844/32395 (64.3) | 9750/20844 (46.8) |
| Pakistan (Khyber Pakhtunkhwa) | 35054/37504 (93.5) | 27935/35054 (79.7) |
| Pakistan (Sindh) | 22335/27398 (81.5) | 11363/22335 (50.9) |
| Pakistan (Punjab) | 60905/68491 (88.9) | 31425/60905 (51.6) |
| **Pooled prevalence (95% CI)** | **87.2 (78.9, 95.5)** | **63.2 (52.7, 73.7)** |
| **East Asia and the Pacific** | | |
| Fiji | 4594/4726 (97.2) | 556/4594 (12.1) |
| Kiribati | 3451/3519 (98.1) | 564/3451 (16.3) |
| Lao PDR | 18281/22346 (81.8) | 632/18281 (3.5) |
| Mongolia | 8669/9489 (91.4) | 245/8669 (2.8) |
| Samoa | 3541/3858 (91.8) | 693/3541 (19.6) |
| Tonga | 2522/2678 (94.2) | 25/2522 (1.0) |
| Tuvalu | 692/728 (95.0) | 128/692 (18.6) |
| Vietnam | 9959/10147 (98.2) | 120/9959 (1.2) |
| **Pooled prevalence (95% CI)** | **93.5 (89.7, 97.2)** | **9.2 (3.7, 14.8)** |
| **Europe and Central Asia** | | |
| Kosovo | 4979/5020 (99.2) | 162/4979 (3.3) |
| Kyrgyzstan | 5018/5175 (97.0) | 933/5018 (18.6) |
| Montenegro | 2092/2156 (97.0) | 87/2092 (4.1) |
| Republic of North Macedonia | 2980/3023 (98.6) | 26/2980 (0.9) |
| Serbia | 3469/3527 (98.4) | 18/3469 (0.5) |
| Turkmenistan | 4902/4946 (99.1) | 42/4902 (0.8) |
| Uzbekistan | 4209/4360 (96.5) | 624/4209 (14.8) |
| **Pooled prevalence (95% CI)** | **98.0 (97.2, 98.8)** | **6.1 (0.6, 11.6)** |
| **Latin America and Caribbean** | | |
| Costa Rica | 6710/6795 (98.7) | 131/6710 (2.0) |
| Cuba | 8048/8243 (97.6) | 216/8048 (2.7) |
| Dominican Republic | 20330/20677 (98.3) | 455/20330 (2.2) |
| Guyana | 5219/5411 (96.5) | 106/5219 (2.0) |
| Honduras | 17240/17572 (98.1) | 538/17240 (3.1) |
| Suriname | 5983/6441 (92.9) | 227/5983 (3.8) |
| Turks & Caicos Islands | 785/789 (99.4) | 9/785 (1.2) |
| **Pooled prevalence (95% CI)** | **97.4 (95.9, 99.0)** | **2.4 (1.8, 3.0)** |
| **All regions** | | |
| **Pooled prevalence (95% CI)** | **94.9 (93.2, 96.7)** | **28.8 (20.0, 37.7)** |

All values are weighted to reflect the survey sampling design. Data are n/N (%).
*Proportion of women and girls using menstrual materials who used *reusable* materials. Some women and girls who used menstrual materials had missing data for reusable menstrual materials and were therefore not included in the total sample size for reusable menstrual materials.
†Guinea-Bissau results were excluded as the MICS report did not consider estimates for the use of menstrual materials to reflect the reality of Guinea-Bissau due to an error that was detected when adapting the Computer-Assisted Personal Interviewing (CAPI) to tablets.

**Table S15: Prevalence of having a private place to wash at home in study participants.**

| Country | Had a private place to wash at home during menstruation |
|---|---|
| **West and Central Africa** | |
| Central African Republic | 6528/7093 (92.0) |
| Chad | 17007/18205 (93.4) |
| Democratic Republic of Congo | 15352/16987 (90.4) |
| Gambia | 11701/12177 (96.1) |
| Ghana | 12068/12855 (93.9) |
| Guinea-Bissau* | 720/909 (79.2) |
| Nigeria | 30815/33195 (92.8) |
| Sao Tome & Principe | 2695/2858 (94.3) |
| Sierra Leone | 12728/13700 (92.9) |
| Togo | 5565/6080 (91.5) |
| **Pooled prevalence (95% CI)** | **93.1 (92.0, 94.2)** |
| **Eastern and Southern Africa** | |
| Lesotho | 5349/5648 (94.7) |
| Madagascar | 12765/14060 (90.8) |
| Malawi | 18968/20498 (92.5) |
| Zimbabwe | 8253/8543 (96.6) |
| **Pooled prevalence (95% CI)** | **93.7 (91.2, 96.2)** |
| **Middle East and North Africa** | |
| Algeria | 29824/33074 (90.2) |
| Iraq | 14889/16799 (88.6) |
| State of Palestine | 5170/6425 (80.5) |
| Tunisia | 3185/5668 (56.2) |
| **Pooled prevalence (95% CI)** | **78.9 (63.5, 94.3)** |
| **South Asia** | |
| Afghanistan | 36857/40257 (91.6) |
| Bangladesh | 56287/58198 (96.7) |
| Nepal | 11638/13446 (86.6) |
| Pakistan (Balochistan) | 20954/32395 (64.7) |
| Pakistan (Khyber Pakhtunkhwa) | 33236/37504 (88.6) |
| Pakistan (Sindh) | 22902/27398 (83.6) |
| Pakistan (Punjab) | 61772/68491 (90.2) |
| **Pooled prevalence (95% CI)** | **86.0 (78.4, 93.6)** |
| **East Asia and the Pacific** | |
| Fiji | 4525/4726 (95.8) |
| Kiribati | 3266/3519 (92.8) |
| Lao PDR | 18091/22346 (81.0) |
| Mongolia | 8492/9489 (89.5) |
| Samoa | 3266/3858 (84.7) |
| Tonga | 2518/2678 (94.1) |
| Tuvalu | 687/728 (94.4) |
| Vietnam | 9848/10147 (97.1) |
| **Pooled prevalence (95% CI)** | **91.1 (87.2, 95.1)** |
| **Europe and Central Asia** | |
| Kosovo | 4943/5020 (98.5) |
| Kyrgyzstan | 4828/5175 (93.3) |
| Montenegro | 2094/2156 (97.1) |
| Republic of North Macedonia | 2952/3023 (97.7) |
| Serbia | 3489/3527 (98.9) |
| Turkmenistan | 4890/4946 (98.9) |
| Uzbekistan | 4216/4360 (96.7) |
| **Pooled prevalence (95% CI)** | **97.3 (95.9, 98.8)** |
| **Latin America and Caribbean** | |
| Costa Rica | 6724/6795 (99.0) |
| Cuba | 7869/8243 (95.5) |
| Dominican Republic | 19706/20677 (95.3) |
| Guyana | 5051/5411 (93.3) |
| Honduras | 16963/17572 (96.5) |
| Suriname | 6185/6441 (96.0) |
| Turks & Caicos Islands | 761/789 (96.5) |
| **Pooled prevalence (95% CI)** | **96.0 (94.8, 97.3)** |
| **All regions** | |
| **Pooled prevalence (95% CI)** | **91.6 (89.2, 93.9)** |

All values are weighted to reflect the survey sampling design. Data are n/N (%).
*Guinea-Bissau results were excluded as the MICS report did not consider estimates for the availability of a private place to wash at home during menstruation to reflect the reality of Guinea-Bissau due to an error that was detected when adapting the Computer-Assisted Personal Interviewing (CAPI) to tablets.

**Table S16: Prevalence of current contraceptive use in study participants.**

| Country | Currently using any method of contraception | Currently using hormonal contraception* |
|---|---|---|
| **West and Central Africa** | | |
| Central African Republic | 1201/6160 (19.5) | 686/1200 (57.1) |
| Chad | 1347/15525 (8.7) | 816/1334 (61.2) |
| Democratic Republic of Congo | 4540/14738 (30.8) | 1322/4479 (29.5) |
| Gambia | 1416/11103 (12.8) | 1298/1369 (94.8) |
| Ghana | 2772/11966 (23.2) | 2035/2671 (76.2) |
| Guinea-Bissau | 3378/10034 (33.7) | 1810/2841 (63.7) |
| Nigeria | 6168/30044 (20.5) | 3467/5872 (59.1) |
| Sao Tome & Principe | 1024/2681 (38.2) | 745/981 (76.0) |
| Sierra Leone | 3714/12485 (29.7) | 3512/3689 (95.2) |
| Togo | 1346/5604 (24.0) | 767/1307 (58.7) |
| **Pooled prevalence (95% CI)** | **24.1 (18.3, 29.8)** | **67.2 (55.2, 79.3)** |
| **Eastern and Southern Africa** | | |
| Lesotho | 2879/5444 (52.9) | 1852/2791 (66.4) |
| Madagascar | 5208/12897 (40.4) | 4385/5085 (86.2) |
| Malawi | 9766/18959 (51.5) | 7819/9598 (81.5) |
| Zimbabwe | – | – |
| **Pooled prevalence (95% CI)** | **48.3 (40.5, 56.0)** | **78.0 (66.4, 89.7)** |
| **Middle East and North Africa** | | |
| Algeria | 9907/15679 (63.2) | 7282/9456 (77.0) |
| Iraq | 10143/17725 (57.2) | 3873/8433 (45.9) |
| State of Palestine | 3788/5707 (66.4) | 510/2002 (25.5) |
| Tunisia | 2865/5320 (53.9) | 1155/1657 (69.7) |
| **Pooled prevalence (95% CI)** | **60.2 (54.6, 65.7)** | **54.5 (31.5, 77.5)** |
| **South Asia** | | |
| Afghanistan | – | – |
| Bangladesh | 30239/42727 (70.8) | 23538/29931 (78.6) |
| Nepal | 4750/9677 (49.1) | 2323/4509 (51.5) |
| Pakistan (Balochistan) | 4483/16437 (27.3) | 2227/4319 (51.6) |
| Pakistan (Khyber Pakhtunkhwa) | 8369/21315 (39.3) | 3986/8106 (49.2) |
| Pakistan (Sindh) | 3856/15486 (24.9) | 1499/3549 (42.3) |
| Pakistan (Punjab) | 14859/37511 (39.6) | 2053/13540 (15.2) |
| **Pooled prevalence (95% CI)** | **41.8 (28.4, 55.2)** | **48.0 (31.7, 64.4)** |
| **East Asia and the Pacific** | | |
| Fiji | 1125/4528 (24.9) | 629/1043 (60.3) |
| Kiribati | 827/3286 (25.2) | 460/812 (56.6) |
| Lao PDR | 9286/21266 (43.7) | 6891/8900 (77.4) |
| Mongolia | 3875/8925 (43.4) | 919/1723 (53.3) |
| Samoa | 387/3620 (10.7) | 265/368 (72.1) |
| Tonga | 449/2559 (17.6) | 210/427 (49.2) |
| Tuvalu | 127/679 (18.8) | 109/127 (85.4) |
| Vietnam | 5438/9868 (55.1) | 1358/3637 (37.3) |
| **Pooled prevalence (95% CI)** | **29.9 (19.1, 40.7)** | **61.3 (50.4, 72.3)** |
| **Europe and Central Asia** | | |
| Kosovo | 2144/4863 (44.1) | 84/2047 (4.1) |
| Kyrgyzstan | 1619/4795 (33.8) | 146/803 (18.1) |
| Montenegro | 380/2076 (18.3) | 32/338 (9.3) |
| Republic of North Macedonia | 1495/2910 (51.4) | 39/1461 (2.7) |
| Serbia | 1807/3446 (52.4) | 92/1748 (5.3) |
| Turkmenistan | 2481/4462 (55.6) | 65/282 (23.2) |
| Uzbekistan | 2001/4053 (49.4) | 97/513 (18.9) |
| **Pooled prevalence (95% CI)** | **43.6 (33.8, 53.4)** | **11.4 (5.3, 17.5)** |
| **Latin America and Caribbean** | | |
| Costa Rica | 3742/6570 (57.0) | 1950/3590 (54.3) |
| Cuba | 5738/8013 (71.6) | 1053/3912 (26.9) |
| Dominican Republic | 9902/19791 (50.0) | 4734/9435 (50.2) |
| Guyana | 1232/5195 (23.7) | 558/1044 (53.5) |
| Honduras | 8024/16767 (47.9) | 3523/7278 (48.4) |
| Suriname | 1922/6212 (30.9) | 1454/1841 (78.9) |
| Turks & Caicos Islands | 264/771 (34.2) | 130/255 (50.8) |
| **Pooled prevalence (95% CI)** | **45.1 (32.8, 57.4)** | **51.9 (40.6, 63.1)** |
| **All regions** | | |
| **Pooled prevalence (95% CI)** | **38.6 (33.8, 43.5)** | **52.2 (44.9, 59.5)** |

All values are weighted to reflect the survey sampling design. Data are n/N (%).

*Hormonal contraception use in women and girls using contraception was derived from reported use of contraceptive methods. Users of hormonal contraception include women and girls using injectables, implants or the pill. Women and girls using intra-uterine devices (IUD) were excluded. Some women and girls using contraception had missing data for the method of contraception used, and were therefore not included in the total sample size for hormonal contraception use.

**Table S17: Prevalence of menstrual-related absenteeism by age group in study participants.**

| Country | Prevalence of menstrual-related absenteeism | | | | | | |
|---|---|---|---|---|---|---|---|
| | 15-19 years | 20-24 years | 25-29 years | 30-34 years | 35-39 years | 40-44 years | 45-49 years |
| **West and Central Africa** | | | | | | | |
| Central African Republic | 537/1539 (34.9) | 448/1423 (31.5) | 352/1253 (28.1) | 299/1067 (28.0) | 278/869 (32.0) | 177/565 (31.4) | 118/376 (31.4) |
| Chad | 1614/4577 (35.3) | 1097/3332 (32.9) | 1002/3202 (31.3) | 817/2611 (31.3) | 714/2201 (32.5) | 448/1438 (31.2) | 250/849 (29.5) |
| Democratic Republic of Congo | 747/4486 (16.6) | 455/3123 (14.6) | 373/2647 (14.1) | 275/2437 (11.3) | 271/2008 (13.5) | 180/1498 (12.0) | 111/787 (14.1) |
| Gambia | 917/2819 (32.5) | 668/2494 (26.8) | 332/2011 (16.5) | 221/1731 (12.8) | 167/1505 (11.1) | 92/982 (9.4) | 66/636 (10.4) |
| Ghana | 606/2761 (22.0) | 431/2052 (21.0) | 355/1918 (18.5) | 377/1884 (20.0) | 234/1716 (13.6) | 247/1499 (16.5) | 176/1025 (17.2) |
| Guinea-Bissau | 242/2328 (10.4) | 222/2238 (9.9) | 153/1911 (8.0) | 110/1511 (7.3) | 91/1373 (6.6) | 49/910 (5.4) | 25/643 (3.8) |
| Nigeria | 1723/7700 (22.4) | 953/5478 (17.4) | 831/4891 (17.0) | 667/4467 (14.9) | 556/4492 (12.4) | 520/3660 (14.2) | 253/2507 (10.1) |
| Sao Tome & Principe | 114/676 (16.9) | 67/470 (14.2) | 28/388 (7.1) | 38/413 (9.1) | 24/401 (6.0) | 30/316 (9.7) | 13/195 (6.4) |
| Sierra Leone | 658/2818 (23.3) | 540/2661 (20.3) | 456/2395 (19.0) | 361/1899 (19.0) | 363/1790 (20.3) | 197/1198 (16.4) | 186/939 (19.8) |
| Togo | 213/1320 (16.1) | 140/994 (14.1) | 112/1006 (11.2) | 101/866 (11.6) | 73/813 (9.0) | 54/661 (8.1) | 43/419 (10.4) |
| **Pooled prevalence (95% CI)** | **23.0 (17.7, 28.3)** | **20.2 (15.4, 25.1)** | **17.1 (12.2, 21.9)** | **16.5 (11.5, 21.4)** | **15.6 (9.7, 21.5)** | **15.3 (9.7, 20.9)** | **15.1 (9.5, 20.8)** |
| **Eastern and Southern Africa** | | | | | | | |
| Lesotho | 184/1231 (14.9) | 166/1030 (16.1) | 108/883 (12.3) | 90/876 (10.3) | 75/686 (10.9) | 64/582 (11.0) | 55/361 (15.2) |
| Madagascar | 365/3616 (10.1) | 235/2625 (8.9) | 160/2145 (7.4) | 134/1688 (7.9) | 108/1631 (6.6) | 107/1456 (7.3) | 60/899 (6.6) |
| Malawi | 1000/5026 (19.9) | 476/3980 (11.9) | 337/3119 (10.8) | 259/2756 (9.4) | 234/2576 (9.1) | 201/1835 (11.0) | 94/1208 (7.8) |
| Zimbabwe | 397/1818 (21.8) | 262/1396 (18.8) | 183/1215 (15.0) | 177/1281 (13.8) | 153/1264 (12.1) | 130/962 (13.5) | 92/608 (15.1) |
| **Pooled prevalence (95% CI)** | **16.7 (11.5, 21.8)** | **13.8 (9.6, 18.1)** | **11.3 (8.2, 14.4)** | **10.3 (7.9, 12.7)** | **9.6 (7.2, 11.9)** | **10.6 (8.1, 13.2)** | **10.9 (6.5, 15.4)** |
| **Middle East and North Africa** | | | | | | | |
| Algeria | 1595/4722 (33.8) | 1582/5092 (31.1) | 1293/5323 (24.3) | 1107/5203 (21.3) | 1034/5018 (20.6) | 887/4495 (19.7) | 530/3224 (16.4) |
| Iraq | 93/1205 (7.8) | 275/2881 (9.5) | 326/3580 (9.1) | 438/3597 (12.2) | 355/3486 (10.2) | 374/2864 (13.0) | 235/2120 (11.1) |
| State of Palestine | 25/168 (14.8) | 138/989 (13.9) | 177/1350 (13.1) | 168/1200 (14.0) | 139/1086 (12.8) | 138/928 (14.8) | 110/704 (15.6) |
| Tunisia | 1/9 (13.2) | 28/202 (14.0) | 69/623 (11.1) | 133/1240 (10.7) | 149/1319 (11.3) | 141/1308 (10.8) | 101/968 (10.4) |
| **Pooled prevalence (95% CI)** | **18.1 (6.3, 29.9)** | **17.2 (7.8, 26.6)** | **14.4 (7.7, 21.1)** | **14.6 (10.0, 19.1)** | **13.8 (9.1, 18.4)** | **14.6 (10.9, 18.4)** | **13.3 (10.3, 16.3)** |
| **South Asia** | | | | | | | |
| Afghanistan | 3767/11568 (32.6) | 2643/8713 (30.3) | 2023/6920 (29.2) | 1218/4488 (27.1) | 1165/3852 (30.2) | 803/2802 (28.7) | 544/1913 (28.5) |
| Bangladesh | 1133/11654 (9.7) | 785/9740 (8.1) | 710/9371 (7.6) | 702/9535 (7.4) | 593/8495 (7.0) | 425/5803 (7.3) | 226/3601 (6.3) |
| Nepal | 216/2582 (8.4) | 215/2332 (9.2) | 185/2183 (8.5) | 193/1974 (9.8) | 181/1847 (9.8) | 170/1509 (11.2) | 105/1018 (10.3) |
| Pakistan (Balochistan) | 1403/7094 (19.8) | 1433/6233 (23.0) | 1329/6264 (21.2) | 796/4764 (16.7) | 611/3647 (16.8) | 401/2403 (16.7) | 383/1990 (19.2) |
| Pakistan (Khyber Pakhtunkhwa) | 1761/8521 (20.7) | 1160/7009 (16.6) | 1112/6819 (16.3) | 760/5424 (14.0) | 612/4581 (13.4) | 453/3098 (14.6) | 232/2051 (11.3) |
| Pakistan (Sindh) | 2372/6059 (39.1) | 2019/5022 (40.2) | 1805/4709 (38.3) | 1624/4227 (38.4) | 1242/3507 (35.4) | 802/2338 (34.3) | 540/1536 (35.2) |
| Pakistan (Punjab) | 2679/14261 (18.8) | 2255/13032 (17.3) | 1838/11810 (15.6) | 1551/9777 (15.9) | 1442/9043 (15.9) | 970/6317 (15.4) | 654/4250 (15.4) |
| **Pooled prevalence (95% CI)** | **21.3 (13.0, 29.6)** | **20.6 (12.1, 29.2)** | **19.5 (11.3, 27.7)** | **18.4 (10.4, 26.5)** | **18.3 (10.5, 26.1)** | **18.3 (11.1, 25.4)** | **18.0 (10.2, 25.7)** |
| **East Asia and the Pacific** | | | | | | | |
| Fiji | 175/777 (22.6) | 150/666 (22.5) | 171/738 (23.1) | 148/685 (21.6) | 167/693 (24.1) | 158/687 (23.0) | 125/481 (25.9) |
| Kiribati | 93/595 (15.6) | 117/706 (16.6) | 96/646 (14.9) | 83/534 (15.5) | 83/470 (17.7) | 51/342 (15.0) | 42/225 (18.7) |
| Lao PDR | 603/4372 (13.8) | 484/3682 (13.2) | 417/3575 (11.7) | 421/3376 (12.5) | 274/2977 (9.2) | 260/2633 (9.9) | 185/1730 (10.7) |
| Mongolia | 59/1173 (5.0) | 37/1044 (3.6) | 42/1550 (2.7) | 49/1722 (2.8) | 39/1461 (2.7) | 46/1453 (3.1) | 30/1088 (2.8) |
| Samoa | 72/795 (9.1) | 73/699 (10.4) | 47/577 (8.1) | 32/478 (6.7) | 45/432 (10.3) | 39/481 (8.1) | 39/396 (9.8) |
| Tonga | 103/635 (16.3) | 74/435 (17.1) | 67/388 (17.3) | 36/353 (10.3) | 41/335 (12.3) | 62/309 (20.0) | 33/224 (14.8) |
| Tuvalu | 15/101 (14.5) | 31/151 (20.6) | 17/151 (11.2) | 15/112 (13.6) | 18/102 (17.3) | 6/62 (10.1) | 12/48 (24.9) |
| Vietnam | 77/1354 (5.7) | 75/1276 (5.9) | 83/1684 (4.9) | 56/1633 (3.4) | 30/1594 (1.9) | 43/1458 (3.0) | 39/1148 (3.4) |
| **Pooled prevalence (95% CI)** | **12.7 (8.5, 16.8)** | **13.4 (8.7, 18.0)** | **11.6 (7.0, 16.2)** | **10.6 (6.2, 15.1)** | **11.7 (6.4, 17.0)** | **11.2 (6.2, 16.3)** | **13.0 (7.0, 19.1)** |
| **Europe and Central Asia** | | | | | | | |
| Kosovo | 132/968 (13.7) | 89/767 (11.6) | 58/711 (8.2) | 53/618 (8.6) | 64/623 (10.3) | 65/730 (8.9) | 63/603 (10.5) |
| Kyrgyzstan | 97/809 (11.9) | 77/763 (10.0) | 44/848 (5.2) | 36/813 (4.5) | 36/699 (5.2) | 36/713 (5.1) | 36/531 (6.8) |
| Montenegro | 33/280 (11.7) | 13/277 (4.7) | 30/283 (10.5) | 14/315 (4.4) | 20/354 (5.5) | 19/364 (5.1) | 18/284 (6.4) |
| Republic of North Macedonia | 31/372 (8.4) | 30/417 (7.1) | 35/435 (8.0) | 26/438 (6.0) | 18/454 (3.9) | 32/492 (6.6) | 28/415 (6.6) |
| Serbia | 53/380 (14.0) | 50/434 (11.5) | 42/424 (9.9) | 38/537 (7.1) | 44/608 (7.2) | 43/569 (7.5) | 54/574 (9.3) |

| | | | | | | | |
|---|---|---|---|---|---|---|---|
| Turkmenistan | 0/42 (0.6)* | 7/514 (1.3) | 9/1093 (0.9) | 7/1110 (0.6) | 5/916 (0.6) | 6/801 (0.7) | 9/469 (1.9) |
| Uzbekistan | 53/628 (8.4) | 31/600 (5.2) | 45/722 (6.2) | 53/778 (6.9) | 60/691 (8.7) | 41/552 (7.4) | 31/389 (8.1) |
| **Pooled prevalence (95% CI)** | **9.6 (6.0, 13.1)** | **7.2 (4.3, 10.1)** | **6.5 (4.0, 9.0)** | **5.3 (3.3, 7.3)** | **5.8 (3.3, 8.2)** | **5.8 (3.7, 7.8)** | **7.0 (4.7, 9.2)** |
| **Latin America and Caribbean** | | | | | | | |
| Costa Rica | 70/974 (7.2) | 77/1126 (6.9) | 63/1087 (5.8) | 67/1085 (6.2) | 71/985 (7.2) | 59/838 (7.0) | 55/700 (7.9) |
| Cuba | 301/1024 (29.4) | 252/987 (25.5) | 339/1177 (28.8) | 345/1339 (25.8) | 302/1049 (28.8) | 299/1244 (24.0) | 433/1422 (30.4) |
| Dominican Republic | 907/3680 (24.6) | 800/3652 (21.9) | 766/3521 (21.8) | 588/2982 (19.7) | 589/2785 (21.1) | 525/2357 (22.3) | 348/1699 (20.4) |
| Guyana | 207/973 (21.3) | 207/1021 (20.2) | 161/893 (18.0) | 122/644 (18.9) | 124/572 (21.8) | 150/707 (21.1) | 125/602 (20.8) |
| Honduras | 722/3524 (20.5) | 595/3075 (19.3) | 508/2781 (18.2) | 481/2400 (20.0) | 398/2211 (18.0) | 419/2126 (19.7) | 261/1454 (17.9) |
| Suriname | 245/1319 (18.6) | 181/965 (18.8) | 153/916 (16.7) | 163/942 (17.3) | 163/885 (18.5) | 107/749 (14.2) | 114/665 (17.1) |
| Turks & Caicos Islands | 14/54 (25.2) | 20/110 (18.3) | 14/92 (15.0) | 18/137 (13.3) | 20/179 (11.2) | 7/118 (5.7) | 8/99 (8.1) |
| **Pooled prevalence (95% CI)** | **20.4 (15.0, 25.7)** | **18.6 (14.2, 23.1)** | **17.7 (12.5, 23.0)** | **17.4 (12.8, 22.0)** | **18.1 (12.8, 23.3)** | **16.4 (10.9, 21.8)** | **17.5 (11.7, 23.2)** |
| **All regions** | | | | | | | |
| **Pooled prevalence (95% CI)** | **17.7 (15.1, 20.3)** | **16.2 (13.8, 18.6)** | **14.4 (12.1, 16.7)** | **13.6 (11.3, 15.8)** | **13.6 (11.2, 16.0)** | **13.4 (11.1, 15.6)** | **13.8 (11.5, 16.2)** |

All values are weighted to reflect the survey sampling design. Data are n/N (%).

*After applying survey weights, 0.26 (rounded to 0) women aged 15-19 years in Turkmenistan experienced menstrual-related absenteeism, representing 0.6% of the sample.

**Table S18: Prevalence of menstrual-related absenteeism by wealth quintile in study participants.**

| Country | Prevalence of menstrual-related absenteeism | | | | |
|---|---|---|---|---|---|
| | Quintile 1 (Poorest) | Quintile 2 | Quintile 3 | Quintile 4 | Quintile 5 (Richest) |
| **West and Central Africa** | | | | | |
| Central African Republic | 460/1327 (34.7) | 480/1334 (36.0) | 425/1339 (31.8) | 412/1419 (29.1) | 431/1674 (25.7) |
| Chad | 1107/3316 (33.4) | 1107/3410 (32.5) | 1168/3573 (32.7) | 1312/3688 (35.6) | 1249/4224 (29.6) |
| Democratic Republic of Congo | 425/2741 (15.5) | 438/2901 (15.1) | 453/2984 (15.2) | 511/3667 (13.9) | 585/4695 (12.5) |
| Gambia | 333/2004 (16.6) | 362/2081 (17.4) | 455/2364 (19.2) | 584/2651 (22.0) | 730/3076 (23.7) |
| Ghana | 371/2045 (18.2) | 546/2323 (23.5) | 502/2608 (19.2) | 491/2778 (17.7) | 516/3101 (16.7) |
| Guinea-Bissau | 151/1915 (7.9) | 155/1989 (7.8) | 136/2076 (6.5) | 197/2299 (8.6) | 252/2635 (9.6) |
| Nigeria | 869/5357 (16.2) | 936/5799 (16.1) | 1166/6481 (18.0) | 1252/7420 (16.9) | 1279/8139 (15.7) |
| Sao Tome & Principe | 53/513 (10.3) | 48/535 (9.0) | 57/546 (10.4) | 76/619 (12.2) | 79/645 (12.3) |
| Sierra Leone | 575/2406 (23.9) | 455/2418 (18.8) | 475/2508 (18.9) | 588/2812 (20.9) | 668/3557 (18.8) |
| Togo | 112/856 (13.1) | 130/1000 (13.0) | 148/1142 (12.9) | 169/1450 (11.7) | 178/1630 (10.9) |
| **Pooled prevalence (95% CI)** | **18.9 (13.3, 24.4)** | **18.8 (13.1, 24.5)** | **18.4 (13.3, 23.6)** | **18.8 (13.6, 24.0)** | **17.5 (13.3, 21.7)** |
| **Eastern and Southern Africa** | | | | | |
| Lesotho | 105/782 (13.4) | 112/886 (12.7) | 144/1109 (13.0) | 168/1250 (13.4) | 213/1621 (13.1) |
| Madagascar | 159/2267 (7.0) | 173/2409 (7.2) | 184/2595 (7.1) | 228/3069 (7.4) | 424/3721 (11.4) |
| Malawi | 469/3864 (12.1) | 481/3750 (12.8) | 497/3821 (13.0) | 540/4090 (13.2) | 613/4973 (12.3) |
| Zimbabwe | 242/1338 (18.1) | 252/1416 (17.8) | 242/1483 (16.3) | 296/1972 (15.0) | 360/2333 (15.4) |
| **Pooled prevalence (95% CI)** | **12.5 (8.1, 17.0)** | **12.6 (8.3, 16.8)** | **12.3 (8.5, 16.1)** | **12.2 (8.9, 15.5)** | **13.0 (11.3, 14.7)** |
| **Middle East and North Africa** | | | | | |
| Algeria | 1360/6412 (21.2) | 1551/6439 (24.1) | 1616/6604 (24.5) | 1698/6679 (25.4) | 1802/6945 (25.9) |
| Iraq | 531/3711 (14.3) | 421/4056 (10.4) | 395/3969 (9.9) | 321/4043 (7.9) | 426/3955 (10.8) |
| State of Palestine | 196/1259 (15.6) | 179/1208 (14.9) | 172/1228 (14.0) | 181/1392 (13.0) | 165/1337 (12.3) |
| Tunisia | 126/943 (13.3) | 117/1151 (10.2) | 122/1139 (10.7) | 155/1225 (12.7) | 103/1210 (8.5) |
| **Pooled prevalence (95% CI)** | **16.2 (12.8, 19.7)** | **14.9 (8.5, 21.2)** | **14.8 (8.2, 21.3)** | **14.8 (7.4, 22.1)** | **14.4 (6.7, 22.1)** |
| **South Asia** | | | | | |
| Afghanistan | 2099/6858 (30.6) | 2181/7427 (29.4) | 2298/7932 (29.0) | 2562/8690 (29.5) | 3024/9349 (32.3) |
| Bangladesh | 867/10098 (8.6) | 811/10953 (7.4) | 981/11727 (8.4) | 964/12377 (7.8) | 951/13044 (7.3) |
| Nepal | 472/2294 (20.6) | 222/2525 (8.8) | 208/2629 (7.9) | 216/2883 (7.5) | 146/3114 (4.7) |
| Pakistan (Balochistan) | 1284/5927 (21.7) | 1683/6440 (26.1) | 1691/6603 (25.6) | 950/6413 (14.8) | 747/7011 (10.7) |
| Pakistan (Khyber Pakhtunkhwa) | 911/6830 (13.3) | 961/7160 (13.4) | 1283/7537 (17.0) | 1452/7832 (18.5) | 1483/8144 (18.2) |
| Pakistan (Sindh) | 2185/4684 (46.7) | 2448/5037 (48.6) | 2553/5606 (45.5) | 1897/5986 (31.7) | 1321/6086 (21.7) |
| Pakistan (Punjab) | 1603/11403 (14.1) | 2015/13168 (15.3) | 2435/14005 (17.4) | 2667/14632 (18.2) | 2669/15282 (17.5) |
| **Pooled prevalence (95% CI)** | **22.1 (12.6, 31.7)** | **21.3 (10.4, 32.1)** | **21.5 (11.7, 31.3)** | **18.3 (11.2, 25.3)** | **16.0 (9.0, 23.0)** |
| **East Asia and the Pacific** | | | | | |
| Fiji | 214/789 (27.1) | 210/924 (22.7) | 215/957 (22.4) | 212/987 (21.5) | 243/1069 (22.7) |
| Kiribati | 88/604 (14.6) | 98/627 (15.7) | 131/690 (19.0) | 130/773 (16.8) | 118/824 (14.3) |
| Lao PDR | 378/3580 (10.6) | 583/4026 (14.5) | 516/4303 (12.0) | 549/4900 (11.2) | 620/5537 (11.2) |
| Mongolia | 76/1749 (4.4) | 44/1739 (2.5) | 68/1906 (3.5) | 46/1984 (2.3) | 70/2111 (3.3) |
| Samoa | 73/743 (9.8) | 74/744 (10.0) | 57/765 (7.4) | 57/775 (7.4) | 84/831 (10.2) |
| Tonga | 96/503 (19.2) | 78/539 (14.4) | 91/556 (16.3) | 78/546 (14.2) | 75/533 (14.0) |
| Tuvalu | 21/132 (16.2) | 26/142 (18.4) | 25/143 (17.5) | 20/148 (13.3) | 22/164 (13.2) |
| Vietnam | 90/1795 (5.0) | 92/2008 (4.6) | 82/2110 (3.9) | 67/2055 (3.2) | 73/2179 (3.3) |
| **Pooled prevalence (95% CI)** | **13.1 (7.9, 18.4)** | **12.6 (7.9, 17.3)** | **12.5 (7.5, 17.5)** | **11.0 (6.4, 15.7)** | **11.3 (6.9, 15.8)** |
| **Europe and Central Asia** | | | | | |
| Kosovo | 129/941 (13.7) | 106/998 (10.6) | 92/1041 (8.9) | 91/1007 (9.0) | 108/1034 (10.5) |
| Kyrgyzstan | 39/975 (4.0) | 48/970 (5.0) | 81/1006 (8.1) | 70/1011 (6.9) | 124/1214 (10.2) |
| Montenegro | 24/356 (6.8) | 28/355 (7.9) | 13/416 (3.1) | 38/515 (7.3) | 43/514 (8.3) |
| Republic of North Macedonia | 65/526 (12.4) | 42/587 (7.1) | 41/584 (7.0) | 30/646 (4.7) | 22/679 (3.2) |
| Serbia | 57/460 (12.4) | 59/647 (9.1) | 66/753 (8.8) | 71/809 (8.8) | 70/857 (8.2) |
| Turkmenistan | 17/952 (1.8) | 5/956 (0.6) | 4/931 (0.4) | 5/985 (0.5) | 12/1121 (1.0) |
| Uzbekistan | 107/894 (12.0) | 69/858 (8.1) | 48/848 (5.6) | 50/858 (5.8) | 40/903 (4.5) |
| **Pooled prevalence (95% CI)** | **8.8 (5.2, 12.4)** | **6.7 (4.2, 9.3)** | **5.8 (3.4, 8.2)** | **6.0 (3.8, 8.3)** | **6.4 (3.6, 9.2)** |
| **Latin America and Caribbean** | | | | | |
| Costa Rica | 109/1245 (8.8) | 114/1351 (8.5) | 105/1447 (7.3) | 70/1389 (5.0) | 63/1363 (4.6) |
| Cuba | 398/1524 (26.1) | 409/1524 (26.8) | 494/1607 (30.8) | 495/1707 (29.0) | 474/1881 (25.2) |
| Dominican Republic | 710/3379 (21.0) | 952/4084 (23.3) | 1030/4283 (24.1) | 975/4617 (21.1) | 855/4313 (19.8) |
| Guyana | 192/865 (22.2) | 191/1008 (18.9) | 258/1150 (22.4) | 236/1233 (19.2) | 218/1154 (18.9) |
| Honduras | 628/2749 (22.8) | 661/3286 (20.1) | 698/3607 (19.3) | 732/4030 (18.2) | 664/3900 (17.0) |
| Suriname | 192/1182 (16.3) | 225/1302 (17.3) | 253/1363 (18.5) | 235/1337 (17.6) | 221/1256 (17.6) |
| Turks & Caicos Islands | 12/146 (8.0) | 19/165 (11.3) | 18/160 (11.5) | 24/174 (14.0) | 27/144 (19.0) |
| **Pooled prevalence (95% CI)** | **18.0 (12.8, 23.1)** | **18.1 (13.4, 22.8)** | **19.3 (13.5, 25.0)** | **17.7 (12.3, 23.1)** | **17.2 (12.5, 22.0)** |
| **All regions** | | | | | |
| **Pooled prevalence (95% CI)** | **16.0 (13.5, 18.5)** | **15.4 (12.8, 18.0)** | **15.3 (12.7, 17.9)** | **14.4 (12.1, 16.8)** | **13.9 (11.8, 16.0)** |

All values are weighted to reflect the survey sampling design. Data are n/N (%).

**Figure S1: Conceptual model showing relationships between the variables of interest.**

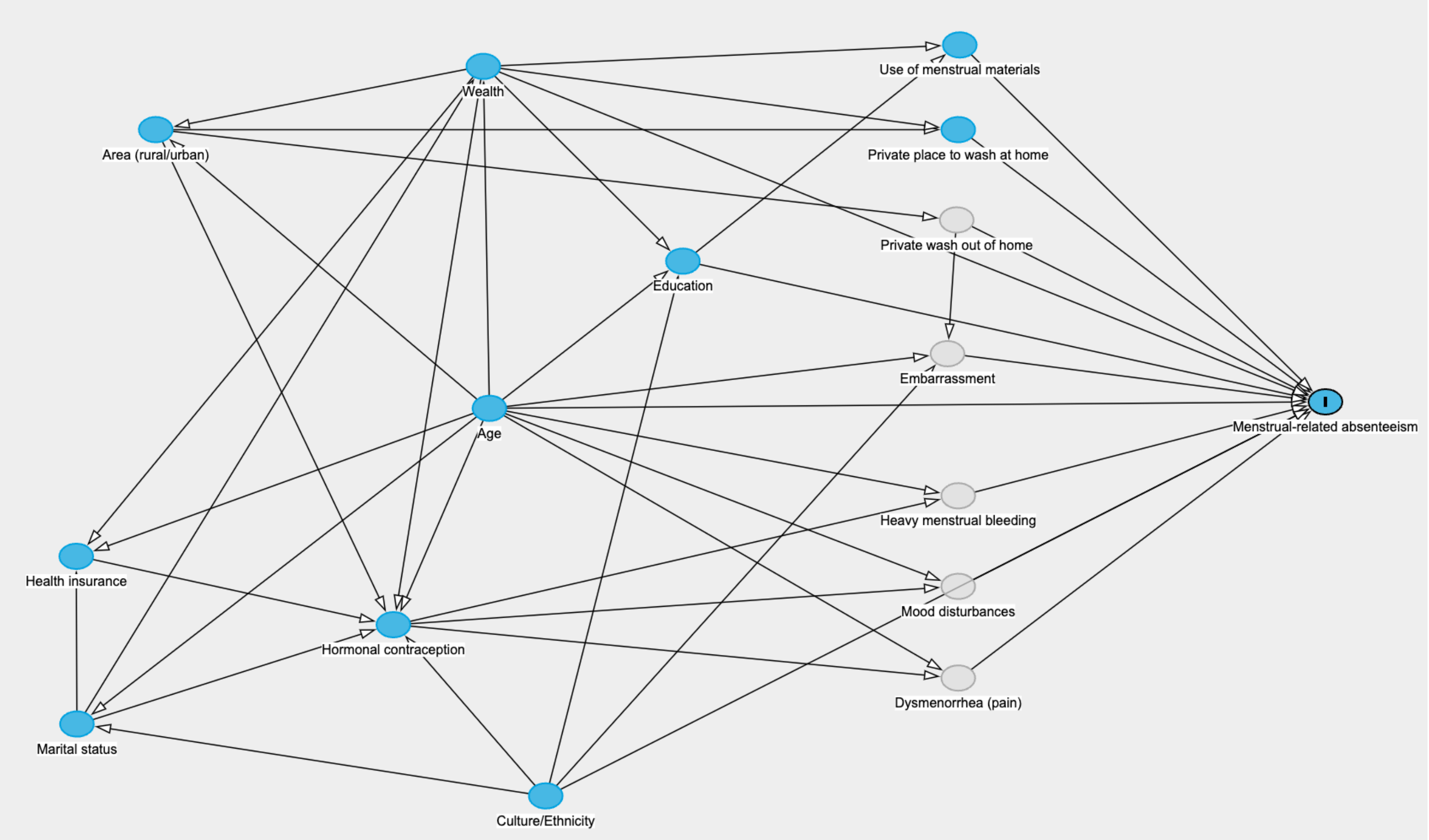

Conceptual representation of the relationships between variables of interest and menstrual-related absenteeism. Unmeasured variables are shown in grey, while all other variables are displayed in blue. The outcome variable (menstrual-related absenteeism) is additionally denoted with an 'I'.

This figure was created using Daggity (Johannes Textor, Benito van der Zander, Mark K. Gilthorpe, Maciej Liskiewicz, George T.H. Ellison. Robust causal inference using directed acyclic graphs: the R package 'dagitty'. *International Journal of Epidemiology* 45(6):1887-1894, 2016.)

The above figure was used to generate directed acyclic graphs (DAGs) to identify confounders to be adjusted for in multivariable models. In the Daggity program, our independent variables of interest were changed to represent the 'exposure' one at a time, and the minimal sufficient adjustment sets were noted for each association (see table below). No confounders for adjustment were identified from the DAG for the association between age and menstrual-related absenteeism. However, area type (urban/rural) was adjusted for in this analysis due to its potential to determine life-expectancy and contribute to menstrual-related absenteeism. For the association between contraception (any method, hormonal method) and menstrual-related absenteeism, culture and ethnicity were identified as a confounder. However, these characteristics were highly variable and difficult to categorise in a standardised way across MICS surveys. We also believed ethnicity and culture to be more important between survey countries rather than within them, and therefore this factor was not adjusted for in our analyses.

| Independent variable(s) | Minimum adjustment set |
|---|---|
| Age (categorical) | Area (urban/rural) |
| Area | Age (continuous)<br>Wealth (quintiles) |
| Wealth | Age (continuous)<br>Marital status (Currently married or in union/Formerly married or in union/Never married or in union) |
| Use of menstrual materials, and use of reusable menstrual materials | Wealth<br>Education (Early childhood education/Primary/Lower secondary/Upper secondary/Higher)<br>Area |
| Availability of a private place to wash at home during menstruation | Wealth<br>Area |
| Use of any contraception, and use of hormonal contraception | Age (continuous)<br>Wealth<br>Education<br>Area |

**Table S19: Univariable analysis between age group and menstrual-related absenteeism.**

| Country | Age group (years) | | | | | | | | | | | | |
|---|---|---|---|---|---|---|---|---|---|---|---|---|---|
| | 15-19 | 20-24 | | 25-29 | | 30-34 | | 35-39 | | 40-44 | | 45-49 | |
| | | PR (95% CI) | P value | PR (95% CI) | P value | PR (95% CI) | P value | PR (95% CI) | P value | PR (95% CI) | P value | PR (95% CI) | P value |
| **West and Central Africa** | | | | | | | | | | | | | |
| Central African Republic | | 0.90 (0.81, 1.02) | 0.090 | 0.81 (0.71, 0.92) | 0.0013 | 0.81 (0.71, 0.92) | 0.0017 | 0.92 (0.80, 1.04) | 0.18 | 0.90 (0.77, 1.05) | 0.19 | 0.90 (0.74, 1.10) | 0.31 |
| Chad | | 0.94 (0.87, 1.01) | 0.11 | 0.89 (0.81, 0.97) | 0.0072 | 0.89 (0.81, 0.97) | 0.0094 | 0.92 (0.84, 1.01) | 0.084 | 0.88 (0.78, 1.00) | 0.044 | 0.84 (0.73, 0.96) | 0.0084 |
| Democratic Republic of Congo | | 0.88 (0.73, 1.05) | 0.16 | 0.85 (0.71, 1.01) | 0.066 | 0.68 (0.54, 0.84) | 0.0005 | 0.81 (0.64, 1.03) | 0.080 | 0.73 (0.56, 0.93) | 0.013 | 0.85 (0.64, 1.12) | 0.24 |
| Gambia | | 0.82 (0.73, 0.93) | 0.0013 | 0.51 (0.44, 0.59) | <0.0001 | 0.39 (0.33, 0.46) | <0.0001 | 0.34 (0.29, 0.41) | <0.0001 | 0.29 (0.23, 0.37) | <0.0001 | 0.32 (0.24, 0.44) | <0.0001 |
| Ghana | | 0.96 (0.84, 1.10) | 0.57 | 0.84 (0.72, 0.99) | 0.042 | 0.91 (0.71, 1.17) | 0.46 | 0.62 (0.52, 0.75) | <0.0001 | 0.75 (0.62, 0.91) | 0.0027 | 0.79 (0.64, 0.97) | 0.026 |
| Guinea-Bissau | Reference | 0.95 (0.79, 1.15) | 0.59 | 0.77 (0.59, 0.99) | 0.044 | 0.70 (0.54, 0.90) | 0.0064 | 0.63 (0.48, 0.83) | 0.0008 | 0.52 (0.35, 0.76) | 0.0008 | 0.37 (0.24, 0.58) | <0.0001 |
| Nigeria | | 0.78 (0.70, 0.86) | <0.0001 | 0.76 (0.68, 0.85) | <0.0001 | 0.67 (0.58, 0.76) | <0.0001 | 0.55 (0.48, 0.63) | <0.0001 | 0.64 (0.55, 0.74) | <0.0001 | 0.45 (0.38, 0.53) | <0.0001 |
| Sao Tome & Principe | | 0.83 (0.58, 1.19) | 0.32 | 0.42 (0.27, 0.65) | 0.0001 | 0.54 (0.36, 0.82) | 0.0038 | 0.35 (0.22, 0.57) | <0.0001 | 0.56 (0.37, 0.87) | 0.0096 | 0.38 (0.20, 0.71) | 0.0026 |
| Sierra Leone | | 0.87 (0.76, 1.00) | 0.044 | 0.82 (0.73, 0.93) | 0.0013 | 0.82 (0.70, 0.95) | 0.0078 | 0.87 (0.76, 0.99) | 0.036 | 0.71 (0.59, 0.85) | 0.0002 | 0.85 (0.71, 1.01) | 0.071 |
| Togo | | 0.87 (0.70, 1.09) | 0.22 | 0.69 (0.54, 0.89) | 0.0047 | 0.72 (0.54, 0.97) | 0.030 | 0.56 (0.41, 0.76) | 0.0002 | 0.50 (0.36, 0.70) | <0.0001 | 0.64 (0.38, 1.08) | 0.093 |
| **Pooled PR (95% CI)** | | **0.88 (0.83, 0.93)** | N/A | **0.74 (0.64, 0.85)** | N/A | **0.70 (0.60, 0.82)** | N/A | **0.64 (0.51, 0.79)** | N/A | **0.63 (0.51, 0.78)** | N/A | **0.61 (0.48, 0.79)** | N/A |
| **Eastern and Southern Africa** | | | | | | | | | | | | | |
| Lesotho | | 1.08 (0.84, 1.38) | 0.54 | 0.82 (0.61, 1.11) | 0.20 | 0.69 (0.50, 0.95) | 0.023 | 0.73 (0.56, 0.96) | 0.022 | 0.74 (0.52, 1.05) | 0.090 | 1.02 (0.73, 1.43) | 0.91 |
| Madagascar | | 0.88 (0.72, 1.08) | 0.23 | 0.74 (0.60, 0.91) | 0.0047 | 0.79 (0.61, 1.01) | 0.058 | 0.65 (0.52, 0.82) | 0.0003 | 0.72 (0.54, 0.96) | 0.027 | 0.66 (0.48, 0.90) | 0.0085 |
| Malawi | Reference | 0.63 (0.55, 0.72) | <0.0001 | 0.57 (0.49, 0.67) | <0.0001 | 0.51 (0.43, 0.60) | <0.0001 | 0.48 (0.40, 0.57) | <0.0001 | 0.58 (0.46, 0.73) | <0.0001 | 0.40 (0.32, 0.52) | <0.0001 |
| Zimbabwe | | 0.86 (0.74, 1.00) | 0.050 | 0.69 (0.59, 0.81) | <0.0001 | 0.63 (0.51, 0.78) | <0.0001 | 0.56 (0.46, 0.67) | <0.0001 | 0.62 (0.50, 0.76) | <0.0001 | 0.69 (0.55, 0.87) | 0.0016 |
| **Pooled PR (95% CI)** | | **0.84 (0.67, 1.04)** | N/A | **0.68 (0.59, 0.79)** | N/A | **0.63 (0.52, 0.76)** | N/A | **0.59 (0.49, 0.71)** | N/A | **0.64 (0.56, 0.74)** | N/A | **0.65 (0.45, 0.94)** | N/A |
| **Middle East and North Africa** | | | | | | | | | | | | | |
| Algeria | | 0.92 (0.86, 0.99) | 0.022 | 0.72 (0.66, 0.78) | <0.0001 | 0.63 (0.57, 0.69) | <0.0001 | 0.61 (0.56, 0.67) | <0.0001 | 0.59 (0.53, 0.65) | <0.0001 | 0.49 (0.43, 0.55) | <0.0001 |
| Iraq | | 1.23 (0.91, 1.66) | 0.18 | 1.18 (0.88, 1.57) | 0.27 | 1.57 (1.11, 2.22) | 0.010 | 1.32 (0.93, 1.85) | 0.12 | 1.69 (1.24, 2.29) | 0.0008 | 1.43 (1.06, 1.93) | 0.021 |
| State of Palestine | Reference | 0.94 (0.60, 1.46) | 0.77 | 0.88 (0.57, 1.36) | 0.57 | 0.94 (0.60, 1.46) | 0.78 | 0.86 (0.54, 1.35) | 0.51 | 1.00 (0.64, 1.55) | 0.99 | 1.05 (0.68, 1.64) | 0.82 |
| Tunisia | | 1.07 (0.17, 6.92) | 0.94 | 0.85 (0.14, 5.29) | 0.86 | 0.82 (0.13, 5.07) | 0.83 | 0.86 (0.14, 5.24) | 0.87 | 0.82 (0.13, 5.09) | 0.83 | 0.79 (0.13, 4.96) | 0.80 |
| **Pooled PR (95% CI)** | | **0.98 (0.84, 1.14)** | N/A | **0.87 (0.68, 1.11)** | N/A | **0.94 (0.62, 1.43)** | N/A | **0.85 (0.59, 1.22)** | N/A | **0.96 (0.59, 1.57)** | N/A | **0.87 (0.52, 1.45)** | N/A |
| **South Asia** | | | | | | | | | | | | | |
| Afghanistan | | 0.94 (0.88, 1.00) | 0.042 | 0.90 (0.84, 0.96) | 0.0020 | 0.85 (0.79, 0.91) | <0.0001 | 0.93 (0.87, 1.00) | 0.054 | 0.89 (0.81, 0.98) | 0.022 | 0.88 (0.79, 0.97) | 0.013 |
| Bangladesh | Reference | 0.87 (0.79, 0.96) | 0.0064 | 0.83 (0.75, 0.93) | 0.0006 | 0.82 (0.74, 0.90) | <0.0001 | 0.78 (0.71, 0.87) | <0.0001 | 0.83 (0.74, 0.93) | 0.0012 | 0.71 (0.62, 0.82) | <0.0001 |
| Nepal | | 1.10 | 0.29 | 1.02 | 0.86 | 1.17 | 0.14 | 1.17 | 0.11 | 1.35 | 0.027 | 1.23 | 0.082 |

| | | | | | | | | | | | | | |
|---|---|---|---|---|---|---|---|---|---|---|---|---|---|
| | | (0.92, 1.32) | | (0.84, 1.23) | | (0.95, 1.44) | | (0.97, 1.43) | | (1.04, 1.75) | | (0.97, 1.57) | |
| Pakistan (Balochistan) | | 1.17 (1.06, 1.28) | 0.0014 | 1.12 (1.01, 1.23) | 0.028 | 0.88 (0.78, 0.99) | 0.030 | 0.88 (0.78, 1.00) | 0.044 | 0.85 (0.75, 0.97) | 0.014 | 0.99 (0.86, 1.13) | 0.83 |
| Pakistan (Khyber Pakhtunkhwa) | | 0.80 (0.74, 0.87) | <0.0001 | 0.80 (0.73, 0.87) | <0.0001 | 0.68 (0.62, 0.75) | <0.0001 | 0.66 (0.59, 0.72) | <0.0001 | 0.71 (0.64, 0.80) | <0.0001 | 0.56 (0.48, 0.65) | <0.0001 |
| Pakistan (Sindh) | | 1.03 (0.98, 1.08) | 0.31 | 0.98 (0.93, 1.03) | 0.46 | 0.98 (0.93, 1.04) | 0.57 | 0.91 (0.86, 0.96) | 0.0010 | 0.87 (0.82, 0.93) | <0.0001 | 0.90 (0.83, 0.97) | 0.0088 |
| Pakistan (Punjab) | | 0.92 (0.87, 0.97) | 0.0027 | 0.83 (0.78, 0.88) | <0.0001 | 0.85 (0.80, 0.90) | <0.0001 | 0.85 (0.80, 0.90) | <0.0001 | 0.82 (0.77, 0.88) | <0.0001 | 0.83 (0.77, 0.90) | <0.0001 |
| **Pooled PR (95% CI)** | | **0.96 (0.87, 1.06)** | N/A | **0.91 (0.83, 1.00)** | N/A | **0.87 (0.77, 0.98)** | N/A | **0.86 (0.76, 0.98)** | N/A | **0.87 (0.76, 0.99)** | N/A | **0.84 (0.71, 1.01)** | N/A |
| **East Asia and the Pacific** | | | | | | | | | | | | | |
| Fiji | | 0.99 (0.80, 1.22) | 0.91 | 1.02 (0.85, 1.24) | 0.80 | 0.95 (0.78, 1.16) | 0.62 | 1.06 (0.88, 1.26) | 0.55 | 1.01 (0.83, 1.23) | 0.91 | 1.15 (0.95, 1.38) | 0.16 |
| Kiribati | | 1.07 (0.82, 1.39) | 0.63 | 0.96 (0.71, 1.29) | 0.77 | 1.01 (0.75, 1.35) | 0.97 | 1.15 (0.85, 1.55) | 0.36 | 0.97 (0.68, 1.38) | 0.85 | 1.20 (0.88, 1.63) | 0.25 |
| Lao PDR | | 0.95 (0.84, 1.08) | 0.44 | 0.85 (0.74, 0.97) | 0.017 | 0.90 (0.79, 1.04) | 0.15 | 0.67 (0.58, 0.77) | <0.0001 | 0.72 (0.62, 0.84) | <0.0001 | 0.77 (0.64, 0.93) | 0.0075 |
| Mongolia | | 0.71 (0.38, 1.32) | 0.28 | 0.54 (0.31, 0.95) | 0.032 | 0.56 (0.35, 0.92) | 0.021 | 0.53 (0.31, 0.90) | 0.018 | 0.62 (0.36, 1.07) | 0.085 | 0.55 (0.33, 0.94) | 0.030 |
| Samoa | Reference | 1.15 (0.79, 1.69) | 0.47 | 0.90 (0.64, 1.28) | 0.57 | 0.73 (0.48, 1.12) | 0.15 | 1.15 (0.78, 1.70) | 0.48 | 0.90 (0.60, 1.33) | 0.59 | 1.07 (0.71, 1.63) | 0.74 |
| Tonga | | 1.05 (0.81, 1.36) | 0.72 | 1.05 (0.79, 1.42) | 0.72 | 0.63 (0.40, 0.99) | 0.046 | 0.76 (0.55, 1.03) | 0.076 | 1.22 (0.86, 1.74) | 0.27 | 0.90 (0.58, 1.40) | 0.65 |
| Tuvalu | | 1.42 (0.81, 2.49) | 0.22 | 0.78 (0.40, 1.53) | 0.47 | 0.94 (0.45, 1.95) | 0.86 | 1.19 (0.61, 2.34) | 0.61 | 0.70 (0.30, 1.63) | 0.40 | 1.72 (0.88, 3.35) | 0.11 |
| Vietnam | | 1.03 (0.71, 1.50) | 0.86 | 0.88 (0.60, 1.28) | 0.50 | 0.60 (0.40, 0.91) | 0.016 | 0.33 (0.21, 0.53) | <0.0001 | 0.53 (0.35, 0.79) | 0.0018 | 0.60 (0.38, 0.95) | 0.029 |
| **Pooled PR (95% CI)** | | **1.02 (0.90, 1.15)** | N/A | **0.90 (0.78, 1.04)** | N/A | **0.81 (0.68, 0.96)** | N/A | **0.79 (0.58, 1.08)** | N/A | **0.83 (0.68, 1.02)** | N/A | **0.93 (0.72, 1.20)** | N/A |
| **Europe and Central Asia** | | | | | | | | | | | | | |
| Kosovo | | 0.87 (0.68, 1.12) | 0.29 | 0.63 (0.47, 0.86) | 0.0031 | 0.71 (0.52, 0.96) | 0.027 | 0.83 (0.63, 1.11) | 0.20 | 0.73 (0.55, 0.97) | 0.031 | 0.85 (0.65, 1.12) | 0.24 |
| Kyrgyzstan | | 0.84 (0.57, 1.22) | 0.36 | 0.43 (0.28, 0.66) | 0.0001 | 0.38 (0.23, 0.60) | <0.0001 | 0.43 (0.28, 0.67) | 0.0002 | 0.43 (0.26, 0.70) | 0.0007 | 0.57 (0.35, 0.92) | 0.023 |
| Montenegro | | 0.40 (0.17, 0.98) | 0.046 | 0.91 (0.38, 2.16) | 0.82 | 0.37 (0.16, 0.90) | 0.027 | 0.48 (0.20, 1.13) | 0.093 | 0.44 (0.18, 1.06) | 0.067 | 0.55 (0.20, 1.48) | 0.23 |
| Republic of North Macedonia | Reference | 0.84 (0.39, 1.81) | 0.66 | 0.95 (0.44, 2.03) | 0.89 | 0.71 (0.37, 1.36) | 0.30 | 0.46 (0.25, 0.86) | 0.015 | 0.78 (0.45, 1.36) | 0.39 | 0.79 (0.40, 1.57) | 0.50 |
| Serbia | | 0.82 (0.53, 1.27) | 0.38 | 0.71 (0.45, 1.13) | 0.15 | 0.51 (0.32, 0.80) | 0.0038 | 0.51 (0.31, 0.84) | 0.0084 | 0.54 (0.35, 0.83) | 0.0054 | 0.67 (0.42, 1.06) | 0.086 |
| Turkmenistan | | 2.14 (0.23, 20.30) | 0.50 | 1.37 (0.16, 11.52) | 0.77 | 1.04 (0.12, 8.90) | 0.97 | 0.91 (0.10, 8.48) | 0.93 | 1.17 (0.13, 10.61) | 0.89 | 2.98 (0.37, 24.23) | 0.31 |
| Uzbekistan | | 0.62 (0.39, 0.97) | 0.038 | 0.74 (0.47, 1.17) | 0.20 | 0.81 (0.51, 1.29) | 0.37 | 1.03 (0.67, 1.58) | 0.90 | 0.88 (0.56, 1.39) | 0.58 | 0.96 (0.58, 1.58) | 0.87 |
| **Pooled PR (95% CI)** | | **0.77 (0.57, 1.02)** | N/A | **0.67 (0.51, 0.87)** | N/A | **0.59 (0.44, 0.78)** | N/A | **0.63 (0.47, 0.84)** | N/A | **0.64 (0.49, 0.84)** | N/A | **0.76 (0.55, 1.05)** | N/A |
| **Latin America and Caribbean** | | | | | | | | | | | | | |
| Costa Rica | | 0.95 (0.59, 1.56) | 0.85 | 0.80 (0.52, 1.24) | 0.32 | 0.85 (0.55, 1.33) | 0.49 | 0.99 (0.66, 1.50) | 0.98 | 0.97 (0.62, 1.53) | 0.90 | 1.10 (0.72, 1.68) | 0.66 |
| Cuba | | 0.87 (0.69, 1.10) | 0.26 | 0.98 (0.77, 1.24) | 0.84 | 0.87 (0.68, 1.12) | 0.28 | 0.98 (0.75, 1.27) | 0.86 | 0.81 (0.64, 1.03) | 0.087 | 1.03 (0.83, 1.28) | 0.77 |
| Dominican Republic | Reference | 0.90 (0.79, 1.01) | 0.079 | 0.88 (0.78, 1.00) | 0.047 | 0.80 (0.71, 0.91) | 0.0008 | 0.86 (0.76, 0.97) | 0.018 | 0.90 (0.80, 1.03) | 0.13 | 0.83 (0.72, 0.96) | 0.013 |
| Guyana | | 0.95 (0.77, 1.17) | 0.64 | 0.84 (0.66, 1.08) | 0.18 | 0.89 (0.68, 1.17) | 0.40 | 1.03 (0.81, 1.31) | 0.83 | 0.99 (0.79, 1.25) | 0.93 | 0.98 (0.77, 1.24) | 0.85 |
| Honduras | | 0.94 | 0.30 | 0.89 | 0.047 | 0.98 | 0.72 | 0.88 | 0.043 | 0.96 | 0.51 | 0.88 | 0.085 |

| | | | | | | | | | | | | | |
|---|---|---|---|---|---|---|---|---|---|---|---|---|---|
| | | (0.84, 1.05) | | (0.79, 1.00) | | (0.86, 1.11) | | (0.77, 1.00) | | (0.85, 1.09) | | (0.75, 1.02) | |
| Suriname | | 1.01 (0.78, 1.29) | 0.96 | 0.89 (0.66, 1.21) | 0.47 | 0.93 (0.69, 1.24) | 0.61 | 0.99 (0.76, 1.28) | 0.92 | 0.77 (0.57, 1.05) | 0.094 | 0.92 (0.68, 1.23) | 0.56 |
| Turks & Caicos Islands | | 0.69 (0.16, 2.93) | 0.60 | 0.57 (0.16, 2.03) | 0.37 | 0.50 (0.14, 1.81) | 0.28 | 0.42 (0.19, 0.95) | 0.039 | 0.21 (0.09, 0.50) | 0.0011 | 0.30 (0.11, 0.87) | 0.028 |
| **Pooled PR (95% CI)** | | **0.93 (0.86, 1.00)** | N/A | **0.89 (0.81, 0.97)** | N/A | **0.88 (0.79, 0.99)** | N/A | **0.91 (0.76, 1.09)** | N/A | **0.80 (0.56, 1.13)** | N/A | **0.90 (0.70, 1.15)** | N/A |
| **All regions** | | | | | | | | | | | | | |
| **Pooled PR (95% CI)** | Reference | **0.92 (0.87, 0.97)** | N/A | **0.81 (0.76, 0.87)** | N/A | **0.77 (0.71, 0.83)** | N/A | **0.74 (0.67, 0.82)** | N/A | **0.75 (0.68, 0.83)** | N/A | **0.78 (0.70, 0.87)** | N/A |

All values are weighted to reflect the survey sampling design. Women who answered "Don't know/no such activity" or had missing data/no response for menstrual-related absenteeism were excluded from the analysis. P values not adjusted for multiple testing.
PR = Prevalence Ratio, CI = Confidence Interval.

**Table S20: Multivariable analysis between age group and menstrual-related absenteeism.**

| Country | Age group (years) | | | | | | | | | | | | |
|---|---|---|---|---|---|---|---|---|---|---|---|---|---|
| | 15-19 years | 20-24 | | 25-29 | | 30-34 | | 35-39 | | 40-44 | | 45-49 | |
| | | PR (95% CI) | P value | PR (95% CI) | P value | PR (95% CI) | P value | PR (95% CI) | P value | PR (95% CI) | P value | PR (95% CI) | P value |
| **West and Central Africa** | | | | | | | | | | | | | |
| Central African Republic | | 0.90 (0.80, 1.01) | 0.068 | 0.80 (0.70, 0.91) | 0.0008 | 0.80 (0.70, 0.92) | 0.0013 | 0.90 (0.79, 1.02) | 0.11 | 0.88 (0.76, 1.03) | 0.12 | 0.88 (0.72, 1.08) | 0.23 |
| Chad | | 0.94 (0.87, 1.01) | 0.10 | 0.89 (0.81, 0.97) | 0.0065 | 0.89 (0.81, 0.97) | 0.0090 | 0.92 (0.83, 1.01) | 0.070 | 0.88 (0.78, 1.00) | 0.042 | 0.83 (0.73, 0.95) | 0.0075 |
| Democratic Republic of Congo | | 0.88 (0.73, 1.05) | 0.16 | 0.85 (0.71, 1.01) | 0.068 | 0.67 (0.54, 0.84) | 0.0004 | 0.81 (0.64, 1.03) | 0.081 | 0.72 (0.56, 0.93) | 0.012 | 0.84 (0.64, 1.11) | 0.22 |
| Gambia | | 0.82 (0.73, 0.92) | 0.0010 | 0.50 (0.43, 0.58) | <0.0001 | 0.39 (0.33, 0.46) | <0.0001 | 0.34 (0.29, 0.41) | <0.0001 | 0.29 (0.23, 0.37) | <0.0001 | 0.32 (0.23, 0.43) | <0.0001 |
| Ghana | | 0.96 (0.84, 1.11) | 0.60 | 0.85 (0.72, 1.00) | 0.048 | 0.92 (0.72, 1.17) | 0.48 | 0.62 (0.52, 0.75) | <0.0001 | 0.75 (0.62, 0.91) | 0.0035 | 0.79 (0.64, 0.98) | 0.028 |
| Guinea-Bissau | Reference | 0.96 (0.79, 1.15) | 0.65 | 0.77 (0.60, 1.00) | 0.047 | 0.70 (0.54, 0.91) | 0.0081 | 0.65 (0.50, 0.84) | 0.0013 | 0.53 (0.36, 0.77) | 0.0009 | 0.38 (0.24, 0.60) | <0.0001 |
| Nigeria | | 0.78 (0.70, 0.86) | <0.0001 | 0.76 (0.68, 0.85) | <0.0001 | 0.67 (0.59, 0.76) | <0.0001 | 0.55 (0.48, 0.63) | <0.0001 | 0.64 (0.55, 0.74) | <0.0001 | 0.45 (0.38, 0.53) | <0.0001 |
| Sao Tome & Principe | | 0.83 (0.58, 1.18) | 0.30 | 0.42 (0.27, 0.64) | 0.0001 | 0.55 (0.36, 0.83) | 0.0044 | 0.36 (0.22, 0.57) | <0.0001 | 0.56 (0.37, 0.87) | 0.0096 | 0.37 (0.20, 0.69) | 0.0020 |
| Sierra Leone | | 0.87 (0.76, 1.00) | 0.043 | 0.82 (0.73, 0.92) | 0.0012 | 0.81 (0.70, 0.94) | 0.0067 | 0.86 (0.76, 0.98) | 0.029 | 0.70 (0.59, 0.84) | 0.0001 | 0.84 (0.71, 1.00) | 0.053 |
| Togo | | 0.87 (0.70, 1.08) | 0.22 | 0.69 (0.54, 0.90) | 0.0050 | 0.72 (0.54, 0.97) | 0.029 | 0.56 (0.41, 0.76) | 0.0002 | 0.50 (0.36, 0.70) | <0.0001 | 0.64 (0.38, 1.07) | 0.086 |
| **Pooled PR (95% CI)** | | **0.88 (0.83, 0.93)** | N/A | **0.74 (0.64, 0.85)** | N/A | **0.70 (0.60, 0.82)** | N/A | **0.64 (0.51, 0.79)** | N/A | **0.63 (0.51, 0.77)** | N/A | **0.61 (0.48, 0.79)** | N/A |
| **Eastern and Southern Africa** | | | | | | | | | | | | | |
| Lesotho | | 1.09 (0.85, 1.40) | 0.52 | 0.83 (0.62, 1.12) | 0.22 | 0.69 (0.50, 0.97) | 0.031 | 0.74 (0.57, 0.96) | 0.026 | 0.74 (0.52, 1.05) | 0.096 | 1.02 (0.73, 1.44) | 0.89 |
| Madagascar | | 0.89 (0.73, 1.08) | 0.24 | 0.73 (0.59, 0.90) | 0.0034 | 0.78 (0.61, 1.00) | 0.051 | 0.65 (0.52, 0.82) | 0.0003 | 0.72 (0.55, 0.96) | 0.026 | 0.66 (0.48, 0.90) | 0.0099 |
| Malawi | Reference | 0.63 (0.56, 0.72) | <0.0001 | 0.58 (0.49, 0.67) | <0.0001 | 0.51 (0.43, 0.61) | <0.0001 | 0.48 (0.40, 0.57) | <0.0001 | 0.58 (0.46, 0.72) | <0.0001 | 0.40 (0.32, 0.51) | <0.0001 |
| Zimbabwe | | 0.87 (0.75, 1.01) | 0.066 | 0.70 (0.59, 0.82) | <0.0001 | 0.64 (0.52, 0.79) | <0.0001 | 0.56 (0.46, 0.67) | <0.0001 | 0.62 (0.50, 0.76) | <0.0001 | 0.69 (0.55, 0.87) | 0.0019 |
| **Pooled PR (95% CI)** | | **0.84 (0.68, 1.04)** | N/A | **0.68 (0.59, 0.80)** | N/A | **0.63 (0.53, 0.76)** | N/A | **0.59 (0.49, 0.71)** | N/A | **0.64 (0.56, 0.75)** | N/A | **0.65 (0.45, 0.94)** | N/A |
| **Middle East and North Africa** | | | | | | | | | | | | | |
| Algeria | | 0.92 (0.86, 0.99) | 0.022 | 0.72 (0.67, 0.78) | <0.0001 | 0.63 (0.58, 0.70) | <0.0001 | 0.61 (0.56, 0.67) | <0.0001 | 0.59 (0.53, 0.65) | <0.0001 | 0.48 (0.43, 0.54) | <0.0001 |
| Iraq | | 1.24 (0.92, 1.67) | 0.16 | 1.19 (0.89, 1.59) | 0.23 | 1.60 (1.13, 2.26) | 0.0074 | 1.33 (0.95, 1.87) | 0.094 | 1.70 (1.25, 2.30) | 0.0006 | 1.46 (1.08, 1.97) | 0.014 |
| State of Palestine | Reference | 0.93 (0.60, 1.45) | 0.76 | 0.88 (0.57, 1.36) | 0.56 | 0.93 (0.60, 1.45) | 0.76 | 0.85 (0.54, 1.34) | 0.48 | 0.99 (0.64, 1.54) | 0.97 | 1.05 (0.68, 1.63) | 0.83 |
| Tunisia | | 1.11 (0.17, 7.08) | 0.92 | 0.87 (0.14, 5.41) | 0.88 | 0.85 (0.14, 5.24) | 0.86 | 0.90 (0.15, 5.41) | 0.90 | 0.85 (0.14, 5.21) | 0.86 | 0.83 (0.13, 5.13) | 0.84 |
| **Pooled PR (95% CI)** | | **0.98 (0.84, 1.15)** | N/A | **0.88 (0.68, 1.12)** | N/A | **0.95 (0.62, 1.45)** | N/A | **0.86 (0.59, 1.23)** | N/A | **0.97 (0.59, 1.57)** | N/A | **0.88 (0.52, 1.47)** | N/A |
| **South Asia** | | | | | | | | | | | | | |
| Afghanistan | | 0.94 (0.88, 1.00) | 0.041 | 0.90 (0.84, 0.96) | 0.0017 | 0.85 (0.79, 0.91) | <0.0001 | 0.93 (0.87, 1.00) | 0.051 | 0.89 (0.81, 0.98) | 0.021 | 0.88 (0.79, 0.97) | 0.013 |
| Bangladesh | Reference | 0.88 (0.80, 0.97) | 0.0098 | 0.84 (0.76, 0.93) | 0.0011 | 0.82 (0.74, 0.90) | 0.0001 | 0.79 (0.71, 0.87) | <0.0001 | 0.83 (0.74, 0.93) | 0.0016 | 0.71 (0.62, 0.81) | <0.0001 |
| Nepal | | 1.11 | 0.26 | 1.04 | 0.70 | 1.19 | 0.10 | 1.19 | 0.081 | 1.35 | 0.024 | 1.25 | 0.066 |

| | | | | | | | | | | | | | |
|---|---|---|---|---|---|---|---|---|---|---|---|---|---|
| | | (0.93, 1.33) | | (0.86, 1.25) | | (0.97, 1.46) | | (0.98, 1.44) | | (1.04, 1.76) | | (0.99, 1.58) | |
| Pakistan (Balochistan) | | 1.16 (1.06, 1.28) | 0.0016 | 1.11 (1.01, 1.22) | 0.039 | 0.88 (0.78, 0.99) | 0.032 | 0.88 (0.78, 1.00) | 0.045 | 0.85 (0.75, 0.96) | 0.012 | 0.98 (0.86, 1.13) | 0.82 |
| Pakistan (Khyber Pakhtunkhwa) | | 0.80 (0.74, 0.87) | <0.0001 | 0.80 (0.73, 0.87) | <0.0001 | 0.68 (0.62, 0.75) | <0.0001 | 0.66 (0.59, 0.72) | <0.0001 | 0.71 (0.63, 0.80) | <0.0001 | 0.56 (0.48, 0.65) | <0.0001 |
| Pakistan (Sindh) | | 1.04 (0.99, 1.10) | 0.089 | 1.00 (0.95, 1.05) | 0.94 | 1.00 (0.95, 1.05) | 0.89 | 0.93 (0.88, 0.98) | 0.0097 | 0.89 (0.83, 0.95) | 0.0003 | 0.92 (0.85, 0.99) | 0.024 |
| Pakistan (Punjab) | | 0.92 (0.87, 0.97) | 0.0025 | 0.83 (0.78, 0.88) | <0.0001 | 0.85 (0.80, 0.90) | <0.0001 | 0.85 (0.80, 0.90) | <0.0001 | 0.82 (0.77, 0.88) | <0.0001 | 0.83 (0.77, 0.90) | <0.0001 |
| **Pooled PR (95% CI)** | | **0.97 (0.88, 1.06)** | N/A | **0.92 (0.84, 1.01)** | N/A | **0.87 (0.77, 0.99)** | N/A | **0.87 (0.77, 0.99)** | N/A | **0.87 (0.76, 1.00)** | N/A | **0.85 (0.70, 1.01)** | N/A |
| **East Asia and the Pacific** | | | | | | | | | | | | | |
| Fiji | | 0.99 (0.80, 1.22) | 0.91 | 1.01 (0.84, 1.22) | 0.90 | 0.94 (0.77, 1.15) | 0.54 | 1.04 (0.87, 1.24) | 0.65 | 1.00 (0.83, 1.22) | 0.97 | 1.13 (0.94, 1.36) | 0.21 |
| Kiribati | | 1.05 (0.81, 1.37) | 0.69 | 0.95 (0.71, 1.29) | 0.75 | 1.00 (0.75, 1.35) | 0.98 | 1.15 (0.86, 1.55) | 0.35 | 0.97 (0.68, 1.38) | 0.86 | 1.20 (0.88, 1.64) | 0.25 |
| Lao PDR | | 0.95 (0.84, 1.08) | 0.42 | 0.84 (0.74, 0.97) | 0.014 | 0.90 (0.78, 1.04) | 0.15 | 0.67 (0.58, 0.77) | <0.0001 | 0.72 (0.62, 0.83) | <0.0001 | 0.77 (0.64, 0.93) | 0.0065 |
| Mongolia | | 0.71 (0.39, 1.32) | 0.28 | 0.54 (0.31, 0.95) | 0.032 | 0.56 (0.34, 0.91) | 0.020 | 0.53 (0.31, 0.89) | 0.017 | 0.61 (0.36, 1.05) | 0.073 | 0.55 (0.32, 0.93) | 0.027 |
| Samoa | Reference | 1.15 (0.79, 1.69) | 0.47 | 0.90 (0.64, 1.28) | 0.57 | 0.73 (0.48, 1.12) | 0.15 | 1.15 (0.78, 1.70) | 0.48 | 0.89 (0.60, 1.33) | 0.58 | 1.07 (0.71, 1.63) | 0.74 |
| Tonga | | 1.06 (0.81, 1.38) | 0.68 | 1.07 (0.79, 1.43) | 0.67 | 0.63 (0.40, 0.99) | 0.047 | 0.76 (0.56, 1.04) | 0.085 | 1.22 (0.85, 1.74) | 0.27 | 0.91 (0.58, 1.42) | 0.67 |
| Tuvalu | | 1.47 (0.85, 2.52) | 0.16 | 0.79 (0.41, 1.54) | 0.49 | 0.98 (0.47, 2.02) | 0.95 | 1.20 (0.62, 2.32) | 0.59 | 0.68 (0.29, 1.58) | 0.36 | 1.64 (0.84, 3.23) | 0.15 |
| Vietnam | | 1.02 (0.71, 1.48) | 0.91 | 0.88 (0.60, 1.29) | 0.51 | 0.60 (0.40, 0.91) | 0.016 | 0.33 (0.21, 0.53) | <0.0001 | 0.52 (0.35, 0.78) | 0.0016 | 0.60 (0.38, 0.95) | 0.030 |
| **Pooled PR (95% CI)** | | **1.02 (0.90, 1.16)** | N/A | **0.90 (0.78, 1.04)** | N/A | **0.81 (0.68, 0.96)** | N/A | **0.79 (0.58, 1.08)** | N/A | **0.83 (0.67, 1.01)** | N/A | **0.93 (0.72, 1.18)** | N/A |
| **Europe and Central Asia** | | | | | | | | | | | | | |
| Kosovo | | 0.87 (0.68, 1.12) | 0.28 | 0.63 (0.47, 0.86) | 0.0031 | 0.71 (0.52, 0.96) | 0.026 | 0.83 (0.63, 1.10) | 0.20 | 0.73 (0.54, 0.97) | 0.030 | 0.85 (0.65, 1.11) | 0.24 |
| Kyrgyzstan | | 0.83 (0.57, 1.21) | 0.34 | 0.43 (0.28, 0.65) | 0.0001 | 0.37 (0.23, 0.59) | <0.0001 | 0.43 (0.28, 0.66) | 0.0002 | 0.43 (0.26, 0.69) | 0.0006 | 0.56 (0.34, 0.91) | 0.021 |
| Montenegro | | 0.40 (0.16, 0.99) | 0.047 | 0.90 (0.37, 2.17) | 0.81 | 0.37 (0.15, 0.89) | 0.027 | 0.47 (0.20, 1.13) | 0.090 | 0.43 (0.18, 1.06) | 0.067 | 0.54 (0.20, 1.47) | 0.23 |
| Republic of North Macedonia | Reference | 0.85 (0.40, 1.81) | 0.67 | 0.96 (0.45, 2.03) | 0.91 | 0.72 (0.38, 1.38) | 0.32 | 0.48 (0.25, 0.89) | 0.021 | 0.79 (0.46, 1.37) | 0.41 | 0.80 (0.41, 1.60) | 0.53 |
| Serbia | | 0.84 (0.54, 1.30) | 0.43 | 0.73 (0.46, 1.15) | 0.17 | 0.53 (0.33, 0.83) | 0.0057 | 0.52 (0.32, 0.86) | 0.011 | 0.55 (0.35, 0.84) | 0.0063 | 0.68 (0.43, 1.08) | 0.11 |
| Turkmenistan | | 2.15 (0.23, 20.41) | 0.51 | 1.37 (0.16, 11.61) | 0.77 | 1.04 (0.12, 8.95) | 0.97 | 0.91 (0.10, 8.54) | 0.94 | 1.18 (0.13, 10.80) | 0.89 | 3.00 (0.37, 24.68) | 0.30 |
| Uzbekistan | | 0.62 (0.39, 0.97) | 0.037 | 0.74 (0.47, 1.17) | 0.20 | 0.81 (0.51, 1.29) | 0.38 | 1.03 (0.67, 1.57) | 0.90 | 0.88 (0.56, 1.38) | 0.57 | 0.96 (0.58, 1.58) | 0.87 |
| **Pooled PR (95% CI)** | | **0.77 (0.58, 1.03)** | N/A | **0.67 (0.51, 0.87)** | N/A | **0.59 (0.44, 0.78)** | N/A | **0.63 (0.47, 0.85)** | N/A | **0.64 (0.49, 0.84)** | N/A | **0.76 (0.55, 1.06)** | N/A |
| **Latin America and Caribbean** | | | | | | | | | | | | | |
| Costa Rica | | 0.96 (0.59, 1.57) | 0.88 | 0.81 (0.53, 1.25) | 0.34 | 0.86 (0.55, 1.34) | 0.50 | 0.99 (0.66, 1.50) | 0.98 | 0.98 (0.62, 1.53) | 0.92 | 1.11 (0.73, 1.70) | 0.62 |
| Cuba | | 0.89 (0.70, 1.12) | 0.32 | 0.97 (0.76, 1.24) | 0.81 | 0.88 (0.68, 1.13) | 0.31 | 0.98 (0.75, 1.27) | 0.86 | 0.80 (0.63, 1.01) | 0.063 | 1.03 (0.83, 1.28) | 0.78 |
| Dominican Republic | Reference | 0.90 (0.79, 1.01) | 0.083 | 0.89 (0.79, 1.00) | 0.050 | 0.80 (0.71, 0.91) | 0.0009 | 0.86 (0.76, 0.98) | 0.020 | 0.91 (0.80, 1.03) | 0.13 | 0.83 (0.72, 0.96) | 0.014 |
| Guyana | | 0.95 (0.77, 1.17) | 0.63 | 0.84 (0.65, 1.08) | 0.17 | 0.89 (0.68, 1.16) | 0.39 | 1.01 (0.80, 1.29) | 0.91 | 0.99 (0.79, 1.25) | 0.95 | 0.98 (0.78, 1.25) | 0.89 |
| Honduras | | 0.95 | 0.33 | 0.89 | 0.053 | 0.98 | 0.78 | 0.88 | 0.048 | 0.96 | 0.55 | 0.88 | 0.10 |

| | | | | | | | | | | | | | |
|---|---|---|---|---|---|---|---|---|---|---|---|---|---|
| | | (0.85, 1.06) | | (0.79, 1.00) | | (0.86, 1.12) | | (0.77, 1.00) | | (0.85, 1.09) | | (0.76, 1.02) | |
| Suriname | | 1.00<br>(0.78, 1.29) | 0.99 | 0.89<br>(0.66, 1.21) | 0.46 | 0.93<br>(0.69, 1.24) | 0.60 | 0.98<br>(0.76, 1.28) | 0.90 | 0.77<br>(0.57, 1.04) | 0.092 | 0.91<br>(0.68, 1.22) | 0.54 |
| Turks & Caicos Islands | | 0.67<br>(0.16, 2.85) | 0.57 | 0.56<br>(0.16, 1.98) | 0.35 | 0.50<br>(0.14, 1.79) | 0.27 | 0.42<br>(0.19, 0.97) | 0.042 | 0.22<br>(0.09, 0.50) | 0.0012 | 0.31<br>(0.11, 0.87) | 0.029 |
| **Pooled PR (95% CI)** | | **0.93 (0.86, 1.01)** | N/A | **0.89 (0.81, 0.98)** | N/A | **0.89 (0.79, 0.99)** | N/A | **0.91 (0.76, 1.09)** | N/A | **0.80 (0.56, 1.13)** | N/A | **0.90 (0.71, 1.15)** | N/A |
| **All regions** | | | | | | | | | | | | | |
| **Pooled PR (95% CI)** | Reference | **0.92 (0.87, 0.97)** | N/A | **0.81 (0.76, 0.87)** | N/A | **0.77 (0.71, 0.83)** | N/A | **0.75 (0.68, 0.82)** | N/A | **0.75 (0.68, 0.83)** | N/A | **0.78 (0.70, 0.87)** | N/A |

All values are weighted to reflect the survey sampling design. Women who answered “Don’t know/no such activity” or had missing data/no response for menstrual-related absenteeism were excluded from the analysis. P values not adjusted for multiple testing. Results are adjusted for area type (urban/rural).
PR = Prevalence Ratio, CI = Confidence Interval.

**Table S21: Univariable analysis between wealth quintile and menstrual-related absenteeism.**

| Country | Wealth quintile | | | | | | | | |
|---|---|---|---|---|---|---|---|---|---|
| | Quintile 1 (Poorest) | | Quintile 2 | | Quintile 3 | Quintile 4 | | Quintile 5 (Richest) | |
| | PR (95% CI) | P value | PR (95% CI) | P value | | PR (95% CI) | P value | PR (95% CI) | P value |
| **West and Central Africa** | | | | | | | | | |
| Central African Republic | 1.09 (0.92, 1.30) | 0.31 | 1.14 (0.97, 1.33) | 0.12 | Reference | 0.92 (0.78, 1.08) | 0.29 | 0.81 (0.68, 0.97) | 0.022 |
| Chad | 1.02 (0.91, 1.15) | 0.70 | 0.99 (0.90, 1.10) | 0.90 | | 1.09 (0.99, 1.20) | 0.091 | 0.91 (0.80, 1.03) | 0.13 |
| Democratic Republic of Congo | 1.02 (0.81, 1.28) | 0.89 | 0.99 (0.83, 1.17) | 0.87 | | 0.91 (0.72, 1.15) | 0.44 | 0.82 (0.66, 1.02) | 0.081 |
| Gambia | 0.86 (0.72, 1.04) | 0.12 | 0.90 (0.76, 1.07) | 0.24 | | 1.14 (0.98, 1.34) | 0.090 | 1.24 (1.03, 1.48) | 0.021 |
| Ghana | 0.95 (0.78, 1.15) | 0.58 | 1.22 (0.97, 1.54) | 0.093 | | 0.92 (0.77, 1.12) | 0.41 | 0.87 (0.71, 1.05) | 0.15 |
| Guinea-Bissau | 1.21 (0.87, 1.67) | 0.25 | 1.20 (0.90, 1.59) | 0.21 | | 1.31 (0.98, 1.75) | 0.065 | 1.47 (1.02, 2.10) | 0.038 |
| Nigeria | 0.90 (0.79, 1.03) | 0.13 | 0.90 (0.80, 1.00) | 0.056 | | 0.94 (0.82, 1.06) | 0.31 | 0.87 (0.75, 1.01) | 0.073 |
| Sao Tome & Principe | 0.99 (0.67, 1.46) | 0.95 | 0.86 (0.57, 1.28) | 0.45 | | 1.18 (0.78, 1.78) | 0.44 | 1.17 (0.86, 1.60) | 0.32 |
| Sierra Leone | 1.26 (1.05, 1.50) | 0.011 | 1.00 (0.86, 1.16) | 0.95 | | 1.09 (0.92, 1.30) | 0.32 | 0.97 (0.79, 1.20) | 0.80 |
| Togo | 1.01 (0.77, 1.34) | 0.92 | 1.01 (0.77, 1.31) | 0.97 | | 0.90 (0.69, 1.19) | 0.47 | 0.85 (0.66, 1.10) | 0.21 |
| **Pooled PR (95% CI)** | **1.02 (0.93, 1.10)** | N/A | **1.00 (0.93, 1.09)** | N/A | | **1.02 (0.94, 1.11)** | N/A | **0.96 (0.85, 1.08)** | N/A |
| **Eastern and Southern Africa** | | | | | | | | | |
| Lesotho | 1.03 (0.80, 1.33) | 0.80 | 0.97 (0.75, 1.27) | 0.85 | Reference | 1.03 (0.74, 1.43) | 0.85 | 1.01 (0.76, 1.34) | 0.94 |
| Madagascar | 0.99 (0.80, 1.22) | 0.91 | 1.02 (0.80, 1.28) | 0.90 | | 1.05 (0.83, 1.33) | 0.69 | 1.60 (1.28, 2.01) | <0.0001 |
| Malawi | 0.95 (0.81, 1.12) | 0.55 | 1.00 (0.86, 1.17) | 0.96 | | 1.02 (0.87, 1.19) | 0.80 | 0.95 (0.82, 1.11) | 0.53 |
| Zimbabwe | 1.11 (0.91, 1.35) | 0.30 | 1.09 (0.91, 1.30) | 0.34 | | 0.92 (0.77, 1.10) | 0.35 | 0.94 (0.79, 1.13) | 0.53 |
| **Pooled PR (95% CI)** | **1.01 (0.91, 1.12)** | N/A | **1.03 (0.93, 1.13)** | N/A | | **0.99 (0.89, 1.10)** | N/A | **1.09 (0.85, 1.40)** | N/A |
| **Middle East and North Africa** | | | | | | | | | |
| Algeria | 0.87 (0.77, 0.98) | 0.021 | 0.99 (0.90, 1.08) | 0.78 | Reference | 1.04 (0.95, 1.13) | 0.41 | 1.06 (0.96, 1.16) | 0.24 |
| Iraq | 1.44 (1.14, 1.83) | 0.0027 | 1.05 (0.84, 1.31) | 0.69 | | 0.80 (0.64, 1.00) | 0.045 | 1.08 (0.84, 1.40) | 0.54 |
| State of Palestine | 1.12 (0.90, 1.39) | 0.33 | 1.07 (0.84, 1.36) | 0.60 | | 0.93 (0.75, 1.15) | 0.50 | 0.88 (0.72, 1.08) | 0.23 |
| Tunisia | 1.26 (0.94, 1.68) | 0.12 | 0.96 (0.74, 1.24) | 0.73 | | 1.19 (0.94, 1.51) | 0.16 | 0.79 (0.60, 1.05) | 0.10 |
| **Pooled PR (95% CI)** | **1.13 (0.91, 1.40)** | N/A | **1.00 (0.92, 1.08)** | N/A | | **0.98 (0.84, 1.15)** | N/A | **0.97 (0.83, 1.12)** | N/A |
| **South Asia** | | | | | | | | | |
| Afghanistan | 1.06 (0.94, 1.20) | 0.31 | 1.02 (0.94, 1.11) | 0.59 | Reference | 1.02 (0.94, 1.12) | 0.60 | 1.10 (0.99, 1.22) | 0.068 |
| Bangladesh | 1.05 (0.93, 1.19) | 0.44 | 0.89 (0.79, 1.00) | 0.057 | | 0.91 (0.81, 1.02) | 0.10 | 0.84 (0.74, 0.95) | 0.0058 |
| Nepal | 2.60 (1.94, 3.47) | <0.0001 | 1.11 (0.82, 1.51) | 0.50 | | 0.95 (0.70, 1.28) | 0.72 | 0.59 (0.38, 0.93) | 0.024 |
| Pakistan (Balochistan) | 0.85 (0.71, 1.03) | 0.10 | 1.02 (0.87, 1.18) | 0.84 | | 0.57 (0.48, 0.68) | <0.0001 | 0.42 (0.34, 0.51) | <0.0001 |
| Pakistan (Khyber Pakhtunkhwa) | 0.78 (0.68, 0.90) | 0.00 | 0.79 (0.70, 0.89) | 0.0001 | | 1.06 (0.95, 1.20) | 0.29 | 1.05 (0.91, 1.20) | 0.50 |
| Pakistan (Sindh) | 1.02 (0.94, 1.11) | 0.64 | 1.08 (1.02, 1.15) | 0.014 | | 0.69 (0.63, 0.74) | <0.0001 | 0.46 (0.42, 0.52) | <0.0001 |
| Pakistan (Punjab) | 0.82 (0.76, 0.88) | <0.0001 | 0.88 (0.83, 0.94) | 0.0003 | | 1.05 (0.98, 1.12) | 0.19 | 1.00 (0.92, 1.09) | 0.98 |
| **Pooled PR (95% CI)** | **1.06 (0.79, 1.42)** | N/A | **0.95 (0.87, 1.04)** | N/A | | **0.87 (0.73, 1.04)** | N/A | **0.74 (0.55, 1.00)** | N/A |
| **East Asia and the Pacific** | | | | | | | | | |
| Fiji | 1.20 (1.00, 1.45) | 0.054 | 1.02 (0.84, 1.24) | 0.86 | Reference | 0.96 (0.79, 1.16) | 0.67 | 1.02 (0.85, 1.21) | 0.87 |
| Kiribati | 0.77 (0.56, 1.05) | 0.094 | 0.83 (0.64, 1.06) | 0.13 | | 0.89 (0.64, 1.22) | 0.46 | 0.75 (0.54, 1.04) | 0.08 |
| Lao PDR | 0.90 (0.75, 1.08) | 0.25 | 1.21 (1.04, 1.41) | 0.01 | | 0.93 (0.81, 1.06) | 0.28 | 0.93 (0.77, 1.12) | 0.42 |
| Mongolia | 1.23 (0.76, 2.01) | 0.40 | 0.71 (0.40, 1.26) | 0.24 | | 0.65 (0.36, 1.17) | 0.15 | 0.93 (0.52, 1.68) | 0.81 |
| Samoa | 1.32 (0.90, 1.93) | 0.16 | 1.36 (0.95, 1.94) | 0.09 | | 1.00 (0.65, 1.55) | 0.99 | 1.36 (0.96, 1.94) | 0.08 |
| Tonga | 1.17 (0.81, 1.71) | 0.40 | 0.88 (0.61, 1.28) | 0.51 | | 0.87 (0.64, 1.19) | 0.37 | 0.86 (0.58, 1.28) | 0.45 |
| Tuvalu | 0.92 (0.52, 1.61) | 0.77 | 1.04 (0.61, 1.76) | 0.88 | | 0.75 (0.41, 1.38) | 0.35 | 0.75 (0.39, 1.44) | 0.38 |
| Vietnam | 1.30 (0.86, 1.96) | 0.22 | 1.18 (0.81, 1.73) | 0.39 | | 0.83 (0.53, 1.30) | 0.41 | 0.85 (0.58, 1.25) | 0.42 |
| **Pooled PR (95% CI)** | **1.06 (0.91, 1.25)** | N/A | **1.04 (0.89, 1.21)** | N/A | | **0.90 (0.81, 1.01)** | N/A | **0.94 (0.81, 1.09)** | N/A |
| **Europe and Central Asia** | | | | | | | | | |
| Kosovo | 1.59 (1.18, 2.14) | 0.0022 | 1.22 (0.89, 1.66) | 0.21 | Reference | 0.98 (0.73, 1.33) | 0.91 | 1.10 (0.84, 1.45) | 0.48 |
| Kyrgyzstan | 0.50 (0.32, 0.77) | 0.0016 | 0.62 (0.39, 0.98) | 0.039 | | 0.86 (0.60, 1.23) | 0.41 | 1.27 (0.92, 1.76) | 0.15 |

| | | | | | | | | | |
|---|---|---|---|---|---|---|---|---|---|
| Montenegro | 2.19 (0.94, 5.10) | 0.070 | 2.56 (1.00, 6.54) | 0.050 | | 2.38 (0.99, 5.73) | 0.053 | 2.68 (1.16, 6.21) | 0.021 |
| Republic of North Macedonia | 1.78 (1.02, 3.12) | 0.042 | 1.02 (0.57, 1.83) | 0.95 | | 0.67 (0.29, 1.53) | 0.34 | 0.46 (0.23, 0.91) | 0.026 |
| Serbia | 1.43 (0.93, 2.18) | 0.10 | 1.04 (0.68, 1.57) | 0.87 | | 1.01 (0.66, 1.53) | 0.98 | 0.94 (0.60, 1.46) | 0.77 |
| Turkmenistan | 4.36 (1.20, 15.78) | 0.025 | 1.37 (0.31, 6.02) | 0.68 | | 1.23 (0.18, 8.35) | 0.83 | 2.46 (0.69, 8.70) | 0.16 |
| Uzbekistan | 2.11 (1.36, 3.27) | 0.0009 | 1.43 (0.89, 2.28) | 0.14 | | 1.03 (0.64, 1.64) | 0.91 | 0.79 (0.49, 1.28) | 0.34 |
| **Pooled PR (95% CI)** | **1.55 (0.97, 2.47)** | N/A | **1.12 (0.82, 1.55)** | N/A | | **1.01 (0.76, 1.34)** | N/A | **1.09 (0.71, 1.68)** | N/A |
| **Latin America and Caribbean** | | | | | | | | | |
| Costa Rica | 1.21 (0.86, 1.71) | 0.28 | 1.16 (0.83, 1.63) | 0.38 | | 0.68 (0.42, 1.12) | 0.13 | 0.64 (0.39, 1.05) | 0.078 |
| Cuba | 0.85 (0.65, 1.12) | 0.26 | 0.87 (0.71, 1.08) | 0.21 | | 0.94 (0.76, 1.16) | 0.57 | 0.82 (0.65, 1.04) | 0.095 |
| Dominican Republic | 0.88 (0.76, 1.01) | 0.065 | 0.97 (0.86, 1.10) | 0.66 | | 0.88 (0.77, 1.01) | 0.078 | 0.82 (0.71, 0.96) | 0.013 |
| Guyana | 0.99 (0.80, 1.23) | 0.95 | 0.85 (0.66, 1.09) | 0.19 | | 0.86 (0.67, 1.09) | 0.20 | 0.85 (0.66, 1.08) | 0.18 |
| Honduras | 1.19 (1.02, 1.38) | 0.025 | 1.04 (0.91, 1.20) | 0.58 | Reference | 0.94 (0.81, 1.08) | 0.38 | 0.88 (0.75, 1.03) | 0.11 |
| Suriname | 0.88 (0.68, 1.13) | 0.31 | 0.93 (0.73, 1.19) | 0.58 | | 0.95 (0.74, 1.22) | 0.67 | 0.95 (0.71, 1.27) | 0.73 |
| Turks & Caicos Islands | 0.69 (0.25, 1.88) | 0.45 | 0.97 (0.41, 2.31) | 0.95 | | 1.23 (0.35, 4.29) | 0.74 | 1.66 (0.68, 4.04) | 0.25 |
| **Pooled PR (95% CI)** | **0.98 (0.85, 1.12)** | N/A | **0.97 (0.88, 1.06)** | N/A | | **0.90 (0.81, 1.00)** | N/A | **0.86 (0.72, 1.01)** | N/A |
| **All regions** | | | | | | | | | |
| **Pooled PR (95% CI)** | **1.07 (0.98, 1.17)** | N/A | **1.00 (0.95, 1.05)** | N/A | Reference | **0.95 (0.89, 1.00)** | N/A | **0.92 (0.84, 1.01)** | N/A |

All values are weighted to reflect the survey sampling design. Women who answered “Don’t know/no such activity” or had missing data/no response for menstrual-related absenteeism were excluded from the analysis. P values not adjusted for multiple testing.
PR = Prevalence Ratio, CI = Confidence Interval.

**Table S22: Multivariable analysis between wealth quintile and menstrual-related absenteeism.**

| Country | Wealth quintile | | | | | | | | |
|---|---|---|---|---|---|---|---|---|---|
| | Quintile 1 (Poorest) | | Quintile 2 | | Quintile 3 | Quintile 4 | | Quintile 5 (Richest) | |
| | PR (95% CI) | P value | PR (95% CI) | P value | | PR (95% CI) | P value | PR (95% CI) | P value |
| **West and Central Africa** | | | | | | | | | |
| Central African Republic | 1.09 (0.92, 1.30) | 0.32 | 1.14 (0.97, 1.33) | 0.11 | Reference | 0.91 (0.78, 1.08) | 0.28 | 0.80 (0.67, 0.96) | 0.02 |
| Chad | 1.02 (0.91, 1.15) | 0.68 | 0.99 (0.90, 1.10) | 0.92 | | 1.09 (0.99, 1.20) | 0.088 | 0.90 (0.79, 1.02) | 0.11 |
| Democratic Republic of Congo | 1.07 (0.85, 1.33) | 0.57 | 1.03 (0.86, 1.22) | 0.76 | | 0.91 (0.71, 1.15) | 0.41 | 0.79 (0.63, 0.98) | 0.03 |
| Gambia | 0.91 (0.76, 1.10) | 0.32 | 0.94 (0.80, 1.11) | 0.46 | | 1.11 (0.95, 1.30) | 0.18 | 1.12 (0.95, 1.33) | 0.18 |
| Ghana | 0.97 (0.80, 1.17) | 0.72 | 1.23 (0.98, 1.54) | 0.070 | | 0.93 (0.77, 1.12) | 0.45 | 0.87 (0.71, 1.05) | 0.14 |
| Guinea-Bissau | 1.24 (0.89, 1.72) | 0.20 | 1.22 (0.91, 1.62) | 0.18 | | 1.29 (0.96, 1.73) | 0.085 | 1.43 (1.00, 2.06) | 0.05 |
| Nigeria | 0.94 (0.82, 1.07) | 0.33 | 0.92 (0.82, 1.03) | 0.14 | | 0.94 (0.83, 1.07) | 0.36 | 0.90 (0.77, 1.05) | 0.16 |
| Sao Tome & Principe | 1.06 (0.69, 1.63) | 0.79 | 0.89 (0.59, 1.33) | 0.56 | | 1.21 (0.78, 1.86) | 0.40 | 1.22 (0.89, 1.66) | 0.22 |
| Sierra Leone | 1.29 (1.08, 1.55) | 0.0044 | 1.01 (0.87, 1.18) | 0.86 | | 1.07 (0.90, 1.27) | 0.46 | 0.92 (0.75, 1.14) | 0.45 |
| Togo | 1.04 (0.79, 1.37) | 0.77 | 1.04 (0.81, 1.35) | 0.75 | | 0.88 (0.67, 1.15) | 0.35 | 0.81 (0.62, 1.04) | 0.10 |
| **Pooled PR (95% CI)** | **1.04 (0.96, 1.13)** | N/A | **1.02 (0.95, 1.10)** | N/A | | **1.01 (0.93, 1.10)** | N/A | **0.94 (0.83, 1.05)** | N/A |
| **Eastern and Southern Africa** | | | | | | | | | |
| Lesotho | 1.05 (0.82, 1.35) | 0.69 | 0.97 (0.74, 1.27) | 0.84 | Reference | 1.04 (0.74, 1.44) | 0.83 | 1.00 (0.75, 1.33) | 1.00 |
| Madagascar | 0.99 (0.80, 1.22) | 0.91 | 1.02 (0.81, 1.29) | 0.87 | | 1.05 (0.83, 1.34) | 0.67 | 1.60 (1.27, 2.01) | <0.0001 |
| Malawi | 0.99 (0.85, 1.16) | 0.91 | 1.03 (0.89, 1.20) | 0.70 | | 1.01 (0.86, 1.18) | 0.89 | 0.89 (0.77, 1.04) | 0.14 |
| Zimbabwe | 1.13 (0.93, 1.37) | 0.21 | 1.11 (0.93, 1.32) | 0.26 | | 0.92 (0.77, 1.09) | 0.33 | 0.90 (0.75, 1.07) | 0.24 |
| **Pooled PR (95% CI)** | **1.03 (0.93, 1.15)** | N/A | **1.04 (0.94, 1.15)** | N/A | | **0.99 (0.89, 1.10)** | N/A | **1.06 (0.81, 1.38)** | N/A |
| **Middle East and North Africa** | | | | | | | | | |
| Algeria | 0.88 (0.78, 0.99) | 0.033 | 1.01 (0.92, 1.11) | 0.87 | Reference | 1.03 (0.95, 1.12) | 0.50 | 1.02 (0.93, 1.12) | 0.69 |
| Iraq | 1.44 (1.13, 1.82) | 0.0029 | 1.05 (0.84, 1.32) | 0.67 | | 0.80 (0.64, 0.99) | 0.045 | 1.08 (0.83, 1.40) | 0.59 |
| State of Palestine | 1.12 (0.90, 1.40) | 0.31 | 1.06 (0.83, 1.35) | 0.62 | | 0.93 (0.75, 1.15) | 0.50 | 0.88 (0.71, 1.08) | 0.21 |
| Tunisia | 1.26 (0.94, 1.68) | 0.12 | 0.96 (0.74, 1.24) | 0.73 | | 1.19 (0.94, 1.51) | 0.15 | 0.80 (0.60, 1.05) | 0.11 |
| **Pooled PR (95% CI)** | **1.13 (0.91, 1.40)** | N/A | **1.01 (0.94, 1.10)** | N/A | | **0.98 (0.83, 1.15)** | N/A | **0.95 (0.83, 1.09)** | N/A |
| **South Asia** | | | | | | | | | |
| Afghanistan | 1.07 (0.95, 1.20) | 0.29 | 1.02 (0.94, 1.11) | 0.59 | Reference | 1.02 (0.94, 1.12) | 0.60 | 1.10 (0.99, 1.22) | 0.062 |
| Bangladesh | 1.06 (0.94, 1.20) | 0.32 | 0.90 (0.80, 1.01) | 0.07 | | 0.91 (0.81, 1.02) | 0.094 | 0.83 (0.73, 0.94) | 0.0045 |
| Nepal | 2.61 (1.96, 3.49) | <0.0001 | 1.11 (0.82, 1.51) | 0.50 | | 0.95 (0.70, 1.29) | 0.73 | 0.60 (0.38, 0.94) | 0.03 |
| Pakistan (Balochistan) | 0.86 (0.72, 1.04) | 0.12 | 1.02 (0.88, 1.19) | 0.78 | | 0.57 (0.49, 0.68) | <0.0001 | 0.41 (0.34, 0.51) | <0.0001 |
| Pakistan (Khyber Pakhtunkhwa) | 0.79 (0.69, 0.90) | 0.0006 | 0.79 (0.70, 0.89) | 0.0001 | | 1.07 (0.95, 1.20) | 0.28 | 1.05 (0.92, 1.20) | 0.48 |
| Pakistan (Sindh) | 1.02 (0.94, 1.10) | 0.69 | 1.08 (1.01, 1.15) | 0.02 | | 0.69 (0.63, 0.74) | <0.0001 | 0.47 (0.42, 0.52) | <0.0001 |
| Pakistan (Punjab) | 0.83 (0.77, 0.90) | <0.0001 | 0.89 (0.83, 0.95) | 0.0004 | | 1.05 (0.98, 1.12) | 0.20 | 1.00 (0.92, 1.08) | 0.97 |
| **Pooled PR (95% CI)** | **1.07 (0.80, 1.43)** | N/A | **0.96 (0.87, 1.05)** | N/A | | **0.87 (0.73, 1.04)** | N/A | **0.74 (0.55, 1.00)** | N/A |
| **East Asia and the Pacific** | | | | | | | | | |
| Fiji | 1.20 (0.99, 1.44) | 0.061 | 1.02 (0.84, 1.24) | 0.85 | Reference | 0.96 (0.79, 1.16) | 0.65 | 1.02 (0.85, 1.21) | 0.86 |
| Kiribati | 0.77 (0.57, 1.05) | 0.10 | 0.82 (0.64, 1.05) | 0.12 | | 0.88 (0.64, 1.21) | 0.45 | 0.74 (0.54, 1.02) | 0.066 |
| Lao PDR | 0.90 (0.75, 1.09) | 0.27 | 1.21 (1.04, 1.40) | 0.015 | | 0.93 (0.81, 1.07) | 0.30 | 0.92 (0.77, 1.11) | 0.38 |
| Mongolia | 1.26 (0.78, 2.05) | 0.34 | 0.71 (0.40, 1.26) | 0.24 | | 0.65 (0.36, 1.17) | 0.15 | 0.94 (0.52, 1.69) | 0.84 |
| Samoa | 1.33 (0.90, 1.97) | 0.15 | 1.36 (0.95, 1.96) | 0.093 | | 1.00 (0.65, 1.54) | 1.00 | 1.32 (0.93, 1.87) | 0.12 |
| Tonga | 1.17 (0.80, 1.71) | 0.40 | 0.89 (0.61, 1.28) | 0.52 | | 0.87 (0.64, 1.19) | 0.39 | 0.84 (0.56, 1.27) | 0.41 |
| Tuvalu | 0.92 (0.52, 1.62) | 0.77 | 1.03 (0.61, 1.75) | 0.90 | | 0.75 (0.41, 1.38) | 0.36 | 0.75 (0.39, 1.44) | 0.38 |
| Vietnam | 1.37 (0.90, 2.08) | 0.14 | 1.19 (0.81, 1.74) | 0.38 | | 0.84 (0.53, 1.32) | 0.45 | 0.87 (0.60, 1.28) | 0.48 |
| **Pooled PR (95% CI)** | **1.07 (0.91, 1.26)** | N/A | **1.03 (0.88, 1.21)** | N/A | | **0.91 (0.81, 1.01)** | N/A | **0.93 (0.81, 1.08)** | N/A |
| **Europe and Central Asia** | | | | | | | | | |
| Kosovo | 1.62 (1.20, 2.17) | 0.0015 | 1.23 (0.90, 1.67) | 0.19 | Reference | 0.99 (0.74, 1.34) | 0.96 | 1.11 (0.84, 1.46) | 0.46 |
| Kyrgyzstan | 0.51 (0.33, 0.79) | 0.0026 | 0.63 (0.41, 0.99) | 0.046 | | 0.86 (0.60, 1.23) | 0.41 | 1.16 (0.84, 1.59) | 0.36 |
| Montenegro | 2.15 (0.91, 5.10) | 0.081 | 2.53 (1.00, 6.45) | 0.051 | | 2.41 (1.00, 5.84) | 0.051 | 2.77 (1.19, 6.47) | 0.019 |

| | | | | | | | | | |
|---|---|---|---|---|---|---|---|---|---|
| Republic of North Macedonia | 1.80 (1.00, 3.23) | 0.048 | 1.04 (0.57, 1.90) | 0.90 | | 0.67 (0.29, 1.53) | 0.34 | 0.46 (0.23, 0.91) | 0.026 |
| Serbia | 1.44 (0.93, 2.22) | 0.10 | 1.05 (0.69, 1.59) | 0.82 | | 1.02 (0.67, 1.56) | 0.92 | 0.96 (0.62, 1.49) | 0.86 |
| Turkmenistan | 4.33 (1.20, 15.65) | 0.026 | 1.39 (0.31, 6.12) | 0.66 | | 1.27 (0.19, 8.55) | 0.80 | 2.61 (0.75, 9.08) | 0.13 |
| Uzbekistan | 2.05 (1.32, 3.17) | 0.0013 | 1.42 (0.88, 2.27) | 0.15 | | 1.03 (0.64, 1.65) | 0.91 | 0.77 (0.47, 1.25) | 0.29 |
| **Pooled PR (95% CI)** | **1.54 (0.98, 2.44)** | N/A | **1.13 (0.83, 1.55)** | N/A | | **1.02 (0.76, 1.35)** | N/A | **1.09 (0.70, 1.70)** | N/A |
| **Latin America and Caribbean** | | | | | | | | | |
| Costa Rica | 1.22 (0.86, 1.74) | 0.26 | 1.15 (0.82, 1.62) | 0.41 | | 0.68 (0.41, 1.11) | 0.12 | 0.62 (0.38, 1.02) | 0.061 |
| Cuba | 0.86 (0.65, 1.13) | 0.27 | 0.87 (0.71, 1.08) | 0.21 | | 0.94 (0.76, 1.15) | 0.53 | 0.81 (0.64, 1.02) | 0.075 |
| Dominican Republic | 0.89 (0.77, 1.02) | 0.10 | 0.98 (0.87, 1.11) | 0.77 | | 0.88 (0.76, 1.01) | 0.068 | 0.81 (0.70, 0.95) | 0.0086 |
| Guyana | 1.01 (0.81, 1.25) | 0.95 | 0.85 (0.67, 1.08) | 0.18 | | 0.85 (0.67, 1.08) | 0.19 | 0.85 (0.67, 1.10) | 0.21 |
| Honduras | 1.19 (1.02, 1.38) | 0.024 | 1.04 (0.90, 1.19) | 0.58 | Reference | 0.93 (0.80, 1.08) | 0.34 | 0.86 (0.73, 1.01) | 0.059 |
| Suriname | 0.88 (0.69, 1.13) | 0.32 | 0.92 (0.72, 1.18) | 0.52 | | 0.94 (0.73, 1.21) | 0.64 | 0.92 (0.69, 1.22) | 0.55 |
| Turks & Caicos Islands | 0.80 (0.30, 2.18) | 0.65 | 0.96 (0.37, 2.53) | 0.94 | | 1.31 (0.38, 4.48) | 0.66 | 1.57 (0.64, 3.86) | 0.31 |
| **Pooled PR (95% CI)** | **0.99 (0.87, 1.12)** | N/A | **0.97 (0.88, 1.06)** | N/A | | **0.90 (0.80, 1.00)** | N/A | **0.84 (0.72, 0.99)** | N/A |
| **All regions** | | | | | | | | | |
| **Pooled PR (95% CI)** | **1.09 (1.00, 1.19)** | N/A | **1.01 (0.96, 1.06)** | N/A | Reference | **0.94 (0.89, 1.00)** | N/A | **0.91 (0.83, 0.99)** | N/A |

All values are weighted to reflect the survey sampling design. Women who answered “Don’t know/no such activity” or had missing data/no response for menstrual-related absenteeism were excluded from the analysis. P values not adjusted for multiple testing. Results are adjusted for women’s age and marital status.
PR = Prevalence Ratio, CI = Confidence Interval.

**Table S23: Univariable and multivariable analyses between area type (urban/rural) and menstrual-related absenteeism.**

| Country | Prevalence of menstrual-related absenteeism – n/N (%) | | Univariable | | Multivariable* | |
|---|---|---|---|---|---|---|
| | Urban | Rural | PR (95% CI) | P value | PR (95% CI) | P value |
| **West and Central Africa** | | | | | | |
| Central African Republic | 778/2802 (27.7) | 1431/4290 (33.4) | 1.21 (1.04, 1.40) | 0.016 | 1.03 (0.86, 1.25) | 0.73 |
| Chad | 1181/3893 (30.3) | 4761/14318 (33.3) | 1.09 (0.95, 1.26) | 0.22 | 1.01 (0.82, 1.24) | 0.93 |
| Democratic Republic of Congo | 1229/8959 (13.7) | 1184/8028 (14.7) | 1.07 (0.90, 1.27) | 0.43 | 0.87 (0.68, 1.13) | 0.30 |
| Gambia | 1900/8843 (21.5) | 564/3334 (16.9) | 0.79 (0.69, 0.89) | 0.0003 | 0.89 (0.75, 1.05) | 0.17 |
| Ghana | 1200/6636 (18.1) | 1226/6219 (19.7) | 1.09 (0.90, 1.31) | 0.37 | 1.01 (0.79, 1.29) | 0.94 |
| Guinea-Bissau | 425/4464 (9.5) | 467/6449 (7.2) | 0.76 (0.59, 0.99) | 0.04 | 0.78 (0.56, 1.08) | 0.14 |
| Nigeria | 2604/15993 (16.3) | 2898/17202 (16.8) | 1.04 (0.92, 1.17) | 0.55 | 1.05 (0.91, 1.21) | 0.49 |
| Sao Tome & Principe | 195/1928 (10.1) | 118/930 (12.7) | 1.27 (0.98, 1.66) | 0.071 | 1.30 (0.99, 1.70) | 0.055 |
| Sierra Leone | 1374/6922 (19.9) | 1386/6778 (20.4) | 1.04 (0.90, 1.21) | 0.58 | 0.93 (0.73, 1.19) | 0.57 |
| Togo | 353/3062 (11.5) | 384/3018 (12.7) | 1.10 (0.91, 1.35) | 0.33 | 0.98 (0.74, 1.29) | 0.88 |
| **Pooled** | **17.5 (12.7, 22.4)** | **18.6 (12.7, 24.4)** | **1.03 (0.93, 1.14)** | N/A | **0.98 (0.90, 1.08)** | N/A |
| **Eastern and Southern Africa** | | | | | | |
| Lesotho | 342/2738 (12.5) | 400/2909 (13.8) | 1.10 (0.90, 1.35) | 0.35 | 1.17 (0.89, 1.54) | 0.25 |
| Madagascar | 399/3951 (10.1) | 769/10109 (7.6) | 0.75 (0.64, 0.90) | 0.0013 | 0.96 (0.77, 1.21) | 0.75 |
| Malawi | 408/3989 (10.2) | 2192/16509 (13.3) | 1.32 (1.08, 1.60) | 0.0060 | 1.46 (1.18, 1.81) | 0.0004 |
| Zimbabwe | 538/3470 (15.5) | 854/5072 (16.8) | 1.09 (0.96, 1.23) | 0.20 | 0.95 (0.80, 1.14) | 0.61 |
| **Pooled** | **12.1 (9.6, 14.6)** | **12.8 (9.1, 16.6)** | **1.04 (0.83, 1.30)** | N/A | **1.12 (0.91, 1.36)** | N/A |
| **Middle East and North Africa** | | | | | | |
| Algeria | 5310/21058 (25.2) | 2718/12020 (22.6) | 0.90 (0.81, 0.99) | 0.034 | 0.93 (0.84, 1.04) | 0.21 |
| Iraq | 1394/13961 (10.0) | 701/5773 (12.1) | 1.22 (0.99, 1.49) | 0.063 | 1.08 (0.89, 1.31) | 0.44 |
| State of Palestine (West Bank and Gaza) | 710/4974 (14.3) | 101/948 (10.7) | 0.75 (0.60, 0.94) | 0.011 | 0.78 (0.63, 0.97) | 0.03 |
| Tunisia | 400/3937 (10.2) | 223/1731 (12.9) | 1.28 (1.01, 1.60) | 0.038 | 1.30 (1.01, 1.66) | 0.04 |
| **Pooled** | **14.9 (7.9, 21.9)** | **14.6 (9.3, 19.9)** | **1.01 (0.79, 1.28)** | N/A | **1.00 (0.81, 1.22)** | N/A |
| **South Asia** | | | | | | |
| Afghanistan | 3556/11249 (31.6) | 8607/29007 (29.7) | 0.95 (0.87, 1.04) | 0.26 | 0.97 (0.85, 1.11) | 0.67 |
| Bangladesh | 869/13742 (6.3) | 3704/44456 (8.3) | 1.37 (1.26, 1.49) | <0.0001 | 1.37 (1.23, 1.54) | <0.0001 |
| Nepal | 805/9393 (8.6) | 460/4052 (11.3) | 1.32 (1.02, 1.71) | 0.032 | 0.85 (0.66, 1.11) | 0.23 |
| Pakistan (Balochistan) | 1246/8638 (14.4) | 5109/23756 (21.5) | 1.51 (1.16, 1.98) | 0.0025 | 1.17 (0.85, 1.61) | 0.34 |
| Pakistan (Khyber Pakhtunkhwa) | 1184/6352 (18.6) | 4906/31152 (15.7) | 0.87 (0.73, 1.04) | 0.12 | 0.94 (0.78, 1.14) | 0.54 |
| Pakistan (Sindh) | 4692/15102 (31.1) | 5712/12296 (46.5) | 1.53 (1.43, 1.63) | <0.0001 | 1.09 (1.00, 1.17) | 0.043 |
| Pakistan (Punjab) | 4587/26508 (17.3) | 6802/41983 (16.2) | 0.94 (0.89, 1.00) | 0.056 | 1.02 (0.95, 1.09) | 0.60 |
| **Pooled** | **18.3 (10.9, 25.6)** | **21.3 (11.6, 31.0)** | **1.18 (0.98, 1.41)** | N/A | **1.06 (0.94, 1.19)** | N/A |
| **East Asia and the Pacific** | | | | | | |
| Fiji | 632/2942 (21.5) | 460/1784 (25.8) | 1.20 (1.02, 1.41) | 0.026 | 1.16 (0.98, 1.38) | 0.086 |
| Kiribati | 356/2080 (17.1) | 209/1439 (14.5) | 0.85 (0.68, 1.05) | 0.14 | 0.69 (0.46, 1.04) | 0.078 |
| Lao PDR† | 953/7896 (12.1) | 1692/14450 (12.5) | 0.98 (0.85, 1.14) | 0.80 | 0.89 (0.76, 1.06) | 0.19 |
| Mongolia | 202/6604 (3.1) | 101/2885 (3.5) | 1.15 (0.85, 1.56) | 0.37 | 0.85 (0.51, 1.41) | 0.52 |
| Samoa | 70/805 (8.7) | 276/3053 (9.0) | 1.04 (0.79, 1.38) | 0.78 | 1.07 (0.79, 1.45) | 0.65 |
| Tonga | 85/640 (13.2) | 333/2038 (16.3) | 1.23 (0.90, 1.68) | 0.18 | 1.18 (0.87, 1.61) | 0.28 |
| Tuvalu | 69/497 (13.9) | 45/231 (19.3) | 1.38 (0.94, 2.01) | 0.10 | 1.29 (0.87, 1.91) | 0.19 |
| Vietnam | 179/3836 (4.7) | 225/6311 (3.6) | 0.77 (0.55, 1.07) | 0.12 | 0.64 (0.45, 0.91) | 0.012 |
| **Pooled** | **11.7 (7.4, 16.0)** | **12.8 (7.5, 18.1)** | **1.05 (0.91, 1.20)** | N/A | **0.96 (0.80, 1.16)** | N/A |
| **Europe and Central Asia** | | | | | | |
| Kosovo | 230/2136 (10.8) | 295/2885 (10.2) | 0.98 (0.81, 1.18) | 0.85 | 0.93 (0.76, 1.13) | 0.46 |
| Kyrgyzstan | 189/2080 (9.1) | 173/3096 (5.6) | 0.62 (0.49, 0.78) | <0.0001 | 0.81 (0.61, 1.07) | 0.13 |
| Montenegro | 102/1478 (6.9) | 44/678 (6.5) | 0.94 (0.58, 1.53) | 0.81 | 0.95 (0.55, 1.64) | 0.85 |
| Republic of North Macedonia | 116/1922 (6.0) | 84/1100 (7.6) | 1.26 (0.78, 2.03) | 0.34 | 0.77 (0.43, 1.39) | 0.39 |
| Serbia | 185/2215 (8.4) | 138/1312 (10.5) | 1.26 (0.96, 1.65) | 0.094 | 1.18 (0.87, 1.59) | 0.29 |
| Turkmenistan | 19/2240 (0.9) | 24/2706 (0.9) | 1.03 (0.48, 2.21) | 0.94 | 0.77 (0.33, 1.77) | 0.54 |
| Uzbekistan | 154/2066 (7.5) | 161/2295 (7.0) | 0.94 (0.72, 1.22) | 0.65 | 0.73 (0.56, 0.96) | 0.023 |
| **Pooled** | **7.0 (4.7, 9.4)** | **6.8 (4.4, 9.3)** | **0.95 (0.78, 1.16)** | N/A | **0.88 (0.76, 1.03)** | N/A |
| **Latin America and Caribbean** | | | | | | |
| Costa Rica | 317/4886 (6.5) | 145/1909 (7.6) | 1.17 (0.90, 1.52) | 0.24 | 1.01 (0.79, 1.29) | 0.96 |
| Cuba | 1362/5395 (25.2) | 908/2847 (31.9) | 1.27 (1.07, 1.51) | 0.0069 | 1.40 (1.13, 1.72) | 0.0018 |
| Dominican Republic | 3393/15878 (21.4) | 1130/4798 (23.5) | 1.10 (0.95, 1.27) | 0.20 | 1.10 (0.94, 1.28) | 0.23 |
| Guyana | 295/1320 (22.4) | 801/4090 (19.6) | 0.87 (0.69, 1.11) | 0.26 | 0.86 (0.68, 1.10) | 0.23 |
| Honduras | 1558/8664 (18.0) | 1825/8908 (20.5) | 1.14 (1.01, 1.30) | 0.040 | 1.04 (0.88, 1.21) | 0.67 |
| Suriname | 847/4866 (17.4) | 208/1099 (18.9) | 1.09 (0.88, 1.34) | 0.43 | 1.11 (0.90, 1.37) | 0.33 |
| Turks & Caicos Islands | 100/760 (13.1) | 1/30 (3.6) | 0.27 (0.10, 0.77) | 0.017 | 0.30 (0.11, 0.85) | 0.026 |
| **Pooled** | **17.7 (13.0, 22.4)** | **17.9 (10.8, 24.9)** | **1.01 (0.74, 1.39)** | N/A | **1.00 (0.75, 1.34)** | N/A |
| **All regions** | | | | | | |
| **Pooled** | **19.9 (17.7, 22.0)** | **20.4 (18.4, 22.5)** | **1.05 (0.98, 1.13)** | N/A | **1.00 (0.94, 1.07)** | N/A |

All values are weighted to reflect the survey sampling design. Women who answered “Don’t know/no such activity” or had missing data/no response for menstrual-related absenteeism were excluded from the analysis. P values not adjusted for multiple testing. The reference category is urban residence.
PR = Prevalence Ratio, CI = Confidence Interval.
* Multivariable analyses adjusted for women’s age and marital status.
† Area response options in Lao PDR included urban, rural with road, and rural without road. These responses were recoded to urban, and rural (includes both rural with road and rural without road).

**Table S24: Univariable and multivariable analyses between use of menstrual materials and menstrual-related absenteeism.**

| Country | Prevalence of menstrual-related absenteeism – n/N (%) | | Univariable | | Multivariable* | |
|---|---|---|---|---|---|---|
| | Menstrual materials | No menstrual materials | PR (95% CI) | P value | PR (95% CI) | P value |
| **West and Central Africa** | | | | | | |
| Central African Republic | 2146/6745 (31.8) | 62/346 (18.1) | 1.75 (1.31, 2.35) | 0.0002 | 1.74 (1.30, 2.33) | 0.0002 |
| Chad | 5790/17293 (33.5) | 146/901 (16.3) | 2.03 (1.66, 2.49) | <0.0001 | 2.01 (1.64, 2.46) | <0.0001 |
| Democratic Republic of Congo | 2243/16066 (14.0) | 150/890 (16.8) | 0.84 (0.63, 1.10) | 0.20 | 0.88 (0.66, 1.17) | 0.37 |
| Gambia | 2415/11958 (20.2) | 49/216 (22.5) | 0.89 (0.62, 1.28) | 0.52 | 0.93 (0.64, 1.36) | 0.72 |
| Ghana | 2331/12583 (18.5) | 95/270 (35.3) | 0.52 (0.40, 0.69) | <0.0001 | 0.52 (0.39, 0.70) | <0.0001 |
| Guinea-Bissau† | – | – | – | – | – | – |
| Nigeria | 5362/32202 (16.7) | 140/981 (14.3) | 1.15 (0.92, 1.43) | 0.23 | 1.15 (0.92, 1.44) | 0.21 |
| Sao Tome & Principe‡ | – | – | – | – | – | – |
| Sierra Leone | 2702/13305 (20.3) | 55/389 (14.0) | 1.46 (1.07, 2.01) | 0.019 | 1.46 (1.06, 2.01) | 0.020 |
| Togo | 704/5859 (12.0) | 31/214 (14.3) | 0.83 (0.57, 1.22) | 0.35 | 0.85 (0.58, 1.25) | 0.42 |
| **Pooled** | **20.8 (15.4, 26.2)** | **17.9 (13.4, 22.4)** | **1.09 (0.80, 1.50)** | N/A | **1.11 (0.82, 1.51)** | N/A |
| **Eastern and Southern Africa** | | | | | | |
| Lesotho | 727/5539 (13.1) | 15/108 (13.7) | 0.96 (0.61, 1.51) | 0.86 | 0.94 (0.60, 1.47) | 0.78 |
| Madagascar | 1102/13186 (8.4) | 66/873 (7.5) | 1.11 (0.77, 1.60) | 0.57 | 1.00 (0.69, 1.45) | 0.99 |
| Malawi | 2537/19954 (12.7) | 64/543 (11.8) | 1.09 (0.73, 1.62) | 0.67 | 1.08 (0.73, 1.60) | 0.71 |
| Zimbabwe | 1359/8355 (16.3) | 34/185 (18.1) | 0.90 (0.61, 1.33) | 0.59 | 0.95 (0.65, 1.39) | 0.79 |
| **Pooled** | **12.6 (9.4, 15.8)** | **12.0 (7.6, 16.3)** | **1.02 (0.83, 1.25)** | N/A | **0.99 (0.81, 1.21)** | N/A |
| **Middle East and North Africa** | | | | | | |
| Algeria | 7339/31262 (23.5) | 684/1764 (38.8) | 0.58 (0.52, 0.65) | <0.0001 | 0.57 (0.51, 0.64) | <0.0001 |
| Iraq | 2013/16110 (12.5) | 82/686 (11.9) | 1.05 (0.74, 1.49) | 0.78 | 1.00 (0.72, 1.41) | 0.98 |
| State of Palestine (West Bank and Gaza) | 872/6219 (14.0) | 21/205 (10.0) | 1.39 (0.92, 2.10) | 0.12 | 1.35 (0.89, 2.05) | 0.16 |
| Tunisia | 606/5440 (11.1) | 17/226 (7.4) | 1.49 (0.92, 2.43) | 0.11 | 1.46 (0.90, 2.39) | 0.13 |
| **Pooled** | **15.3 (9.8, 20.8)** | **17.0 (2.7, 31.3)** | **1.02 (0.66, 1.56)** | N/A | **0.99 (0.65, 1.51)** | N/A |
| **South Asia** | | | | | | |
| Afghanistan | 10963/36921 (29.7) | 1195/3300 (36.2) | 0.83 (0.74, 0.94) | 0.0034 | 0.83 (0.74, 0.94) | 0.0028 |
| Bangladesh | 4485/56148 (8.0) | 88/2037 (4.3) | 1.83 (1.25, 2.67) | 0.0017 | 1.91 (1.31, 2.80) | 0.0008 |
| Nepal | 1079/12627 (8.5) | 186/816 (22.8) | 0.37 (0.30, 0.47) | <0.0001 | 0.46 (0.37, 0.57) | <0.0001 |
| Pakistan (Balochistan) | 4356/20844 (20.9) | 1951/10847 (18.0) | 1.22 (1.09, 1.37) | 0.0008 | 1.40 (1.25, 1.57) | <0.0001 |
| Pakistan (Khyber Pakhtunkhwa) | 5850/35054 (16.7) | 238/2213 (10.8) | 1.66 (1.38, 1.99) | <0.0001 | 1.65 (1.38, 1.98) | <0.0001 |
| Pakistan (Sindh) | 8596/22335 (38.5) | 1795/4986 (36.0) | 1.07 (1.00, 1.14) | 0.041 | 1.24 (1.17, 1.32) | <0.0001 |
| Pakistan (Punjab) | 10451/60905 (17.2) | 934/7370 (12.7) | 1.34 (1.23, 1.47) | <0.0001 | 1.30 (1.18, 1.43) | <0.0001 |
| **Pooled** | **19.9 (11.7, 28.1)** | **20.0 (10.9, 29.2)** | **1.07 (0.72, 1.58)** | N/A | **1.15 (0.81, 1.63)** | N/A |
| **East Asia and the Pacific** | | | | | | |
| Fiji | 1061/4594 (23.1) | 32/130 (24.7) | 0.94 (0.68, 1.30) | 0.70 | 0.98 (0.71, 1.35) | 0.89 |
| Kiribati | 555/3451 (16.1) | 11/65 (16.6) | 0.97 (0.55, 1.71) | 0.92 | 0.96 (0.54, 1.70) | 0.89 |
| Lao PDR | 2170/18281 (11.9) | 476/4063 (11.7) | 0.99 (0.86, 1.13) | 0.87 | 0.97 (0.85, 1.12) | 0.69 |
| Mongolia | 285/8669 (3.3) | 18/809 (2.2) | 1.45 (0.76, 2.75) | 0.26 | 1.47 (0.78, 2.78) | 0.23 |
| Samoa | 316/3541 (8.9) | 29/310 (9.5) | 0.95 (0.60, 1.48) | 0.81 | 0.96 (0.61, 1.51) | 0.86 |
| Tonga | 402/2522 (15.9) | 15/156 (9.9) | 1.62 (0.88, 2.97) | 0.12 | 1.63 (0.88, 3.03) | 0.12 |
| Tuvalu | 104/692 (15.0) | 10/36 (27.1) | 0.55 (0.32, 0.96) | 0.037 | 0.54 (0.31, 0.95) | 0.034 |
| Vietnam | 393/9959 (3.9) | 11/186 (5.7) | 0.69 (0.34, 1.40) | 0.31 | 0.71 (0.36, 1.42) | 0.33 |
| **Pooled** | **12.2 (7.6, 16.9)** | **12.3 (6.4, 18.1)** | **0.96 (0.76, 1.23)** | N/A | **0.97 (0.76, 1.24)** | N/A |
| **Europe and Central Asia** | | | | | | |
| Kosovo | 517/4979 (10.4) | 8/38 (20.9) | 0.45 (0.24, 0.82) | 0.0092 | 0.48 (0.26, 0.88) | 0.019 |
| Kyrgyzstan | 338/5018 (6.7) | 24/157 (15.2) | 0.45 (0.28, 0.71) | 0.0007 | 0.42 (0.27, 0.67) | 0.0003 |
| Montenegro | 143/2092 (6.9) | 2/58 (3.4) | 2.03 (0.53, 7.80) | 0.30 | 1.98 (0.52, 7.55) | 0.31 |
| Republic of North Macedonia | 197/2980 (6.6) | 2/43 (4.9) | 1.36 (0.37, 5.04) | 0.65 | 1.23 (0.34, 4.44) | 0.75 |
| Serbia | 319/3469 (9.2) | 4/58 (6.8) | 1.36 (0.55, 3.36) | 0.51 | 1.39 (0.56, 3.47) | 0.48 |
| Turkmenistan | 41/4902 (0.8) | 2/43 (5.8) | 0.14 (0.03, 0.63) | 0.010 | 0.19 (0.04, 0.84) | 0.028 |
| Uzbekistan§ | 303/4209 (7.2) | 12/149 (7.8) | 0.92 (0.47, 1.80) | 0.81 | 0.98 (0.50, 1.89) | 0.95 |
| **Pooled** | **6.8 (4.6, 9.1)** | **8.2 (3.8, 12.7)** | **0.71 (0.37, 1.35)** | N/A | **0.73 (0.40, 1.31)** | N/A |
| **Latin America and Caribbean** | | | | | | |
| Costa Rica | 457/6710 (6.8) | 5/79 (6.0) | 1.15 (0.44, 2.97) | 0.78 | 1.19 (0.46, 3.07) | 0.72 |
| Cuba | 2231/8048 (27.7) | 39/187 (21.0) | 1.32 (0.83, 2.09) | 0.23 | 1.32 (0.83, 2.11) | 0.24 |
| Dominican Republic | 4442/20330 (21.8) | 81/321 (25.4) | 0.85 (0.67, 1.08) | 0.20 | 0.87 (0.68, 1.10) | 0.25 |
| Guyana | 1031/5219 (19.8) | 64/188 (34.1) | 0.58 (0.39, 0.86) | 0.0075 | 0.58 (0.39, 0.86) | 0.0067 |
| Honduras | 3289/17240 (19.1) | 92/327 (28.2) | 0.67 (0.55, 0.83) | 0.0002 | 0.68 (0.56, 0.83) | 0.0002 |
| Suriname | 1018/5983 (17.0) | 108/442 (24.4) | 0.69 (0.51, 0.95) | 0.025 | 0.68 (0.50, 0.93) | 0.016 |
| Turks & Caicos Islands | 99/785 (12.6) | 2/5 (39.0) | 0.32 (0.07, 1.53) | 0.15 | 0.31 (0.07, 1.41) | 0.12 |
| **Pooled** | **17.9 (12.9, 22.8)** | **22.9 (15.5, 30.2)** | **0.78 (0.58, 1.04)** | N/A | **0.78 (0.57, 1.05)** | N/A |
| **All regions** | | | | | | |
| **Pooled** | **15.3 (12.9, 17.7)** | **16.1 (13.2, 19.0)** | **0.96 (0.83, 1.10)** | N/A | **0.97 (0.85, 1.11)** | N/A |

All values are weighted to reflect the survey sampling design. Women who answered "Don't know/no such activity" or had missing data/no response for menstrual-related absenteeism were excluded from the analysis. P values not adjusted for multiple testing. The reference category is no use of menstrual materials.
PR = Prevalence Ratio, CI = Confidence Interval.
* Multivariable analyses adjusted for wealth, education, and area type.
† Guinea-Bissau was excluded as the MICS report did not consider estimates for the use of menstrual materials to reflect the reality of Guinea-Bissau due to an error that was detected when adapting the Computer-Assisted Personal Interviewing (CAPI) to tablets.
‡ Sao Tome and Principe (2019) asked about the use of pads, tampons, and pieces of towels in three separate questions. Women who reported using pads, tampons, and/or pieces of towels on any of these questions were categorised as having used menstrual materials. Sao Tome and Principe was excluded from the pooled prevalence estimates, prevalence ratios, and pooled prevalence ratios (both univariable and multivariable) for both West and Central Africa and by all regions due to complete separation (prevalence of menstrual-related absenteeism was 313/2845 (11.0%) in women using menstrual materials, and 0/13 (0.0%) in women not using menstrual materials).
§ The multivariable prevalence ratio for Uzbekistan was obtained by the marginal standardisation technique and CIs using the delta method after a logistic model instead of Log Binomial model due to convergence issues.

**Table S25: Univariable and multivariable analyses between use of reusable menstrual materials and menstrual-related absenteeism.**

| Country | Prevalence of menstrual-related absenteeism – n/N (%) | | Univariable | | Multivariable* | |
|---|---|---|---|---|---|---|
| | Reusable | Only non-reusable | PR (95% CI) | P value | PR (95% CI) | P value |
| **West and Central Africa** | | | | | | |
| Central African Republic | 1391/4388 (31.7) | 747/2333 (32.0) | 0.99 (0.88, 1.11) | 0.85 | 0.84 (0.74, 0.95) | 0.0065 |
| Chad | 5100/14681 (34.7) | 707/2669 (26.5) | 1.31 (1.16, 1.46) | <0.0001 | 1.30 (1.15, 1.48) | <0.0001 |
| Democratic Republic of Congo | 1304/9444 (13.8) | 938/6613 (14.2) | 0.97 (0.84, 1.13) | 0.73 | 0.86 (0.70, 1.05) | 0.15 |
| Gambia | 1306/7077 (18.5) | 1107/4865 (22.8) | 0.81 (0.73, 0.90) | 0.0002 | 0.98 (0.88, 1.09) | 0.70 |
| Ghana | 337/1614 (20.9) | 1994/10968 (18.2) | 1.14 (0.96, 1.37) | 0.14 | 1.13 (0.93, 1.37) | 0.21 |
| Guinea-Bissau† | – | – | – | – | – | – |
| Nigeria | 2548/13702 (18.6) | 2804/18451 (15.2) | 1.23 (1.11, 1.36) | 0.0001 | 1.31 (1.18, 1.46) | <0.0001 |
| Sao Tome & Principe | 245/2371 (10.3) | 14/68 (21.2) | 0.48 (0.27, 0.88) | 0.02 | 0.53 (0.29, 0.98) | 0.041 |
| Sierra Leone | 1776/9257 (19.2) | 925/4040 (22.9) | 0.85 (0.74, 0.97) | 0.02 | 0.74 (0.63, 0.86) | 0.0001 |
| Togo | 396/3479 (11.4) | 307/2378 (12.9) | 0.88 (0.72, 1.08) | 0.22 | 0.82 (0.64, 1.06) | 0.13 |
| **Pooled** | **19.8 (14.4, 25.3)** | **20.5 (16.3, 24.7)** | **0.97 (0.83, 1.15)** | N/A | **0.95 (0.80, 1.13)** | N/A |
| **Eastern and Southern Africa** | | | | | | |
| Lesotho | 67/427 (15.8) | 660/5106 (12.9) | 1.22 (0.94, 1.58) | 0.13 | 1.26 (0.95, 1.67) | 0.11 |
| Madagascar | 796/10262 (7.8) | 305/2918 (10.4) | 0.74 (0.64, 0.87) | 0.0002 | 0.92 (0.76, 1.11) | 0.39 |
| Malawi | 1746/14039 (12.4) | 791/5914 (13.4) | 0.91 (0.81, 1.03) | 0.12 | 0.91 (0.80, 1.03) | 0.12 |
| Zimbabwe | 311/1845 (16.9) | 1048/6511 (16.1) | 1.05 (0.92, 1.19) | 0.46 | 0.99 (0.86, 1.14) | 0.90 |
| **Pooled** | **13.0 (9.0, 17.1)** | **13.2 (11, 15.5)** | **0.95 (0.78, 1.16)** | N/A | **0.98 (0.86, 1.13)** | N/A |
| **Middle East and North Africa** | | | | | | |
| Algeria | 423/1665 (25.4) | 7015/29805 (23.5) | 1.08 (0.95, 1.24) | 0.23 | 1.20 (1.05, 1.36) | 0.0069 |
| Iraq | 285/1871 (15.2) | 1728/14238 (12.1) | 1.25 (1.03, 1.53) | 0.026 | 1.19 (0.98, 1.45) | 0.086 |
| State of Palestine (West Bank and Gaza) | 28/137 (20.6) | 836/6048 (13.8) | 1.48 (1.00, 2.21) | 0.053 | 1.45 (0.97, 2.17) | 0.071 |
| Tunisia | 37/207 (18.1) | 568/5229 (10.9) | 1.69 (1.21, 2.36) | 0.0019 | 1.57 (1.11, 2.22) | 0.010 |
| **Pooled** | **19.8 (15.2, 24.4)** | **15.1 (9.5, 20.7)** | **1.29 (1.06, 1.58)** | N/A | **1.27 (1.10, 1.47)** | N/A |
| **South Asia** | | | | | | |
| Afghanistan | 9021/30226 (29.8) | 1938/6671 (29.0) | 1.06 (0.96, 1.17) | 0.23 | 1.11 (0.99, 1.23) | 0.062 |
| Bangladesh | 2838/38504 (7.4) | 1643/17573 (9.4) | 0.82 (0.76, 0.89) | <0.0001 | 0.71 (0.65, 0.78) | <0.0001 |
| Nepal | 775/7925 (9.8) | 303/4700 (6.5) | 1.52 (1.23, 1.87) | <0.0001 | 1.08 (0.89, 1.32) | 0.42 |
| Pakistan (Balochistan) | 2419/9750 (24.8) | 1899/10830 (17.5) | 1.45 (1.21, 1.73) | <0.0001 | 1.24 (1.03, 1.50) | 0.023 |
| Pakistan (Khyber Pakhtunkhwa) | 4825/27935 (17.3) | 1022/7061 (14.5) | 1.24 (1.11, 1.40) | 0.0003 | 1.31 (1.17, 1.47) | <0.0001 |
| Pakistan (Sindh) | 5073/11363 (44.6) | 3496/10830 (32.3) | 1.42 (1.33, 1.51) | <0.0001 | 1.04 (0.98, 1.10) | 0.16 |
| Pakistan (Punjab) | 5059/31425 (16.1) | 5385/29447 (18.3) | 0.88 (0.84, 0.92) | <0.0001 | 0.91 (0.87, 0.96) | 0.0008 |
| **Pooled** | **21.4 (11.8, 30.9)** | **18.2 (11.1, 25.2)** | **1.16 (0.96, 1.39)** | N/A | **1.03 (0.89, 1.20)** | N/A |
| **East Asia and the Pacific** | | | | | | |
| Fiji | 134/556 (24.2) | 926/4035 (23.0) | 1.05 (0.88, 1.25) | 0.59 | 0.96 (0.79, 1.16) | 0.65 |
| Kiribati | 81/564 (14.4) | 473/2886 (16.4) | 0.88 (0.68, 1.13) | 0.31 | 0.91 (0.71, 1.18) | 0.48 |
| Lao PDR | 53/632 (8.4) | 2114/17623 (12.0) | 0.69 (0.53, 0.91) | 0.0081 | 0.66 (0.50, 0.87) | 0.0029 |
| Mongolia | 20/245 (8.2) | 264/8404 (3.1) | 2.60 (1.46, 4.61) | 0.0012 | 2.29 (1.24, 4.23) | 0.0082 |
| Samoa | 59/693 (8.4) | 258/2846 (9.1) | 0.93 (0.71, 1.21) | 0.58 | 0.95 (0.73, 1.23) | 0.68 |
| Tonga | 7/25 (26.6) | 395/2488 (15.9) | 1.67 (0.73, 3.82) | 0.22 | 1.58 (0.69, 3.60) | 0.27 |
| Tuvalu | 22/128 (17.0) | 82/563 (14.6) | 1.16 (0.74, 1.83) | 0.52 | 1.08 (0.67, 1.76) | 0.75 |
| Vietnam | 8/120 (6.4) | 385/9839 (3.9) | 1.80 (0.78, 4.12) | 0.17 | 1.85 (0.79, 4.32) | 0.15 |
| **Pooled** | **12.9 (8.0, 17.9)** | **12.2 (7.5, 16.8)** | **1.13 (0.84, 1.53)** | N/A | **1.07 (0.81, 1.43)** | N/A |
| **Europe and Central Asia** | | | | | | |
| Kosovo | 19/162 (11.7) | 498/4813 (10.4) | 1.23 (0.81, 1.88) | 0.34 | 1.11 (0.71, 1.72) | 0.65 |
| Kyrgyzstan | 54/933 (5.7) | 284/4073 (7.0) | 0.82 (0.60, 1.14) | 0.24 | 1.02 (0.72, 1.43) | 0.92 |
| Montenegro | 6/87 (6.8) | 138/2002 (6.9) | 0.99 (0.29, 3.38) | 0.99 | 0.99 (0.30, 3.31) | 0.99 |
| Republic of North Macedonia | 3/26 (9.8) | 195/2953 (6.6) | 1.48 (0.40, 5.56) | 0.56 | 1.38 (0.38, 5.03) | 0.62 |
| Serbia | 5/18 (30.2) | 314/3451 (9.1) | 3.32 (1.11, 9.88) | 0.031 | 3.10 (1.04, 9.21) | 0.042 |
| Turkmenistan | 0/42 (0.9)‡ | 41/4860 (0.8) | 1.08 (0.14, 8.28) | 0.94 | 1.07 (0.13, 8.52) | 0.95 |
| Uzbekistan§ | 76/624 (12.2) | 226/3578 (6.3) | 1.92 (1.47, 2.51) | <0.0001 | 1.63 (1.25, 2.14) | 0.0004 |
| **Pooled** | **8.0 (3.1, 13.0)** | **6.7 (4.4, 8.9)** | **1.35 (0.93, 1.96)** | N/A | **1.31 (0.96, 1.79)** | N/A |
| **Latin America and Caribbean** | | | | | | |
| Costa Rica | 16/131 (12.1) | 441/6548 (6.7) | 1.81 (0.91, 3.62) | 0.092 | 1.71 (0.87, 3.37) | 0.12 |
| Cuba | 74/216 (34.2) | 2155/7826 (27.5) | 1.45 (0.91, 2.32) | 0.12 | 1.42 (0.89, 2.25) | 0.14 |
| Dominican Republic | 105/455 (23.2) | 4317/19819 (21.8) | 1.07 (0.84, 1.36) | 0.57 | 1.06 (0.83, 1.36) | 0.62 |
| Guyana | 32/106 (30.2) | 998/5110 (19.5) | 1.54 (1.06, 2.24) | 0.022 | 1.46 (1.02, 2.09) | 0.036 |
| Honduras | 115/538 (21.4) | 3162/16646 (19.0) | 1.12 (0.92, 1.38) | 0.26 | 1.04 (0.85, 1.27) | 0.72 |
| Suriname | 47/227 (20.7) | 968/5747 (16.8) | 1.24 (0.87, 1.77) | 0.23 | 1.33 (0.93, 1.91) | 0.12 |
| Turks & Caicos Islands | 1/9 (16.2) | 97/758 (12.8) | 1.26 (0.23, 6.89) | 0.78 | 1.53 (0.27, 8.76) | 0.61 |
| **Pooled** | **22.1 (16.6, 27.5)** | **17.8 (12.9, 22.7)** | **1.24 (1.05, 1.46)** | N/A | **1.22 (1.03, 1.43)** | N/A |
| **All regions** | | | | | | |
| **Pooled** | **17.1 (14.5, 19.7)** | **15.0 (12.8, 17.2)** | **1.12 (1.02, 1.23)** | N/A | **1.08 (0.99, 1.17)** | N/A |

All values are weighted to reflect the survey sampling design. Women who answered "Don't know/no such activity" or had missing data/no response for menstrual-related absenteeism were excluded from the analysis. P values not adjusted for multiple testing. The reference category is no use of reusable menstrual materials.
PR = Prevalence Ratio, CI = Confidence Interval.
* Multivariate analyses adjusted for wealth, education, and area type.
† Guinea-Bissau was excluded as the MICS report did not consider estimates for the use of reusable menstrual materials to reflect the reality of Guinea-Bissau due to an error that was detected when adapting the Computer-Assisted Personal Interviewing (CAPI) to tablets.
‡ After applying survey weights, 0.37 (rounded to 0) women in Turkmenistan using reusable sanitary materials experienced menstrual-related absenteeism, representing 0.9% of the sample.
§ The multivariable prevalence ratio for Uzbekistan was obtained by the marginal standardisation technique and CIs using the delta method after a logistic model instead of Log Binomial model due to convergence issues.

**Table S26: Univariable and multivariable analyses between availability of a private place to wash at home during menstruation and menstrual-related absenteeism.**

| Country | Prevalence of menstrual-related absenteeism – n/N (%) | | Univariable | | Multivariable* | |
|---|---|---|---|---|---|---|
| | Private place to wash | No private place to wash | PR (95% CI) | P value | PR (95% CI) | P value |
| **West and Central Africa** | | | | | | |
| Central African Republic | 2138/6528 (32.8) | 70/563 (12.5) | 2.59 (1.96, 3.42) | <0.0001 | 2.60 (1.97, 3.45) | <0.0001 |
| Chad | 5793/17007 (34.1) | 147/1188 (12.4) | 2.73 (2.22, 3.35) | <0.0001 | 2.72 (2.22, 3.35) | <0.0001 |
| Democratic Republic of Congo | 2284/15352 (14.9) | 128/1607 (7.9) | 1.89 (1.36, 2.62) | 0.0002 | 1.92 (1.39, 2.66) | 0.0001 |
| Gambia | 2408/11701 (20.6) | 57/470 (12.0) | 1.70 (1.18, 2.44) | 0.0042 | 1.78 (1.24, 2.57) | 0.0019 |
| Ghana | 2343/12068 (19.4) | 83/787 (10.5) | 1.85 (1.32, 2.60) | 0.0003 | 1.88 (1.34, 2.62) | 0.0003 |
| Guinea-Bissau† | – | – | – | – | – | – |
| Nigeria | 5245/30815 (17.0) | 256/2371 (10.8) | 1.56 (1.25, 1.95) | <0.0001 | 1.56 (1.25, 1.95) | 0.0001 |
| Sao Tome & Principe | 295/2695 (10.9) | 19/162 (11.5) | 0.94 (0.58, 1.54) | 0.82 | 0.95 (0.58, 1.56) | 0.85 |
| Sierra Leone | 2669/12728 (21.0) | 87/958 (9.1) | 2.32 (1.75, 3.07) | <0.0001 | 2.35 (1.77, 3.10) | <0.0001 |
| Togo | 665/5565 (12.0) | 72/514 (14.0) | 0.86 (0.64, 1.15) | 0.30 | 0.86 (0.64, 1.16) | 0.33 |
| **Pooled** | **20.2 (14.9, 25.6)** | **10.9 (9.5, 12.3)** | **1.74 (1.33, 2.27)** | N/A | **1.76 (1.35, 2.29)** | N/A |
| **Eastern and Southern Africa** | | | | | | |
| Lesotho | 709/5349 (13.3) | 33/296 (11.1) | 1.19 (0.79, 1.79) | 0.40 | 1.19 (0.79, 1.78) | 0.41 |
| Madagascar | 1088/12765 (8.5) | 79/1291 (6.1) | 1.38 (1.05, 1.81) | 0.020 | 1.43 (1.07, 1.90) | 0.014 |
| Malawi | 2480/18968 (13.1) | 121/1528 (7.9) | 1.74 (1.31, 2.31) | 0.0001 | 1.79 (1.35, 2.37) | 0.0001 |
| Zimbabwe | 1353/8253 (16.4) | 40/290 (13.8) | 1.18 (0.83, 1.69) | 0.36 | 1.20 (0.84, 1.72) | 0.32 |
| **Pooled** | **12.8 (9.6, 16.0)** | **9.2 (5.9, 12.4)** | **1.39 (1.13, 1.70)** | N/A | **1.42 (1.15, 1.75)** | N/A |
| **Middle East and North Africa** | | | | | | |
| Algeria | 7148/29824 (24.0) | 877/3209 (27.3) | 0.86 (0.77, 0.96) | 0.0061 | 0.84 (0.76, 0.93) | 0.0014 |
| Iraq | 1780/14889 (12.0) | 313/1907 (16.4) | 0.73 (0.62, 0.86) | 0.0002 | 0.73 (0.62, 0.86) | 0.0001 |
| State of Palestine (West Bank and Gaza) | 738/5170 (14.3) | 155/1247 (12.4) | 1.15 (0.94, 1.40) | 0.17 | 1.18 (0.96, 1.44) | 0.11 |
| Tunisia | 427/3185 (13.4) | 194/2475 (7.9) | 1.71 (1.41, 2.08) | <0.0001 | 1.72 (1.42, 2.09) | <0.0001 |
| **Pooled** | **15.9 (10.6, 21.3)** | **15.9 (7.8, 24.1)** | **1.05 (0.73, 1.51)** | N/A | **1.05 (0.72, 1.52)** | N/A |
| **South Asia** | | | | | | |
| Afghanistan | 11585/36857 (31.4) | 570/3357 (17.0) | 1.80 (1.54, 2.11) | <0.0001 | 1.81 (1.54, 2.12) | <0.0001 |
| Bangladesh | 4461/56287 (7.9) | 99/1883 (5.2) | 1.51 (1.24, 1.85) | <0.0001 | 1.53 (1.26, 1.87) | <0.0001 |
| Nepal | 860/11638 (7.4) | 404/1802 (22.4) | 0.33 (0.27, 0.40) | <0.0001 | 0.44 (0.36, 0.55) | <0.0001 |
| Pakistan (Balochistan) | 4795/20954 (22.9) | 1529/10776 (14.2) | 1.68 (1.48, 1.91) | <0.0001 | 1.96 (1.72, 2.23) | <0.0001 |
| Pakistan (Khyber Pakhtunkhwa) | 5548/33236 (16.7) | 540/4018 (13.4) | 1.31 (1.15, 1.49) | <0.0001 | 1.30 (1.14, 1.47) | 0.0001 |
| Pakistan (Sindh) | 9590/22902 (41.9) | 809/4417 (18.3) | 2.27 (2.05, 2.52) | <0.0001 | 2.55 (2.31, 2.82) | <0.0001 |
| Pakistan (Punjab) | 10552/61772 (17.1) | 834/6504 (12.8) | 1.35 (1.24, 1.47) | <0.0001 | 1.34 (1.23, 1.46) | <0.0001 |
| **Pooled** | **20.7 (11.5, 30.0)** | **14.6 (10.7, 18.5)** | **1.30 (0.81, 2.07)** | N/A | **1.41 (0.93, 2.13)** | N/A |
| **East Asia and the Pacific** | | | | | | |
| Fiji | 1060/4525 (23.4) | 32/198 (15.9) | 1.48 (1.02, 2.15) | 0.041 | 1.50 (1.04, 2.18) | 0.032 |
| Kiribati | 540/3266 (16.5) | 25/252 (10.0) | 1.65 (1.05, 2.58) | 0.028 | 1.72 (1.10, 2.69) | 0.018 |
| Lao PDR | 2132/18091 (11.8) | 513/4249 (12.1) | 0.96 (0.83, 1.10) | 0.55 | 0.96 (0.83, 1.12) | 0.62 |
| Mongolia | 233/8492 (2.7) | 70/970 (7.2) | 0.38 (0.27, 0.53) | <0.0001 | 0.37 (0.27, 0.52) | <0.0001 |
| Samoa | 316/3266 (9.7) | 30/567 (5.3) | 1.81 (1.19, 2.76) | 0.0055 | 1.85 (1.20, 2.83) | 0.0052 |
| Tonga | 402/2518 (16.0) | 15/159 (9.4) | 1.70 (0.85, 3.41) | 0.13 | 1.73 (0.86, 3.48) | 0.12 |
| Tuvalu | 108/687 (15.7) | 6/40 (14.0) | 1.13 (0.51, 2.51) | 0.77 | 1.02 (0.46, 2.27) | 0.95 |
| Vietnam | 394/9848 (4.0) | 9/297 (3.2) | 1.27 (0.60, 2.68) | 0.54 | 1.28 (0.60, 2.70) | 0.52 |
| **Pooled** | **12.4 (7.6, 17.2)** | **8.9 (5.9, 11.9)** | **1.15 (0.79, 1.68)** | N/A | **1.16 (0.79, 1.70)** | N/A |
| **Europe and Central Asia** | | | | | | |
| Kosovo | 517/4943 (10.5) | 8/73 (10.8) | 0.94 (0.49, 1.79) | 0.84 | 0.98 (0.51, 1.87) | 0.95 |
| Kyrgyzstan | 309/4828 (6.4) | 53/346 (15.3) | 0.42 (0.30, 0.58) | <0.0001 | 0.38 (0.27, 0.53) | <0.0001 |
| Montenegro | 132/2094 (6.3) | 13/56 (23.5) | 0.27 (0.10, 0.70) | 0.0069 | 0.26 (0.10, 0.72) | 0.0091 |
| Republic of North Macedonia | 191/2952 (6.5) | 8/70 (11.4) | 0.57 (0.23, 1.42) | 0.22 | 0.52 (0.20, 1.31) | 0.16 |
| Serbia | 320/3489 (9.2) | 3/37 (8.3) | 1.06 (0.36, 3.10) | 0.92 | 1.06 (0.36, 3.11) | 0.92 |
| Turkmenistan | 42/4890 (0.9) | 1/55 (2.1) | 0.40 (0.08, 1.95) | 0.26 | 0.42 (0.09, 2.08) | 0.29 |
| Uzbekistan | 279/4216 (6.6) | 34/142 (24.0) | 0.28 (0.19, 0.41) | <0.0001 | 0.33 (0.23, 0.50) | <0.0001 |
| **Pooled** | **6.6 (4.3, 8.9)** | **12.2 (6.4, 18.1)** | **0.47 (0.31, 0.72)** | N/A | **0.48 (0.32, 0.73)** | N/A |
| **Latin America and Caribbean** | | | | | | |
| Costa Rica | 456/6724 (6.8) | 6/64 (8.6) | 0.78 (0.34, 1.81) | 0.57 | 0.77 (0.33, 1.76) | 0.53 |
| Cuba | 2242/7869 (28.5) | 28/364 (7.8) | 3.64 (1.76, 7.56) | 0.0005 | 3.53 (1.70, 7.32) | 0.0008 |
| Dominican Republic | 4403/19706 (22.3) | 120/945 (12.7) | 1.75 (1.33, 2.30) | 0.0001 | 1.73 (1.32, 2.27) | <0.0001 |
| Guyana | 1052/5051 (20.8) | 44/356 (12.2) | 1.71 (1.24, 2.36) | 0.0012 | 1.71 (1.24, 2.35) | 0.0010 |
| Honduras | 3317/16963 (19.6) | 65/601 (10.7) | 1.82 (1.37, 2.42) | <0.0001 | 1.84 (1.38, 2.45) | <0.0001 |
| Suriname | 1099/6185 (17.8) | 27/237 (11.6) | 1.54 (0.99, 2.39) | 0.053 | 1.51 (0.97, 2.34) | 0.070 |
| Turks & Caicos Islands | 96/761 (12.6) | 4/28 (15.8) | 0.81 (0.20, 3.31) | 0.75 | 0.84 (0.22, 3.19) | 0.79 |
| **Pooled** | **18.4 (13.2, 23.5)** | **11.1 (9.1, 13.2)** | **1.66 (1.17, 2.37)** | N/A | **1.64 (1.16, 2.33)** | N/A |
| **All regions** | | | | | | |
| **Pooled** | **15.6 (13.1, 18.1)** | **11.7 (10.3, 13.2)** | **1.22 (1.03, 1.45)** | N/A | **1.25 (1.05, 1.48)** | N/A |

All values are weighted to reflect the survey sampling design. Women who answered "Don't know/no such activity" or had missing data/no response for menstrual-related absenteeism were excluded from the analysis. P values not adjusted for multiple testing. The reference category is no available private place to wash at home during menstruation.
PR = Prevalence Ratio, CI = Confidence Interval.
* Multivariate analyses adjusted for wealth and area type.
† Guinea-Bissau was excluded as the MICS report did not consider estimates for the availability of a private place to wash at home during menstruation to reflect the reality of Guinea-Bissau due to an error that was detected when adapting the Computer-Assisted Personal Interviewing (CAPI) to tablets.

**Table S27: Univariable and multivariable analyses between current use of contraception (any method) and menstrual-related absenteeism.**

| Country | Prevalence of menstrual-related absenteeism – n/N (%) | | Univariable | | Multivariable* | |
|---|---|---|---|---|---|---|
| | Contraception | No contraception | PR (95% CI) | P value | PR (95% CI) | P value |
| **West and Central Africa** | | | | | | |
| Central African Republic | 311/1201 (25.9) | 1630/4952 (32.9) | 0.78 (0.69, 0.89) | 0.0003 | 0.84 (0.73, 0.96) | 0.012 |
| Chad | 398/1347 (29.5) | 4819/14149 (34.1) | 0.87 (0.76, 0.98) | 0.025 | 0.90 (0.79, 1.02) | 0.088 |
| Democratic Republic of Congo | 630/4540 (13.9) | 1501/10156 (14.8) | 0.94 (0.82, 1.07) | 0.33 | 1.01 (0.88, 1.16) | 0.89 |
| Gambia | 167/1416 (11.8) | 2176/9675 (22.5) | 0.52 (0.44, 0.63) | <0.0001 | 0.71 (0.59, 0.85) | 0.0002 |
| Ghana | 478/2772 (17.2) | 1750/9194 (19.0) | 0.91 (0.81, 1.02) | 0.12 | 0.93 (0.82, 1.04) | 0.21 |
| Guinea-Bissau | 269/3378 (8.0) | 572/6656 (8.6) | 0.92 (0.74, 1.15) | 0.48 | 0.84 (0.69, 1.01) | 0.064 |
| Nigeria | 806/6168 (13.1) | 4153/23785 (17.5) | 0.75 (0.67, 0.83) | <0.0001 | 0.85 (0.76, 0.95) | 0.0053 |
| Sao Tome and Principe | 71/1024 (6.9) | 222/1647 (13.5) | 0.51 (0.39, 0.67) | <0.0001 | 0.57 (0.43, 0.75) | 0.0001 |
| Sierra Leone | 703/3714 (18.9) | 1821/8750 (20.8) | 0.91 (0.80, 1.02) | 0.10 | 0.90 (0.81, 1.01) | 0.084 |
| Togo | 161/1346 (12.0) | 516/4247 (12.1) | 0.98 (0.82, 1.18) | 0.86 | 1.04 (0.86, 1.25) | 0.70 |
| **Pooled estimate (95% CI)** | **15.6 (11.1, 20.1)** | **19.5 (14.3, 24.7)** | **0.80 (0.69, 0.92)** | N/A | **0.86 (0.78, 0.95)** | N/A |
| **Eastern and Southern Africa** | | | | | | |
| Lesotho | 341/2879 (11.8) | 376/2560 (14.7) | 0.81 (0.69, 0.94) | 0.0048 | 0.83 (0.71, 0.97) | 0.020 |
| Madagascar | 321/5208 (6.2) | 740/7685 (9.6) | 0.64 (0.54, 0.76) | <0.0001 | 0.69 (0.58, 0.82) | <0.0001 |
| Malawi | 881/9766 (9.0) | 1551/9185 (16.9) | 0.55 (0.49, 0.62) | <0.0001 | 0.61 (0.55, 0.69) | <0.0001 |
| Zimbabwe | – | – | – | – | – | – |
| **Pooled** | **9.0 (5.8, 12.1)** | **13.7 (9.5, 17.9)** | **0.66 (0.53, 0.81)** | N/A | **0.70 (0.59, 0.83)** | N/A |
| **Middle East and North Africa** | | | | | | |
| Algeria | 1721/9907 (17.4) | 1182/5758 (20.5) | 0.85 (0.77, 0.93) | 0.0006 | 0.84 (0.76, 0.92) | 0.0003 |
| Iraq | 1094/10143 (10.8) | 801/7550 (10.6) | 1.02 (0.86, 1.21) | 0.83 | 1.04 (0.87, 1.23) | 0.69 |
| State of Palestine | 516/3788 (13.6) | 291/1914 (15.2) | 0.89 (0.76, 1.05) | 0.18 | 0.88 (0.74, 1.04) | 0.13 |
| Tunisia | 321/2865 (11.2) | 261/2449 (10.7) | 1.04 (0.87, 1.24) | 0.65 | 1.04 (0.87, 1.24) | 0.67 |
| **Pooled** | **13.3 (10.3, 16.2)** | **14.2 (9.6, 18.8)** | **0.93 (0.84, 1.04)** | N/A | **0.93 (0.83, 1.04)** | N/A |
| **South Asia** | | | | | | |
| Afghanistan | – | – | – | – | – | – |
| Bangladesh | 2136/30239 (7.1) | 955/12476 (7.7) | 0.90 (0.83, 0.98) | 0.012 | 0.92 (0.85, 0.99) | 0.035 |
| Nepal | 452/4750 (9.5) | 503/4911 (10.2) | 0.93 (0.81, 1.07) | 0.31 | 0.91 (0.79, 1.05) | 0.19 |
| Pakistan (Balochistan) | 609/4483 (13.6) | 2580/11504 (22.4) | 0.66 (0.55, 0.78) | <0.0001 | 0.82 (0.69, 0.97) | 0.019 |
| Pakistan (Khyber Pakhtunkhwa) | 1234/8369 (14.7) | 1849/12827 (14.4) | 1.02 (0.93, 1.12) | 0.71 | 1.01 (0.93, 1.11) | 0.77 |
| Pakistan (Sindh) | 1359/3856 (35.2) | 4612/11598 (39.8) | 0.88 (0.83, 0.94) | <0.0001 | 0.94 (0.89, 0.99) | 0.027 |
| Pakistan (Punjab) | 2321/14859 (15.6) | 3577/22592 (15.8) | 0.98 (0.92, 1.03) | 0.44 | 0.97 (0.92, 1.02) | 0.27 |
| **Pooled** | **15.9 (8.0, 23.9)** | **18.4 (9.1, 27.7)** | **0.90 (0.80, 1.01)** | N/A | **0.94 (0.89, 0.99)** | N/A |
| **East Asia and the Pacific** | | | | | | |
| Fiji | 215/1125 (19.1) | 836/3394 (24.6) | 0.77 (0.67, 0.89) | 0.0004 | 0.74 (0.64, 0.86) | 0.0001 |
| Kiribati | 133/827 (16.0) | 405/2452 (16.5) | 0.97 (0.79, 1.20) | 0.79 | 0.96 (0.78, 1.18) | 0.70 |
| Lao PDR | 904/9286 (9.7) | 1617/11966 (13.5) | 0.72 (0.66, 0.79) | <0.0001 | 0.75 (0.68, 0.83) | <0.0001 |
| Mongolia | 119/3875 (3.1) | 173/5032 (3.4) | 0.89 (0.65, 1.24) | 0.50 | 0.95 (0.66, 1.37) | 0.79 |
| Samoa | 40/387 (10.4) | 285/3208 (8.9) | 1.16 (0.81, 1.64) | 0.42 | 1.18 (0.83, 1.66) | 0.35 |
| Tonga | 64/449 (14.3) | 333/2059 (16.2) | 0.88 (0.68, 1.15) | 0.34 | 0.88 (0.66, 1.17) | 0.38 |
| Tuvalu | 17/127 (13.0) | 90/549 (16.4) | 0.79 (0.44, 1.43) | 0.43 | 0.75 (0.41, 1.36) | 0.34 |
| Vietnam | 182/5438 (3.3) | 216/4414 (4.9) | 0.68 (0.53, 0.87) | 0.0025 | 0.89 (0.67, 1.18) | 0.40 |
| **Pooled** | **10.9 (6.9, 15.0)** | **13.0 (8.1, 17.9)** | **0.83 (0.73, 0.94)** | N/A | **0.85 (0.76, 0.96)** | N/A |
| **Europe and Central Asia** | | | | | | |
| Kosovo | 204/2144 (9.5) | 308/2716 (11.4) | 0.92 (0.77, 1.09) | 0.32 | 0.97 (0.79, 1.19) | 0.77 |
| Kyrgyzstan | 78/1619 (4.8) | 265/3176 (8.3) | 0.57 (0.43, 0.76) | 0.0002 | 0.67 (0.51, 0.90) | 0.0066 |
| Montenegro | 25/380 (6.5) | 112/1676 (6.7) | 0.96 (0.50, 1.85) | 0.90 | 0.99 (0.47, 2.09) | 0.97 |
| Republic of North Macedonia | 79/1495 (5.3) | 114/1415 (8.1) | 0.65 (0.45, 0.96) | 0.030 | 0.67 (0.44, 1.01) | 0.053 |
| Serbia | 140/1807 (7.8) | 175/1626 (10.7) | 0.72 (0.55, 0.94) | 0.017 | 0.77 (0.58, 1.03) | 0.075 |
| Turkmenistan | 19/2481 (0.8) | 24/1979 (1.2) | 0.64 (0.33, 1.23) | 0.18 | 0.64 (0.33, 1.24) | 0.19 |
| Uzbekistan | 125/2001 (6.2) | 172/2049 (8.4) | 0.74 (0.58, 0.94) | 0.015 | 0.72 (0.57, 0.93) | 0.010 |
| **Pooled** | **5.7 (3.7, 7.8)** | **7.8 (5.3, 10.3)** | **0.74 (0.63, 0.86)** | N/A | **0.78 (0.67, 0.90)** | N/A |
| **Latin America and Caribbean** | | | | | | |
| Costa Rica | 241/3742 (6.4) | 211/2821 (7.5) | 0.86 (0.68, 1.10) | 0.24 | 0.76 (0.57, 1.00) | 0.048 |
| Cuba | 1610/5738 (28.1) | 600/2249 (26.7) | 1.05 (0.90, 1.23) | 0.54 | 1.05 (0.89, 1.22) | 0.57 |
| Dominican Republic | 2196/9902 (22.2) | 2147/9862 (21.8) | 1.02 (0.95, 1.09) | 0.65 | 1.03 (0.96, 1.11) | 0.37 |
| Guyana | 213/1232 (17.3) | 836/3947 (21.2) | 0.82 (0.68, 0.99) | 0.034 | 0.81 (0.68, 0.97) | 0.023 |
| Honduras | 1481/8024 (18.5) | 1732/8721 (19.9) | 0.93 (0.85, 1.01) | 0.076 | 0.92 (0.85, 1.00) | 0.060 |
| Suriname | 272/1922 (14.2) | 827/4265 (19.4) | 0.73 (0.62, 0.86) | 0.0002 | 0.73 (0.62, 0.87) | 0.0005 |
| Turks & Caicos Islands | 19/264 (7.0) | 78/505 (15.5) | 0.45 (0.24, 0.85) | 0.016 | 0.44 (0.23, 0.84) | 0.016 |
| **Pooled** | **16.2 (10.5, 22.0)** | **18.8 (14.4, 23.3)** | **0.87 (0.73, 1.03)** | N/A | **0.85 (0.71, 1.02)** | N/A |
| **All regions** | | | | | | |
| **Pooled** | **12.8 (10.6, 14.9)** | **15.4 (13.1, 17.8)** | **0.82 (0.78, 0.88)** | N/A | **0.86 (0.82, 0.90)** | N/A |

All values are weighted to reflect the survey sampling design. Women who answered "Don't know/no such activity" or had missing data/no response for menstrual-related absenteeism were excluded from the analysis. P values not adjusted for multiple testing. The reference category is no contraception use.
PR = Prevalence Ratio, CI = Confidence Interval.
* Multivariate analyses adjusted for wealth, area, age, and education.
– Contraception use not measured.

**Table S28: Univariable and multivariable analyses between use of hormonal contraception (injectables, implants, and/or the pill) and menstrual-related absenteeism.**

| Country | Prevalence of menstrual-related absenteeism – n/N (%) | | Univariable | | Multivariable | |
|---|---|---|---|---|---|---|
| | Hormonal | Non-hormonal or no contraception | PR (95% CI) | P value | PR (95% CI) | P value |
| **West and Central Africa** | | | | | | |
| Central African Republic | 165/686 (24.0) | 1776/5466 (32.5) | 0.74 (0.62, 0.89) | 0.0012 | 0.81 (0.68, 0.97) | 0.025 |
| Chad | 267/819 (32.5) | 4947/14664 (33.7) | 0.96 (0.83, 1.12) | 0.64 | 1.02 (0.88, 1.19) | 0.79 |
| Democratic Republic of Congo | 163/1322 (12.3) | 1966/13313 (14.8) | 0.83 (0.63, 1.09) | 0.19 | 0.92 (0.70, 1.21) | 0.55 |
| Gambia | 154/1298 (11.9) | 2185/9747 (22.4) | 0.53 (0.44, 0.64) | <0.0001 | 0.71 (0.59, 0.87) | 0.0008 |
| Ghana | 384/2035 (18.9) | 1833/9831 (18.6) | 1.02 (0.90, 1.16) | 0.78 | 1.03 (0.91, 1.18) | 0.61 |
| Guinea-Bissau | 137/1830 (7.5) | 656/7651 (8.6) | 0.87 (0.64, 1.18) | 0.37 | 0.82 (0.62, 1.10) | 0.19 |
| Nigeria | 474/3467 (13.7) | 4445/26189 (17.0) | 0.80 (0.70, 0.93) | 0.0026 | 0.95 (0.82, 1.09) | 0.45 |
| Sao Tome and Principe | 43/745 (5.7) | 249/1883 (13.2) | 0.43 (0.31, 0.60) | <0.0001 | 0.49 (0.35, 0.70) | <0.0001 |
| Sierra Leone | 694/3512 (19.8) | 1826/8928 (20.5) | 0.96 (0.85, 1.08) | 0.51 | 0.96 (0.86, 1.08) | 0.49 |
| Togo | 87/767 (11.3) | 584/4786 (12.2) | 0.93 (0.70, 1.22) | 0.59 | 1.04 (0.77, 1.39) | 0.81 |
| **Pooled estimate (95% CI)** | **15.6 (10.6, 20.6)** | **19.3 (14.2, 24.4)** | **0.79 (0.67, 0.94)** | N/A | **0.88 (0.76, 1.00)** | N/A |
| **Eastern and Southern Africa** | | | | | | |
| Lesotho | 175/1852 (9.5) | 528/3498 (15.1) | 0.63 (0.51, 0.78) | <0.0001 | 0.64 (0.52, 0.80) | <0.0001 |
| Madagascar | 251/4385 (5.7) | 796/8386 (9.5) | 0.60 (0.50, 0.72) | <0.0001 | 0.66 (0.55, 0.79) | <0.0001 |
| Malawi | 694/7819 (8.9) | 1731/10964 (15.8) | 0.58 (0.51, 0.65) | <0.0001 | 0.62 (0.55, 0.70) | <0.0001 |
| Zimbabwe | – | – | – | – | – | – |
| **Pooled** | **8.0 (5.7, 10.2)** | **13.4 (9.5, 17.3)** | **0.59 (0.54, 0.65)** | N/A | **0.63 (0.58, 0.69)** | N/A |
| **Middle East and North Africa** | | | | | | |
| Algeria | 1229/7336 (16.8) | 1503/7868 (19.1) | 0.88 (0.80, 0.97) | 0.0084 | 0.87 (0.79, 0.96) | 0.0048 |
| Iraq | 390/3873 (10.1) | 1324/12110 (10.9) | 0.92 (0.72, 1.17) | 0.50 | 0.88 (0.70, 1.10) | 0.25 |
| State of Palestine | 64/510 (12.6) | 531/3406 (15.6) | 0.80 (0.61, 1.05) | 0.12 | 0.78 (0.59, 1.02) | 0.065 |
| Tunisia | 189/1618 (11.6) | 235/2096 (11.2) | 1.03 (0.87, 1.24) | 0.71 | 1.02 (0.86, 1.22) | 0.81 |
| **Pooled** | **12.9 (10.0, 15.7)** | **14.2 (10.4, 18.0)** | **0.91 (0.82, 1.02)** | N/A | **0.89 (0.80, 1.00)** | N/A |
| **South Asia** | | | | | | |
| Afghanistan | – | – | – | – | – | – |
| Bangladesh | 1524/23538 (6.5) | 1534/18870 (8.1) | 0.78 (0.72, 0.84) | <0.0001 | 0.76 (0.71, 0.82) | <0.0001 |
| Nepal | 244/2323 (10.5) | 685/7097 (9.6) | 1.09 (0.91, 1.30) | 0.35 | 1.00 (0.84, 1.19) | 1.00 |
| Pakistan (Balochistan) | 339/2227 (15.2) | 2793/13596 (20.5) | 0.78 (0.65, 0.95) | 0.012 | 0.92 (0.77, 1.09) | 0.34 |
| Pakistan (Khyber Pakhtunkhwa) | 608/3986 (15.2) | 2418/16947 (14.3) | 1.08 (0.97, 1.20) | 0.15 | 1.11 (1.00, 1.23) | 0.051 |
| Pakistan (Sindh) | 578/1499 (38.5) | 5298/13647 (38.8) | 0.99 (0.91, 1.08) | 0.88 | 0.95 (0.87, 1.03) | 0.19 |
| Pakistan (Punjab) | 347/2053 (16.9) | 5354/34079 (15.7) | 1.08 (0.97, 1.20) | 0.18 | 1.10 (0.99, 1.23) | 0.082 |
| **Pooled** | **17.1 (8.2, 25.9)** | **17.8 (8.9, 26.8)** | **0.96 (0.84, 1.09)** | N/A | **0.96 (0.86, 1.08)** | N/A |
| **East Asia and the Pacific** | | | | | | |
| Fiji | 117/629 (18.5) | 915/3808 (24.0) | 0.77 (0.63, 0.93) | 0.0071 | 0.74 (0.61, 0.90) | 0.0024 |
| Kiribati | 75/460 (16.2) | 461/2804 (16.4) | 0.99 (0.78, 1.26) | 0.92 | 0.98 (0.77, 1.25) | 0.87 |
| Lao PDR | 670/6891 (9.7) | 1812/13975 (13.0) | 0.75 (0.68, 0.83) | <0.0001 | 0.78 (0.70, 0.86) | <0.0001 |
| Mongolia | 22/919 (2.4) | 191/5836 (3.3) | 0.73 (0.43, 1.22) | 0.22 | 0.73 (0.43, 1.22) | 0.23 |
| Samoa | 26/265 (9.7) | 300/3310 (9.1) | 1.07 (0.68, 1.67) | 0.77 | 1.08 (0.69, 1.70) | 0.73 |
| Tonga | 28/210 (13.4) | 366/2276 (16.1) | 0.83 (0.56, 1.25) | 0.38 | 0.83 (0.55, 1.27) | 0.39 |
| Tuvalu | 14/109 (12.4) | 93/567 (16.4) | 0.76 (0.40, 1.41) | 0.37 | 0.72 (0.38, 1.37) | 0.31 |
| Vietnam | 40/1358 (2.9) | 300/6692 (4.5) | 0.66 (0.45, 0.96) | 0.030 | 0.71 (0.48, 1.04) | 0.078 |
| **Pooled** | **10.4 (6.3, 14.5)** | **12.8 (8.0, 17.6)** | **0.80 (0.71, 0.91)** | N/A | **0.81 (0.71, 0.91)** | N/A |
| **Europe and Central Asia** | | | | | | |
| Kosovo | 8/84 (9.9) | 496/4679 (10.6) | 1.04 (0.53, 2.05) | 0.92 | 1.05 (0.54, 2.04) | 0.90 |
| Kyrgyzstan | 7/146 (4.9) | 289/3834 (7.5) | 0.65 (0.31, 1.36) | 0.25 | 0.76 (0.37, 1.59) | 0.47 |
| Montenegro | 0/32 (0.6)† | 135/1983 (6.8) | 0.09 (0.01, 0.71) | 0.023 | 0.09 (0.01, 0.71) | 0.023 |
| Republic of North Macedonia | 2/39 (4.0) | 189/2837 (6.7) | 0.59 (0.10, 3.57) | 0.57 | 0.69 (0.11, 4.19) | 0.69 |
| Serbia | 2/92 (2.0) | 312/3282 (9.5) | 0.21 (0.05, 0.86) | 0.030 | 0.22 (0.05, 0.91) | 0.037 |
| Turkmenistan | 1/65 (1.3) | 25/2196 (1.1) | 1.11 (0.15, 8.41) | 0.92 | 1.05 (0.14, 8.01) | 0.96 |
| Uzbekistan‡ | 6/97 (6.7) | 212/2465 (8.6) | 0.78 (0.32, 1.92) | 0.58 | 0.73 (0.29, 1.82) | 0.50 |
| **Pooled** | **3.3 (0.9, 5.8)** | **7.2 (5.0, 9.5)** | **0.57 (0.29, 1.13)** | N/A | **0.60 (0.30, 1.18)** | N/A |
| **Latin America and Caribbean** | | | | | | |
| Costa Rica | 99/1950 (5.1) | 348/4462 (7.8) | 0.66 (0.49, 0.89) | 0.0064 | 0.66 (0.49, 0.89) | 0.0065 |
| Cuba | 328/1053 (31.1) | 1330/5108 (26.0) | 1.19 (1.00, 1.43) | 0.055 | 1.17 (0.97, 1.41) | 0.11 |
| Dominican Republic | 1061/4734 (22.4) | 3198/14563 (22.0) | 1.02 (0.94, 1.11) | 0.65 | 1.01 (0.92, 1.10) | 0.87 |
| Guyana | 117/558 (20.9) | 916/4433 (20.7) | 1.02 (0.80, 1.29) | 0.88 | 1.01 (0.80, 1.28) | 0.91 |
| Honduras | 633/3523 (18.0) | 2445/12476 (19.6) | 0.92 (0.83, 1.01) | 0.085 | 0.90 (0.81, 0.99) | 0.025 |
| Suriname | 210/1454 (14.4) | 874/4653 (18.8) | 0.77 (0.64, 0.93) | 0.0058 | 0.77 (0.64, 0.94) | 0.0085 |
| Turks & Caicos Islands | 10/130 (7.7) | 87/631 (13.8) | 0.55 (0.20, 1.49) | 0.22 | 0.54 (0.20, 1.45) | 0.21 |
| **Pooled** | **17.1 (10.6, 23.6)** | **18.4 (14.0, 22.8)** | **0.91 (0.77, 1.09)** | N/A | **0.90 (0.76, 1.07)** | N/A |
| **All regions** | | | | | | |
| **Pooled** | **12.6 (10.2, 15.0)** | **15.1 (12.8, 17.4)** | **0.83 (0.75, 0.91)** | N/A | **0.85 (0.78, 0.93)** | N/A |

All values are weighted to reflect the survey sampling design. Women who answered "Don't know/no such activity" or had missing data/no response for menstrual-related absenteeism were excluded from the analysis. P values not adjusted for multiple testing. The reference category is use of only non-hormonal or no contraception. PR = Prevalence Ratio, CI = Confidence Interval.
* Multivariate analyses adjusted for wealth, area, age, and education.
† After applying survey weights, 0.19 (rounded to 0) women in Montenegro using hormonal contraception experienced menstrual-related absenteeism, representing 0.6% of the sample.
‡ The multivariable prevalence ratio for Uzbekistan was obtained by the marginal standardisation technique and CIs using the delta method after a logistic model instead of Log Binomial model due to convergence issues.
– Contraception use not measured.
